# The FELIX Project: Deep Networks To Detect Pancreatic Neoplasms

**DOI:** 10.1101/2022.09.24.22280071

**Authors:** Yingda Xia, Qihang Yu, Linda Chu, Satomi Kawamoto, Seyoun Park, Fengze Liu, Jieneng Chen, Zhuotun Zhu, Bowen Li, Zongwei Zhou, Yongyi Lu, Yan Wang, Wei Shen, Lingxi Xie, Yuyin Zhou, Christopher Wolfgang, Ammar Javed, Daniel Fadaei Fouladi, Shahab Shayesteh, Jefferson Graves, Alejandra Blanco, Eva S. Zinreich, Miriam Klauss, Philipp Mayer, Benedict Kinny-Köster, Kenneth Kinzler, Ralph H. Hruban, Bert Vogelstein, Alan L. Yuille, Elliot K. Fishman

## Abstract

Tens of millions of abdominal images are obtained with computed tomography (CT) in the U.S. each year but pancreatic cancers are sometimes not initially detected in these images. We here describe a suite of algorithms (named FELIX) that can recognize pancreatic lesions from CT images without human input. Using FELIX, *>*95% of patients with pancreatic ductal adenocarcinomas were detected at a specificity of *>*95% in patients without pancreatic disease. FELIX may be able to assist radiologists in identifying pancreatic cancers earlier, when surgery and other treatments offer more hope for long-term survival.

## 1 Introduction

Pancreatic ductal adenocarcinomas (PDAC) are among the deadliest of all malignancies. They typically appear as solid hypo-enhancing mass lesions on CT scans. Over 40 million abdominal CT scans are performed in the US each year, providing an opportunity for the earlier detection of pancreatic cancer. Most such CT scans are taken for reasons unrelated to suspected pancreatic neoplasia. Retrospective reviews of CT scans demonstrate that early PDACs are missed in a substantial number of scans performed before patients become symptomatic^1,2^.

Recent improvements in the power of Artificial Intelligence (AI) to identify objects in images suggest that AI might be able to assist radiologists in a variety of ways. Deep networks^3^ are the most natural form of AI for detecting and localizing cancerous tumors. They have already been applied to many types of radiographic images, including those of the pancreas (reviewed in Appendix A). But the detection of pancreatic neoplasms is especially challenging, in part because the shape of the normal pancreas is more variable than the shape of many other organs and the pancreas can move unpredictably within the abdominal cavity during the imaging process, unlike other organs such as the brain.

We here describe a suite of algorithms that have been specifically created for the purpose of detecting pancreatic cancers using deep networks. This project was commissioned by the Lustgarten Foundation for Pancreatic Cancer Research five years ago, and was named FELIX.

## 2 Results

### 2.1 Task I: Recognizing the normal pancreas and neighboring abdominal organs

The first step in developing algorithms that could recognize a pancreatic cancer is to train algorithms that recognize the normal pancreas. For this purpose, we assembled a set of 836 abdominal CT images from healthy individuals at Johns Hopkins Hospital. For each patient, there was one venous and one arterial set of images, for a total of 1,672 CT scans, each containing from 319 to 1,051 CT slices. Each set of images was manually annotated by an expert, with outlines of the pancreas drawn in all three spatial dimensions, as described in §4. In addition to the pancreas, the annotation included that of 19 neighboring abdominal organ structures because we initially expected that these other organs might subsequently be useful for distinguishing lesions within the pancreas from those of neighboring organs. It required an average of 3 hours to manually annotate the images of one healthy individual. This curated dataset of abdominal CT images from healthy individuals is unprecedented in scale, exceeding the total of all previously published abdominal CT scans used for designing deep networks^4–7^.

To recognize the pancreas and neighboring abdominal organs, we modified 3D U-Net, a basic symmetric deep network architecture consisting of encoder and decoder sub-networks. The final algorithm (FELIX 1.0) made for normal pancreas segmentation (i.e., the allocation of pixels within the image to the pancreas) or for the segmentation of other abdominal organs (e.g., allocating pixels to the liver or spleen) are detailed in §4. FELIX 1.0 took the arterial and venous phases as input, aligned them with an auto-alignment algorithm (see §4), and then applied the deep network to obtain the segmentation. But it could also be run using each phase separately. Performance was assessed by training the algorithm on a training set (531 patients) from Cohort 1, and independently validated on a test set of 305 individuals.

Previous studies showed that the pancreas is difficult to segment compared to other organs such as the liver and that its precise boundaries are hard to determine even by an expert radiologist^8–13^. The FELIX 1.0 algorithm was able to “find” and segment the pancreas in 100% of the 305 individuals in the test set. However, this 100% figure is only meaningful if the size and shape of the predicted pancreas matches that of the “ground truth”, i.e., the pancreas size and shape determined by an expert radiologist. The reliability of segmentation algorithms is often evaluated by DSC (Dice Similarity Coefficients), which are indices of spatial overlap. DSC can range from 0, indicating no spatial overlap between the ground truth and the AI prediction, to 1, indicating complete overlap. The DSC obtained by FELIX 1.0 averaged 87% (IQR 85% to 91%) and the DSC for the venous or arterial phases alone averaged 86% (IQR 83% to 91%) on the test set. The DSCs were also high on most of the 19 neighboring abdominal organs, with a liver DSC of 97% and spleen of 96%. Examples of the original CT images, the manually annotated images, and the FELIX-predicted images are shown in Figure 1.

**Figure 1.**
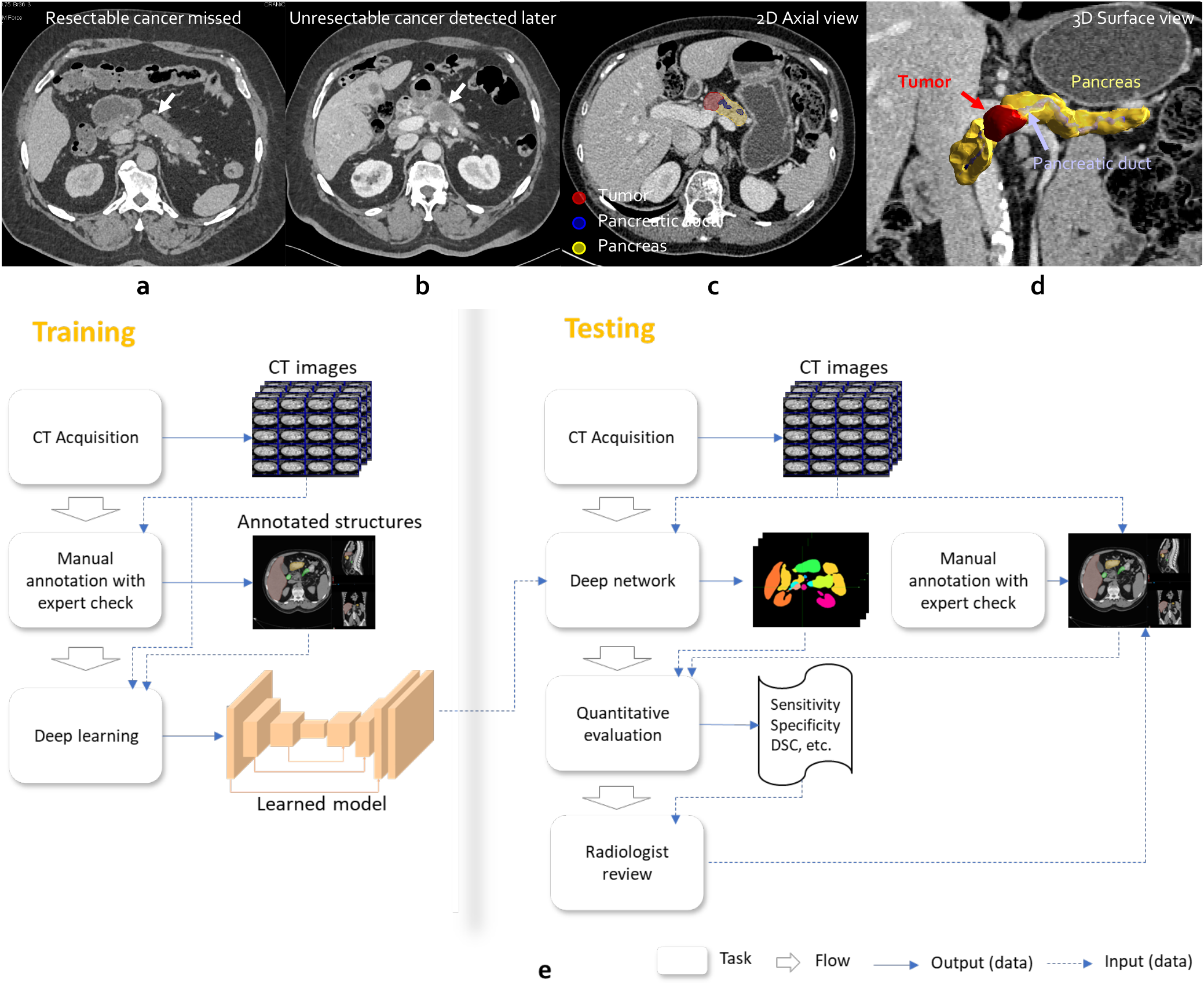
a,b: Early signs of pancreatic cancer are subtle (see arrow) and it is easy to miss a resectable (i.e., treatable) cancer. c,d: the pancreas is annotated in yellow, the PDAC tumor in red, and the pancreatic duct in blue. e: the workflow of FELIX.

### 2.2 Task II: Recognizing a PDAC within the pancreas

For this task, we assembled a set of CT images from 426 patients with PDAC from Johns Hopkins Hospital (Cohort 2, Table 1). We assessed only patients in whom the excised PDAC was confirmed through evaluation by an expert pathologist. As with the healthy individuals from Cohort 1, there was one venous and one arterial set of images from each patient in Cohort 2, for a total of 852 CT scans, and each set of images was manually annotated by an expert team (§4). This curated dataset of abdominal images from patients with PDAC, like the set from healthy individuals, is unprecedented in scale^6^.

**Table 1.** The statistics of datasets for evaluation. Detailed demographic information can be found in the attached supporting file.

| name | component | slice thickness | venous | arterial | source |
| --- | --- | --- | --- | --- | --- |
| Cohort 1 | 300 healthy individuals | 0.5mm | ✓ | ✓ | collected at Johns Hopkins Hospital |
| Cohort 2 | 213 PDAC patients | 0.5mm | ✓ | ✓ | collected at Johns Hopkins Hospital |
| Cohort 3 | 399 PDAC patients | [1.0, 5.0]mm | ✓ |  | collected at hospitals in Johns Hopkins |
| Cohort 4 | 82 healthy individuals | [1.5, 2.5]mm | ✓ |  | taken from the NIH Pancreas-CT dataset |
| Cohort 5 | 164 healthy individuals | [0.64, 2.0]mm | ✓ | ✓ | collected at Heidelberg |
| Cohort 6 | 78 PDAC patients | [0.64, 2.0]mm | ✓ | ✓ | collected at Heidelberg |
| Cohort 7 | 450 PanNET patients | 0.5mm | ✓ | ✓ | collected at Johns Hopkins Hospital |
| Cohort 8 | 458 Cyst patients | 0.5mm | ✓ | ✓ | collected at Johns Hopkins Hospital |

The AI algorithms developed for Task II were trained to predict which voxels in the images represented healthy pancreatic tissue and which represented PDAC. This task required an additional suite of algorithms, in aggregate called FELIX 1.1. A U-Net architecture was used to incorporate a “bounding box” into FELIX 1.0 that surrounded the pancreas and aligned the venous and arterial phases. Using the two aligned scans as input, FELIX 1.1 then segmented all the voxels within the bounding box as either normal or abnormal voxels. These and other components of FELIX 1.1 are detailed in §4.

FELIX 1.1 was trained on 1,592 patients from JHH, and then independently validated on images from 213 other patients.

Examples of the original CT images, the manually annotated images, and the FELIX-predicted images are shown in Figure 2. Box plots of DSC and ASSD scores to judge performance in the independent validation set of 213 patients are shown in Figure 5a and Figure 10a, respectively. The predictions had a sensitivity for detecting pancreatic cancers of 97% at a specificity of 99% (Figure 4a). The performance of the venous or arterial phases alone (sensitivity and specificity of 93% and 99%) was less than the performance of the dual-phase images. This highlighted the importance of the auto-alignment and other modules of the algorithms in FELIX 1.1 that were able to combine the arterial and venous phase images into a single, more informative set of images.

**Figure 2.**
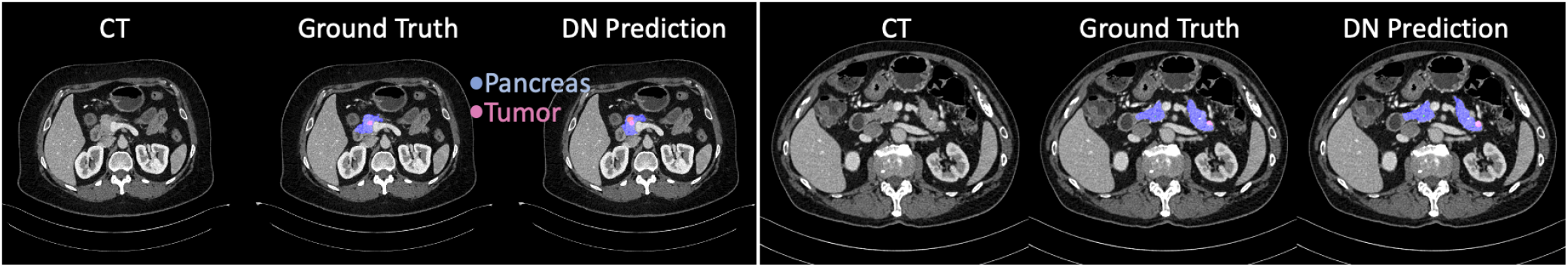
Visualization of CT scans inputs, ground-truths and our predictions.

The 97% sensitivity for detecting a PDAC within the pancreas does not fully illustrate the performance of FELIX. We defined a true positive not only as a PDAC that was predicted to exist within the pancreas, but was also localized correctly. This is quite different from what can be achieved with radiomics techniques, for example, which predict the existence of a lesion but not its location^14^. In Cohort 2, the average DSC obtained by FELIX 1.0 was 65% (IQR 58% to 85%) and the DSC for the venous or arterial phases alone averaged 63% (IQR 49% to 82%), meaning that that at least half of the pixels predicted to be PDAC were actually PDAC.

### 2.3 Task III: Recognizing PDAC in CT images from other institutions

The patients in Cohorts 1-2 were universally imaged using radiologic protocols at the Johns Hopkins Hospital on Siemens’ CT instruments. But there are well-documented cases where AI algorithms perform extremely well on datasets similar to those on which they were trained, but fail when tested on datasets from other institutions or under different conditions^15–17^. In the AI community, this is known as the domain transfer problem^18^. This problem is particularly challenging for the detection of PDACs because there are so many variables that could impact performance (see examples in Figure 3). These variables include the type and manufacturer of the CT scanner, the resolution of the scanner, the CT slice thickness, the nature and timing of the contrast dye injection, the times at which images were obtained following contrast dye injection, whether single phase (venous only) or 2-phase (arterial and venous) images are obtained, whether oral contrast as well as intravenous contrast dyes are administered, whether patients have fasted before imaging and the duration of such fasting, and the angle of the scanner with respect to the patient’s coronal axis (sometimes this axis is tilted to highlight certain abdominal organs). It would be nearly impossible to get training sets that capture the diversity of these variables as well as the heterogeneity inherent in PDAC characteristics such as size, shape, texture and location within the pancreas.

**Figure 3.**
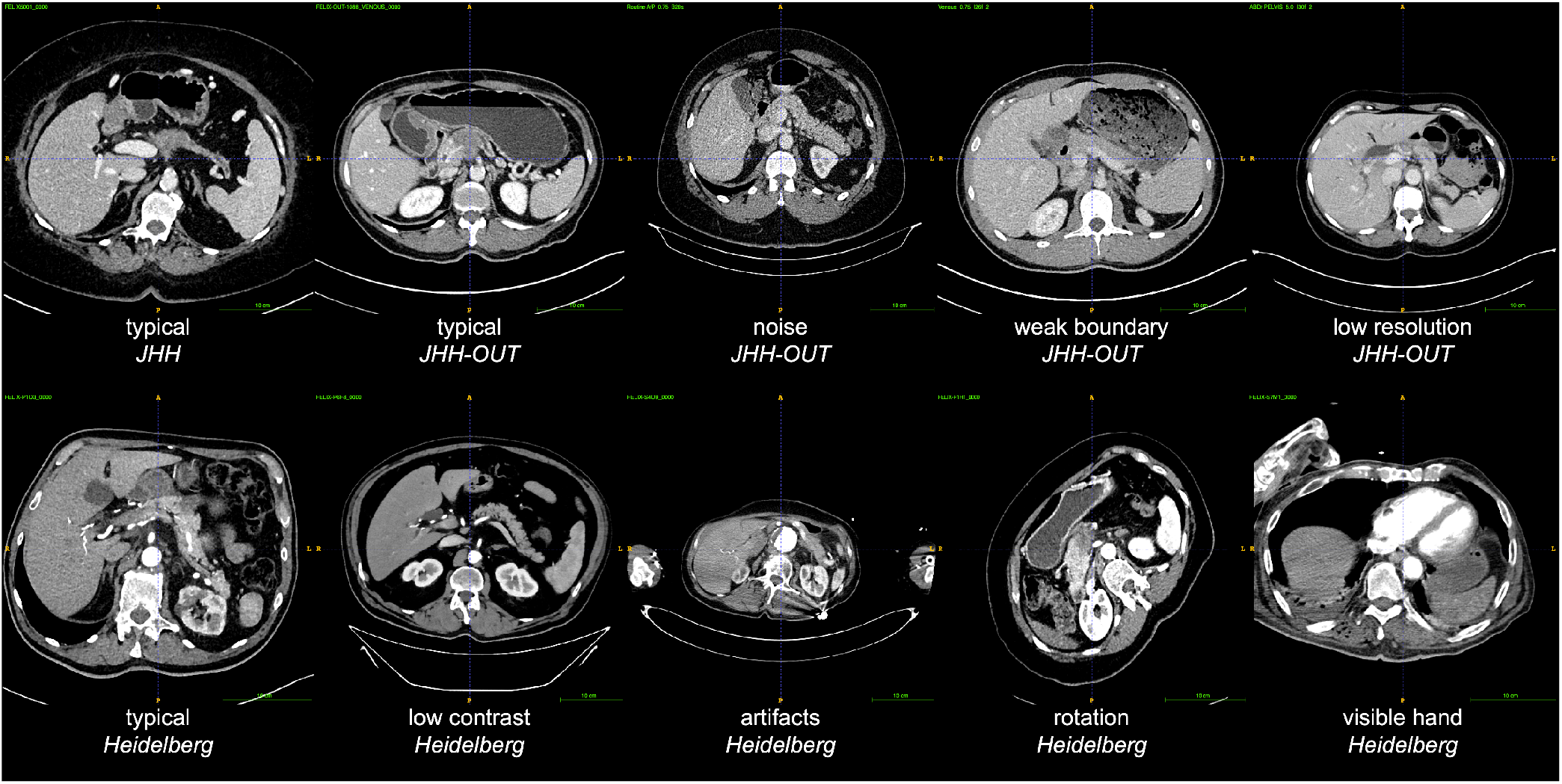
Examples of CT scans from different hospitals (domains) illustrating the variability in the CT scans caused by different scanners and protocols. In the FELIX project we trained the AI algorithms on the JHH data only and tested them on JHH data and on CT scans from other datasets, including multi-center, multi-phase, and multi-vendor cases.

To being to surmount this challenge, we artificially created a much larger training dataset by applying data augmentation techniques to the JHH training set. For example, we simulated three-dimensional rotations of the CT scans and adjustments of other scan properties such as CT slice thickness. The resultant large increase in data enabled us to train a much larger deep network simply by adding extra components to our original network rather than acquiring a much larger number of CT scans. The resulting algorithms were in aggregate called FELIX 1.2, elaborated in §4.

We assessed four other cohorts to assess the performance of FELIX 1.2 in scans from other institutions. None of the patients in these cohorts were used for training purposes. The CT scans from Cohort 3 were obtained from 399 patients with PDAC, with images taken in the U.S. but not at Johns Hopkins Hospital (Table 1). The images were acquired with GE, Siemens, Phillips, and Toshiba scanners but the slice thicknesses varied widely. Moreover, for most of the scans, only venous phase images (rather than venous plus arterial phase images) were available, and other components of the imaging protocol were often different than those performed at Johns Hopkins. Despite these differences, the sensitivities for detecting PDAC were *>*97% (Figure 4c). In Cohort 3, the average DSC for the venous phase was 58% (IQR 41% to 80%), as shown in Figure 5c.

**Figure 4.**
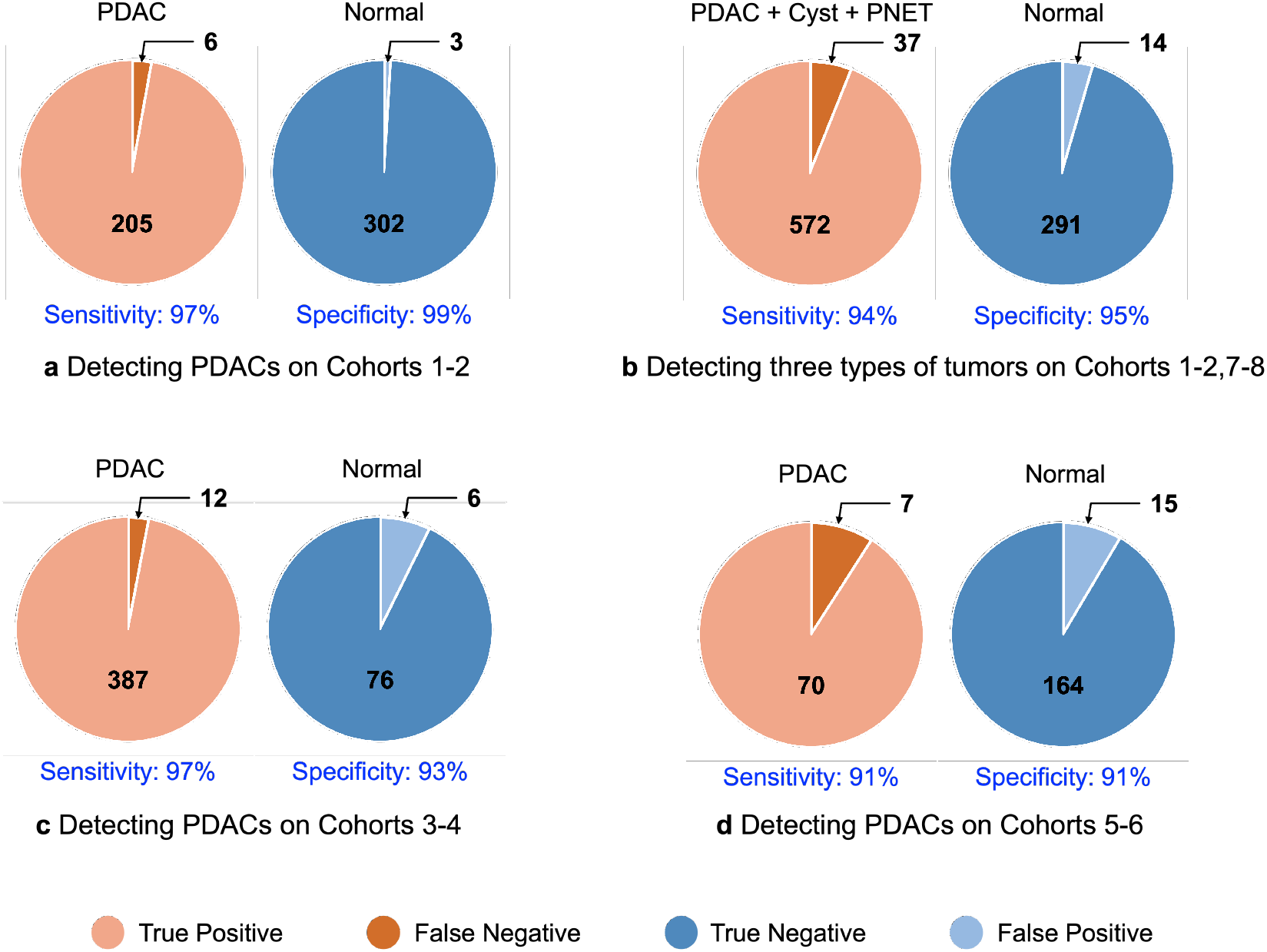
A summary of our AI algorithm performing on CT scans from different hospitals. The AI trained on JHH data performed at level close to expert radiologists on JHH test set, but performance declined somewhat on data from other hospitals. The AI algorithms were trained on 1,592 *×* 2 CT scans from JHH.

**Figure 5.**
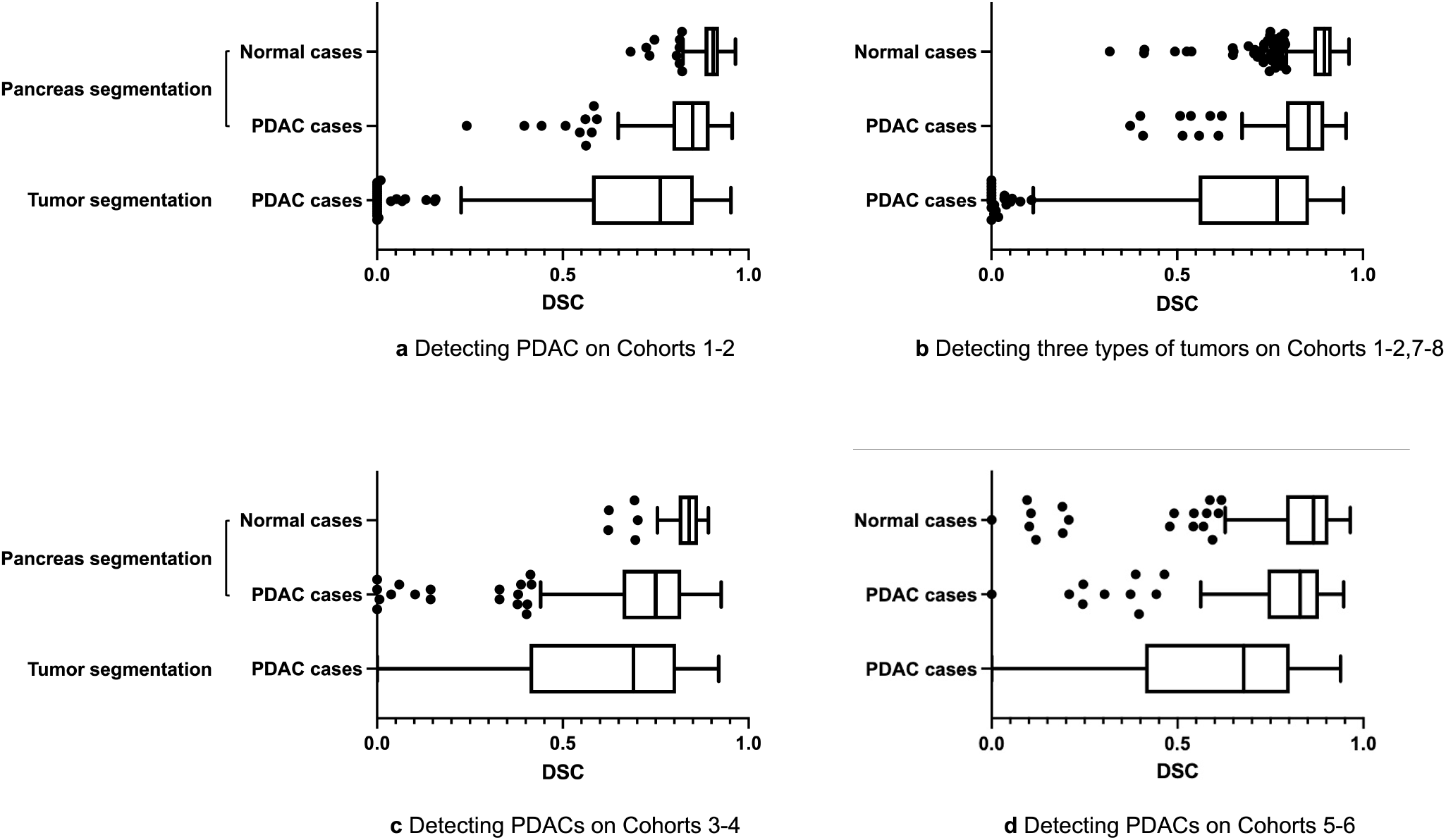
Performance of the pancreas segmentation and tumor localization evaluated by Dice-Sorenson similarity coefficient (DSC). Observe that the DSC scores are typically high but with some small outlier cases indicated by the black dots.

The CT scans from Cohort 4 were obtained from 82 healthy individuals without pancreatic disease, with images taken at the NIH. The images were acquired on Philips as well as Siemens scanners and the slice thicknesses (1.0 to 5.0mm) were considerably larger than those (0.5mm) from the healthy individuals in Cohort 1. Nevertheless, the DSC for the normal pancreas (83%, IQR 81% to 86%) were nearly as high as those obtained for the test set in Cohort 1 (87%, IQR 85% to 91%), as shown in Figure 5c.

The CT scans from Cohorts 5-6 were obtained from 164 individuals without pancreatic disease and 78 with PDAC (Table 1). The vast majority of these were acquired with Siemens scanners. In 77 scans, subjects were rotated along the vertical axis from 30 to 60 degrees (examples in Figure 3). Sensitivity and specificity were >90%, when either single-phase venous images or dual-phase images, were available. The DSC for the normal pancreas (84%, IQR 83% to 89%) were nearly as high as those obtained for the test set in Cohorts 1-2 (Figure 5d).

### 2.4 Task IV: Recognizing other pancreatic tumor types

Though PDACs are the most dangerous form of pancreatic tumors, they comprise a minority of those occurring in the pancreas. Intraductal Papillary Mucinous Neoplasms (IPMN), a benign pancreatic cystic neoplasm with the potential of malignant transformation, are more than ten-fold as frequent and the only radiologically detectable tumor entity more common than PDAC. Malignant neoplasms named Pancreatic Neuroendocrine Tumors (PanNETs) occur*∼* five-fold less frequently than PDACs, but can often be cured. Detection of these lesions is an important component of any approach designed to evaluate abdominal CT scans.

Detecting pancreatic cysts and PanNETs raises additional challenges for AI algorithms because these lesions exhibit a greater variety of texture patterns than PDACs. But we were able to train FELIX to recognize them with only a few modifications to those described above for detecting PDACs (modified algorithm suite named FELIX 1.3, §4). One of the most important of these modifications was multiscale processing, which proved critical for recognizing smaller lesions (see Figure 9).

The algorithmic development for FELIX 1.3 was done similarly to that for the other algorithms, with training and testing sets kept independent. When tested on healthy individuals in Cohort 1 and patients with PDACs in Cohort 2, its sensitivity and specificity remained as high as it was with FELIX 1.1, as expected. We then assembled a set of CT images from 450 patients with PanNETs and 458 patients with pancreatic cysts (Cohorts 7 and 8, respectively). The sensitivities for recognizing pancreatic cysts and PanNETs were 95% and 94%, respectively. The specificity of detecting three types of tumors was 95% (Figure 4b). As with PDAC, we defined a true positive as a lesion within the pancreas that was not only detected but correctly localized. The localization of these tumors was similar to that obtained with PDAC—a DSC of 57% (IQR 25% to 86%) for PanNETs and 66% (IQR 52% to 88%) for pancreatic cysts (Figure 5b).

The pancreatic cysts within Cohort 8 also provided an opportunity to assess the performance of the FELIX algorithms for detecting small lesions. PDACs are generally rather large when diagnosed, which is one of the major issues confronting their effective treatment. Because our study was retrospective in nature, the vast majority of the PDACs in Cohorts 2, 4, and 5 were larger than 2cm, though we were able to detect and localize PDACs smaller than 2cm with 77% sensitivity at a specificity of 88%. Pancreatic cysts are often detected adventitiously in abdominal CT scans carried out for other purposes, and many of them were <2cm in diameter. The sensitivity of FELIX for detecting pancreatic cysts <2cm was 76% at a specificity of 88%, with cysts as small as 2mm in diameter detectable (Figure 9). A cyst of 2mm in diameter is represented by only 30 voxels out of the 131,072,000 voxels in a typical CT image.

## 3 Discussion

The results summarized in Figures 4–5 show that pancreatic tumors, and in particular PDAC, can be detected and localized with FELIX algorithms at sensitivity and specificity >90%. When tested on Cohorts 1 and 2, from Johns Hopkins Hospital, the sensitivity and specificity were >95%. Algorithms were able to evaluate CT scans generated through a variety of protocols, with varying resolutions, slice thicknesses, radiographic protocols, and scanning instruments. The scale of these studies and the clinical performance of the FELIX algorithms substantially exceed those of previous studies. We anticipate that better performance can be achieved in future work by training on even larger datasets and by exploiting technical advances in AI algorithms.

But the FELIX study has several limitations. We certainly have not “solved” the domain transfer problem for pancreatic tumors. Though FELIX performed fairly similarly regardless of the source of the CT scan and the radiographic procedures used, it performed highest on scans from Johns Hopkins Hospital. Moreover, there are a large number of variables that can affect this performance that have not yet been tested. These include images obtained with instruments other than those we have tested on (predominantly manufactured by Siemens), those taken after oral contrast agents are administered, and those taken when there are extraneous features, such as clips or stents, within the patient. These extraneous features are easy to recognize by humans, but not by computers, unless they are represented in the training set.

A second limitation is in the detection of very small tumors. Optimally, an AI-based method would be able to detect PDACs as small as 5mm in diameter, as the earlier the detection the greater the chance for effective therapy. Moreover, small tumors are more likely to be missed by practicing radiologists. But the number of patients with PDACs that are detected when their tumors are <1cm in diameter is small, even in relatively large pancreatic cancer centers such as at Johns Hopkins or Heidelberg. It will require a large, multi-institutional collaborative study to acquire a sufficient number of small PDACs to engender cohorts for adequate training and testing of very small PDACs.

Third, though FELIX algorithms can detect pancreatic cysts and PanNETs in addition to PDACs, there are other pancreatic diseases, such as acute or chronic pancreatitis and metastatic lesions from other organs to the pancreas, that have not yet been evaluated.

Finally, our study was retrospective in nature, with diagnoses all previously made and confirmed through histopathological analysis. The eventual goal of FELIX is to be able to act as a “second reader”, providing the radiologist with a simple and instantaneously available tool to call attention to pancreatic lesions of interest. The next generation of FELIX will develop better AI algorithms, incorporate both radiologic and clinical features to predict the existence, size, boundaries, and type of lesion within the pancreas. This will enable the AI algorithms to be tested in a large, prospective study and to evaluate its clinical utility.

## 4 Methods

### 4.1 Study participants and sampling procedures

Table 1 summarizes the datasets used in this study. The distribution of tumor size in each dataset is presented in Figure 6. The attached supporting file contains the detailed demographic information.

**Figure 6.**
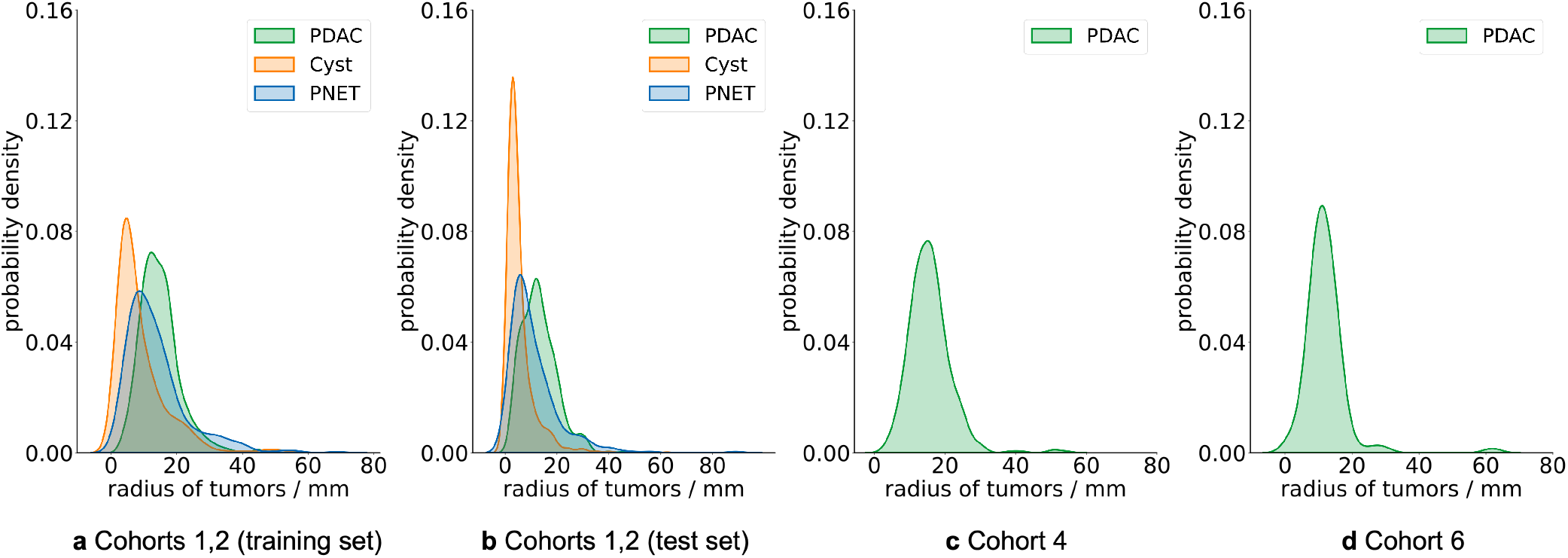
Tumor size distributions in Cohorts 1–2 (training set), Cohorts 1–2 (test set), Cohort 4, and Cohort 6.

*Cohorts 1, 2, 7, and 8* consisted of 2,519 subject cases, containing cases of Normal, PDAC, Cyst, and PanNET, respectively. This was a retrospective study approved by Johns Hopkins Hospital institutional review board. Pancreatic protocol CTs were retrospectively identified from clinical, pathological and radiological databases compiled between 2003 and 2020. Total 1,982 patients with pathologically proven 686 PDAC and 286 PNET were retrospectively collected from Radiology and Pathology databases. Patients with pancreatic tumors between 2011 to 2020 were scanned with a dual-source MDCT scanner (Somatom Definition, Definition Flash, or Force, Siemens Healthineers) and the patients between 2003 to 2010 were scanned on a 16- or 64-slice MDCT scanner (Somatom Sensation 16 or 64, Siemens Healthineers). 799 renal donors without pancreatic tumors were considered to be normal controls for classification purposes. Patients and renal donors were injected with 100-120 mL of iohexol (Omnipaque, GE Healthcare) at an injection rate of 4-5 mL/sec. Scan protocols were customized for each patient to minimize dose but were on the order of 120 kVp, effective mAs of 300, pitch of 0.6-0.8. The collimation was 128×0.6 mm or 192×0.6 mm for the dual source scanner, 16×0.75 mm for the 16-slice scanner, and 64×0.6 mm for the 64-slice scanner. Most (99%) of these renal donor cases were collected prior to 2010 so as to ensure that they did not develop pancreatic disease following their scans. Arterial phase imaging was performed with fixed delay or bolus triggering, usually between 25-30 seconds post-injection, and venous phase imaging was performed at 60 seconds. Both of arterial and venous phase images were collected for patients with pancreatic tumors and renal donors, so there were 5,038 annotated scans in total. We randomly split the 5,038 scans into 3,192 and 1,846 scans for training and testing. Each CT scan consists of 319*∼*1,051 slices of 512*×*512 pixels, and have a voxel spatial resolution of ([0.523*∼*0.977]*×* [0.523*∼*0.977]*×*0.5)mm^3^. We split the union of the four Cohorts into training and test sets. The training set contains a total of 3,192 CT scans (560*×*2 PDACs, 205*×*2 Cysts, 300*×*2 PanNETs and 531*×*2 Normals. For the 1,846 (i.e., 923*×*2) testing set, it contains 215*×*2 PDACs, 253*×*2 Cysts, 150*×*2 PanNETs and 305*×*2 Normals.

*Cohort 3* consisted of 246 subjects with 399 abnormal CT scans (pathologically confirmed PDACs were included). Slice thickness ranges from 1*∼* 5mm. The outside CT scans were performed at various institutions across the United States and were acquired with GE (39%), Siemens (38%), Phillips (12%), and Toshiba (11%) scanners. The scan protocol varied based on local institutional preferences. External validation was performed on CT datasets obtained at various institutions in the United States that were uploaded in the Johns Hopkins Hospital picture archiving and communication system (PACS) for second opinion interpretation.

*Cohort 4* consisted of 82 abdominal contrast enhanced venous phase CT scans. The scans had resolutions of 512*×*512 pixels with varying pixel sizes and slice thicknesses between 1.5 *∼* 2.5mm, and were acquired with Philips and Siemens MDCT scanners during portal venous phase. The National Institutes of Health Clinical Center performed 82 abdominal contrast enhanced 3D CT scans (*∼* 70 seconds after intravenous contrast injection in portal-venous) from 53 male and 27 female subjects. Seventeen of the subjects are healthy kidney donors scanned prior to nephrectomy. The remaining 65 patients were selected by a radiologist from patients who neither had major abdominal pathologies nor pancreatic cancer lesions.

*Cohorts 5, 6* consisted of 242 dual phase CT scans collect at Heidelberg for external validation. 78 cases with surgically resected PDAC were retrospectively identified from the surgery and radiology database between 2011 and 2020. 164 cases (matched for age and gender) who underwent CT scans for trauma evaluation were used as controls. The CT scans of these patients were reviewed to ensure that there no pancreatic abnormality and no significant abdominal trauma. The CT scans with PDACs and normal controls were acquired with dual-source MDCT scanner (Somatom Definition, Definition Flash, Siemens Healthineers) or 64-slice MDCT scanner (Somatom Sensation 64, Siemens Healthineers). Arterial and venous phase CT imaging was performed for patients with PDAC and normal controls. CT scans had resolutions of 512*×*512 pixels with varying pixel sizes (0.57*∼* 0.97mm), and slice thickness between 0.64*∼* 2.0mm. Most scans included the whole upper body of the patient in addition to the abdomen. In 77 cases, subjects were rotated along the vertical axis, with a degree ranges from 30 to 60. In the pre-processing stage of FELIX 1.2, arterial scans were aligned to venous scans with isometric transformations, so that the rotations in the venous phase were kept.

### 4.2 Establishment of Ground-truth by manual annotation

The whole three-dimensional volumes of pancreas and tumors were manually segmented by five trained annotators using commercial segmentation software. For the subjects with dual-phase CT images, pancreas and pancreatic tumors were separately annotated in both arterial and venous phases by one of the five annotators. The boundaries and tumor locations of each subject were then verified by one of three additional experienced radiologists, none of whom performed the annotations.

System and human errors can affect the training and evaluation of machine learning algorithms. Therefore, data cleaning, corrections of errors after the initial data is obtained, was an important step. Possible human mistakes and intra-/inter-observer variations were first visually checked for by human experts. Errors or major inconsistencies by missing annotation of a slice or a part of organ with region of interest (ROI) were then doubly-checked by our in-house software. ROI information, in which the annotated target abdominal structures were recorded, were computed by the software and used for training and testing. Radiologist re-review, see Appendix B, was used to correct for errors in the ground truth which can occur, for example, if a small tumor was not annotated or if its annotated location was slightly incorrect.

### 4.3 Algorithm Development

Our goal is to detect the pancreas and three types of tumors from unaligned venous and arterial CT scans. We address this goal using deep networks trained for semantic segmentation^13^. We used the U-Net architecture as the basic segmentation method. This consists of a shared Siamese encoder for encoding images to features and a decoder for projecting features to predictions. The input of the network can be either dual-phase scans or single-phase (venous or arterial) scan, and the output is the segmentation prediction. For dual phase, to include information from both venous and arterial scans, we designed an auto-alignment module that can register and align the two phases. This auto-alignment module is inserted at the end of different encoder blocks. It contains a variety of alignment operation such as summation, concatenation, spatial transform, and cross attention (illustrated in Figure 7). We used neural architecture search over the set of alignment operations to optimize performance. Postprocessing was applied after the networks to decrease the number of false positives as a result of the prediction of lesions outside the pancreas by the algorithms.

**Figure 7.**
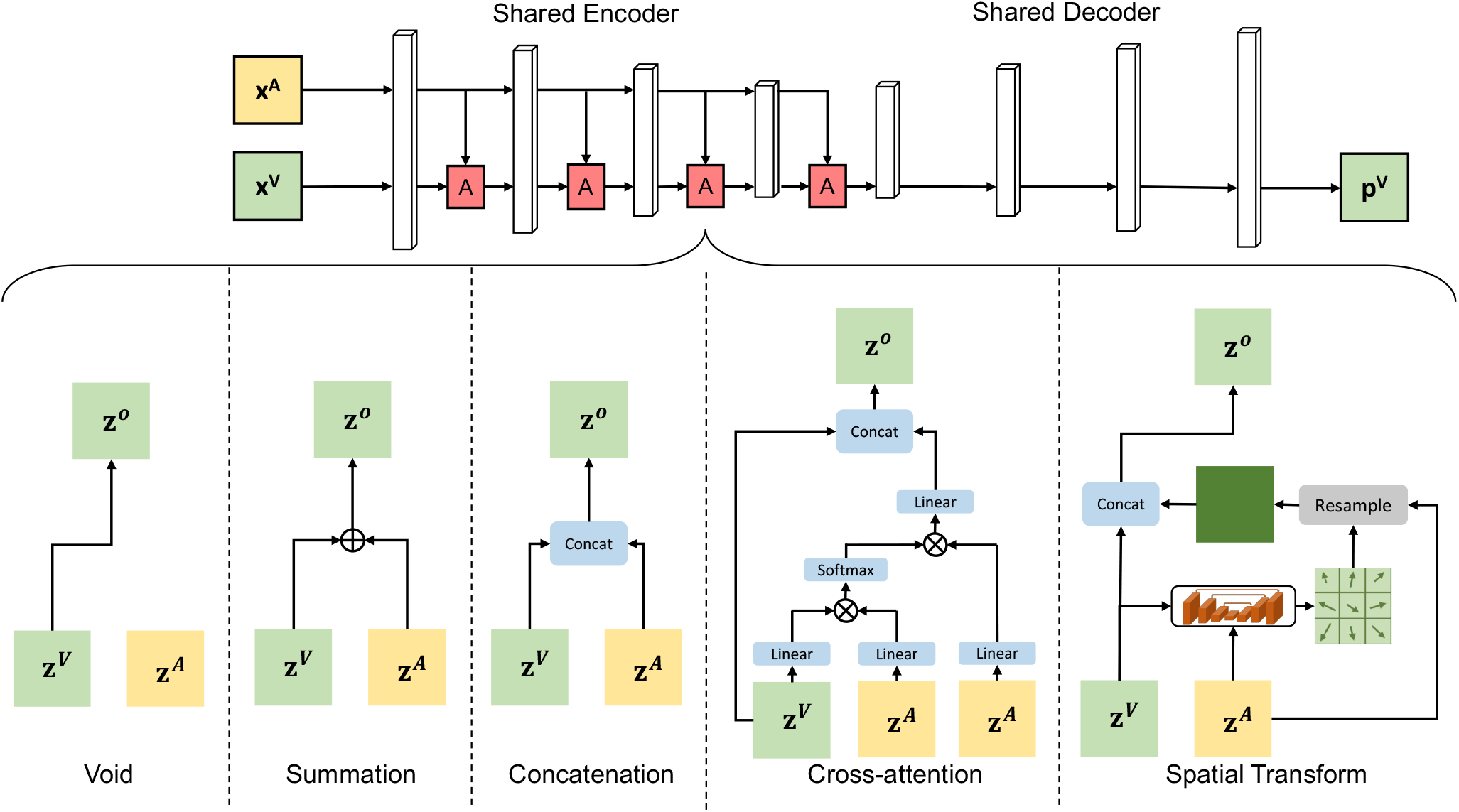
An illustration of FELIX 1.0 and the proposed auto-alignment for dual-phase scans. During the search process, an auto-alignment module is inserted after every encoder block to perform a dual-phase feature alignment, where the alignment operations can be chosen from the following: void (no alignment), summation, concatenation, cross-attention, and spatial transform. We note that this search space includes both alignment location and operation.

#### FELIX 1.0

FELIX 1.0 can process either dual-phase or single-phase scans. The alternative versions (FELIX 1.1-1.3) are the extensions of FELIX 1.0 for different tasks.

##### Single-phase algorithm

We used 3D U-Net^19,20^, which is a symmetric architecture consisting of encoder and decoder sub-networks. The encoder sub-network took the input image and reduced the spatial resolution in successive layers while increasing the channels; the decoder sub-network increased the spatial resolution while reducing the channels. Four residual blocks were used between poolings in the encoder and bilinear interpolations in the decoder. In the end, a 1*×*1*×*1 convolution was used to map the channels to the desired number of classes, e.g., background, pancreas, PDAC, Cyst, PanNET, etc. Skip connections were used between the encoder and decoder sub-networks to recover fine-grained details of the target objects, allowing U-Net to segment fine-grained structures such as small tumors.

##### Dual-phase algorithm

Following previous studies^21,22^, the dual-phase algorithm used arterial to help venous prediction. Unlike single-phase algorithm, the U-Net structure for dual phase consisted of a shared Siamese encoder to encode images to features and a decoder to project features to predictions. The input of the dual-phase algorithm is a pair of venous and arterial scans, and the output is the segmentation prediction of the venous scan. To incorporate information from both venous and arterial scans, we design an auto-alignment module that can determine the operation of dual-phase alignment. The possible alignment operation includes void (no alignment), summation, concatenation, cross attention, and spatial transform. The auto-alignment module is inserted at the end of different encoder blocks. Instead of using a hand-designed architecture, we learn the architecture by Neural Architecture Search (NAS)^23^ (illustrated in Figure 7). Formally, the entire dataset is denoted as 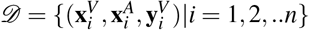 where *n* is the total number of subjects, 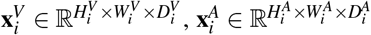 are venous and arterial CT scans of the *i*-th subject, and 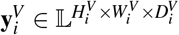 is the voxel-wise annotated label in the venous scan. Here, 𝕃 = {0, 1, 2, 3} represents our segmentation targets, i.e., background, healthy pancreas tissue, pancreatic duct (crucial for PDAC clinical diagnoses), and PDAC mass. Our goal is to find a mapping function *ℱ* whose inputs and outputs are a pair of two-phase scans **x**^*V*^, **x**^*A*^ and segmentation results **p**^*V*^, respectively, i.e., **p**^*V*^ = *ℱ* (**x**^*V*^, **x**^*A*^). We denote the encoded features of the arterial and venous scans at a certain level by **z**^*V*^ and **z**^*A*^. An alignment operation aims to align and fuse the dual-phase features. We denote by **z**^*O*^ the output feature map after a certain alignment operation. The following operations are considered for alignment: *(1) Void:* The venous and arterial features do not align with each other: **z**^*O*^ = **z**^*V*^. *(2) Summation:* The output features are the element-wise summation of venous and arterial features: **z**^*O*^ = **z**^*V*^ + **z**^*A*^. *(3) Concatenation:* The output features are the concatenation of venous and arterial features along the channel dimension: **z**^*O*^ = **z**^*V*^*⊕* **z**^*A*^, where *⊕* denotes the concatenation operation of the two vectors. *(4) Cross-attention:* We consider two-phase collaboration in a non-local attention manner, which can globally encode each location in the venous features by receiving information from the entire arterial features. Conceptually, **z**^*O*^ = **z**^*V*^ *⊕* (*so f tmax*(**z**^*V*^ **z**^*A*^)**z**^*A*^) *(5) Spatial transform:* Spatial transform^24^ was widely adopted in the task of registration between two images. We consider it as an operation which can handle the large offsets between the venous and arterial scans. The spatial transform was applied to the arterial scan only. Specifically, we use a light-weighted U-Net to first estimate a deformation field *φ* of the arterial feature map **z**^*A*^ to the venous feature map **z**^*V*^. Afterwards, we fuse the deformed arterial feature map to the venous feature map by concatenation. This process can be formulated as follows: **z**^*O*^ = **z**^*V*^ *⊕* (*φº* **z**^*A*^), where *⊕* and º denote the concatenation of two tensors and the element-wise deformation operations on a tensor, respectively.

#### FELIX 1.1

In addition to pancreas segmentation, FELIX 1.1 was capable of detecting and segmenting PDACs from either single-phase or dual-phase scans. This involved two stages: pancreas detection and tumor segmentation. In the first stage, we used FELIX 1.0 to detect the rough location of the pancreas from the whole CT scan and place a bounding box that surrounds the pancreas. The first stage could 100% accurately localize the pancreas, with a DSC score of 87% and 86% for dual-phase and single-phase algorithms, respectively. The second stage took the cropped CT sub-volume as input (in the center of the bounding box of the pancreas) and used a U-Net to segment the pancreas into normal voxels and voxels that belong to PDACs. The DSC of PDAC localization obtained by FELIX 1.1 averaged 65% and the DSC for the venous or arterial phase along averaged 63% on the test set (Figure 5a).

#### FELIX 1.2

To enable the algorithms to generalize to data from other institutions, we created a much bigger training dataset by applying data augmentation techniques to the JHH data, including 3D rotations of the CT scans and adjusting other scan properties such as slice thickness (normalized to 30mm). The increased variety of training data enabled us to train a much larger deep network, created by adding a few extra components to our original network, which was able to exploit the extra training data without overfitting. The single-phase algorithm was used for external data such as Cohort 3 and Cohort 4 because only venous-phase scans were provided.

#### FELIX 1.3

This algorithm aims at detecting and recognizing two other tumor types (pancreatic cysts and PanNETs) in addition to FELIX 1.1 that detects PDACs. Cysts and PanNETs exhibit varying texture patterns and tumor sizes (Figure 6). To improve the detection and localization of very small tumors, we applied standard multi-scale techniques that processed the CT scans at different levels of resolution and then combined the results. we train a multi-scale algorithm on JHH data and evaluate it on Cohorts 7-8 with five scales (1.0, 1.25, 1.5, 1.75, 2.0), then we further merge those results from different scales. Without multi-scale training, our dual-phase algorithm can obtain 83.4% sensitivity of detecting small tumors. The multiscale training strategy greatly improves performance, achieving an overall sensitivity of 88.7% (+5.3) before radiologist re-review and 89.3% (+5.9) after radiologist re-review for small tumors, while the specificity remains competitive (88.2%) to base algorithms. The smallest lesion we detected was 2mm radius. Performance of pancreatic tumor detection stratified by tumor size is presented in Figure 8, and examples of small tumor detection are illustrated in Figure 9.

**Figure 8.**
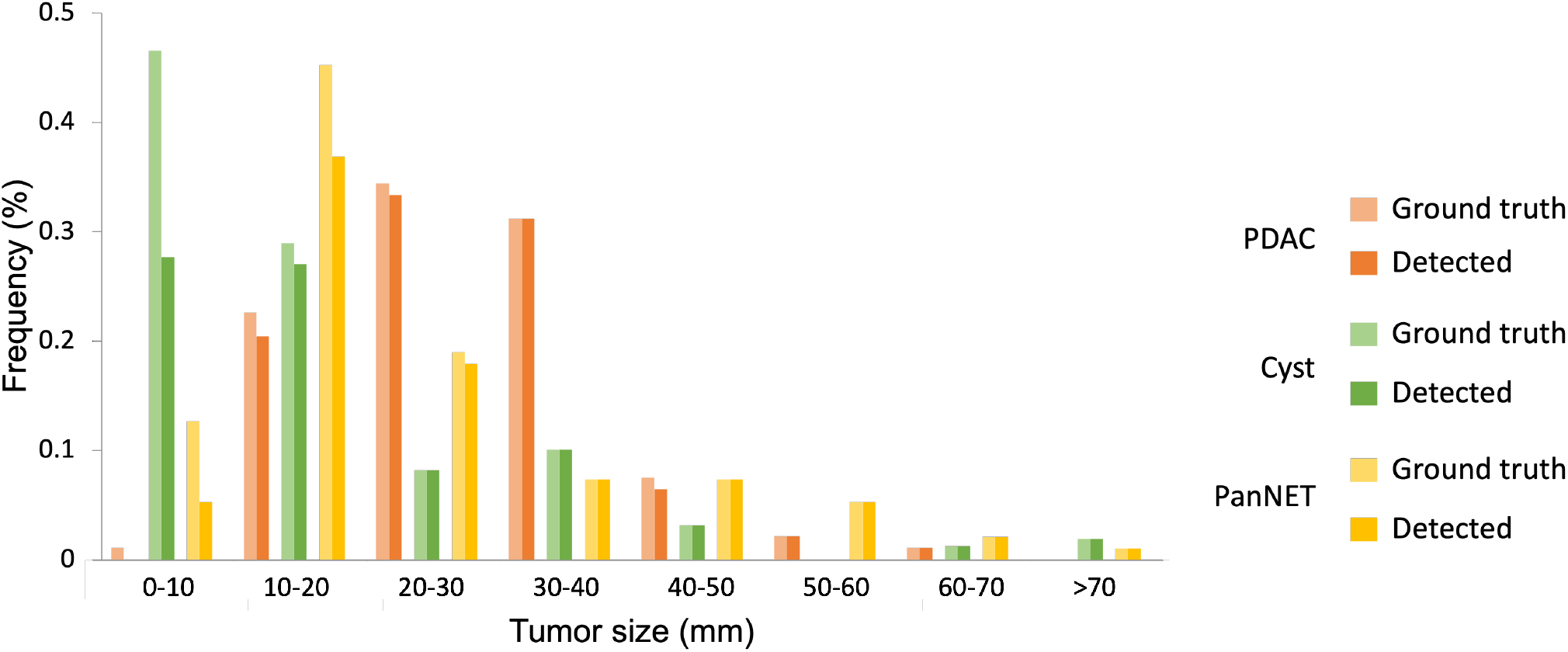
Performance of pancreatic tumor detection stratified by tumor size. The smallest tumor we detected was 2mm. Our false negatives are mostly smaller than 20mm, frequently smaller than 10mm. We increase sensitivity to small tumors by multi-scale training.

**Figure 9.**
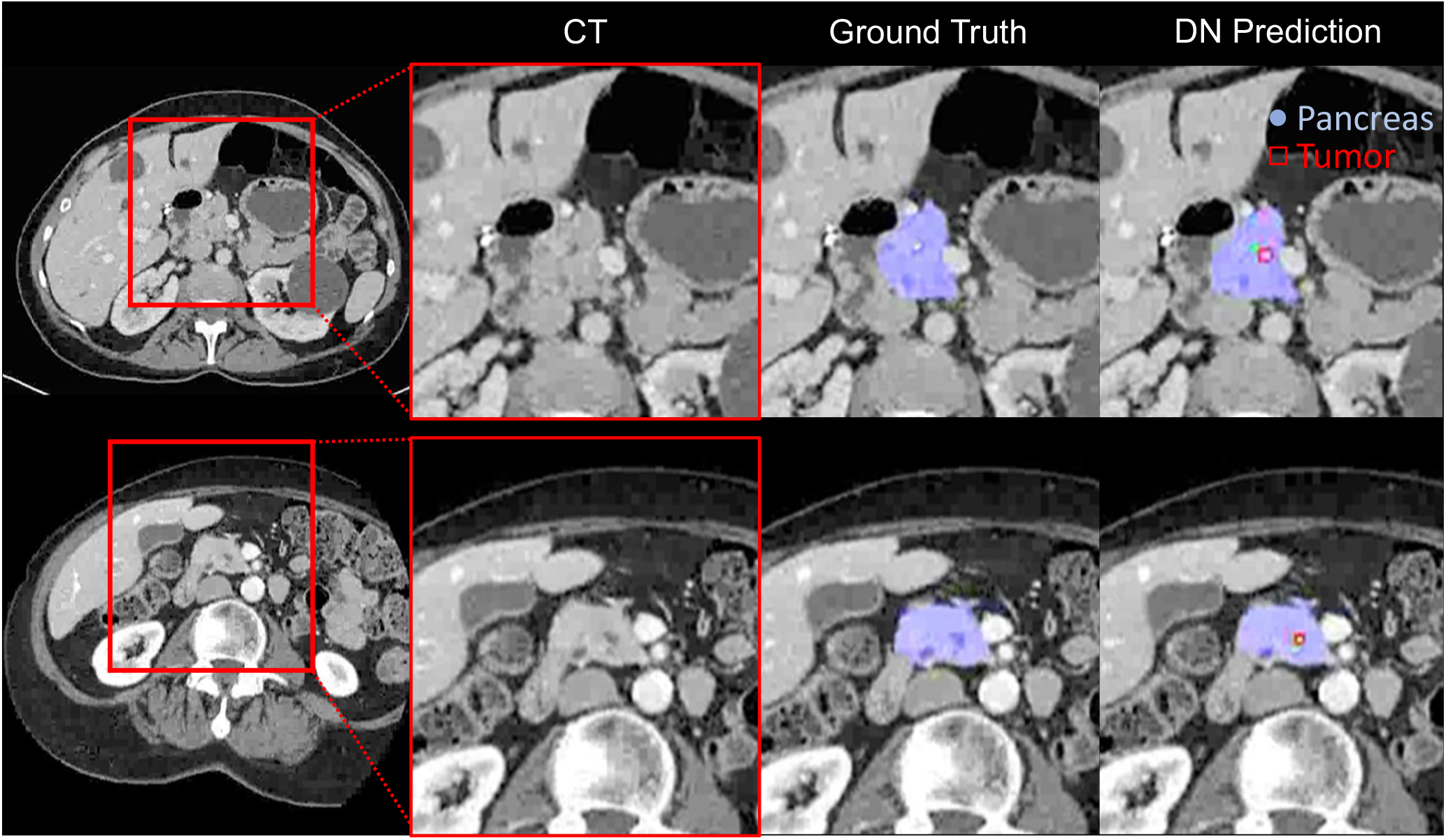
Our multiscale algorithm can detect Cysts that were not annotated by radiologists. These predictions were verified to be correct through radiologist re-review. Top: Cyst of 2mm radius. Bottom: Cyst of 4mm radius.

#### Post-processing

The post-processing stage is to eliminate most false positives using a variety of cues, such as prediction size, distance from the pancreas, and several handcrafted features. These cues are usually not fully exploited by deep learning algorithms. First, for PDAC detection, we discard the predicted components with less than 500 voxels; for Cyst and PanNET detection, we discard the predicted components with less than 30 voxels. Second, we dismissed the predictions if the surface of the predicted tumor is not attached with the surface of the predicted pancreas. Third, handcrafted features were extracted from four different perspectives, i.e., uncertainty, quality assessment, shape, and geometry. We used a two-way cross-validation on the validation set for hyper-parameters tuning to compute these imaging features. A sequential feature selection^25^ was then conducted on the hybrid feature pool. Specifically, starting from an empty set, we picked one feature at a time from the remaining feature pool that minimized a validation loss. Consequently, we adopted VAE, sphericity, and surface volume ratio for PDAC detection, uncertainty, VAE, and sphericity for Cyst and PanNET detection. A predicted component was considered as positive only if all these imaging features agree it is positive.

##### (1) Uncertainty

We hypothesize that segmentation with bad quality is more likely to be a false positive. Inspire by^26^, we used an entropy-based uncertainty to assess the quality of segmentation and distinguish between false positives and true positives. We calculate the uncertainty in a way by accumulating the entropy on the voxel that is predicted as lesion in **p**^*V*^. Specifically, we have

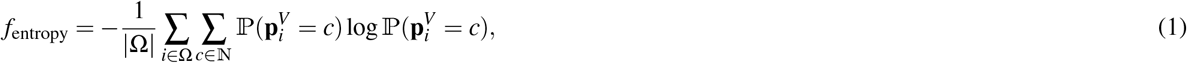

where 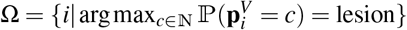.

##### (2) VAE

A variational autoencoder (VAE) is learned to reconstruct the ground truth and then the reconstruction error is used to evaluate the segmentation quality. The quality assessment feature is usually targeted at anomaly detection^27^. In false positive reduction, we treat the properties within tumor region as target distribution so that the false positives, which do not correspond to tumor region become anomalies. Shape and texture can represent orthogonal properties of pancreatic lesions so that they provide complementary cues when combined together. Specifically,

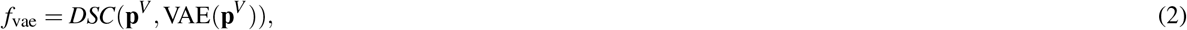

where DSC(*·*) is the function to calculate dice coefficient, formulated in Equation 6.

##### (3) Surface volume ratio

We adopted the ratio between surface and volume of a predicted component to reject false positives by analyzing shape features. A lower ratio indicates a more compact (sphere-like) shape.

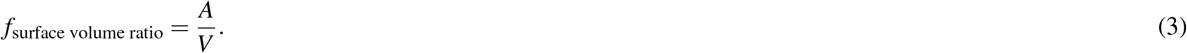

Surface area (*A*) is obtained by taking the number of all voxels that belong to the edges of a predicted component. Mesh volume (*V*) is the total number of voxels in a predicted component.

##### (4) Sphericity

Sphericity is the ratio of the surface area of a sphere to the surface area of the particle^28^. Sphericity measures the roundness of the shape of the tumor region relative to a sphere. It has a value in the range of [0, 1], where a value of 1 indicates a perfect sphere.

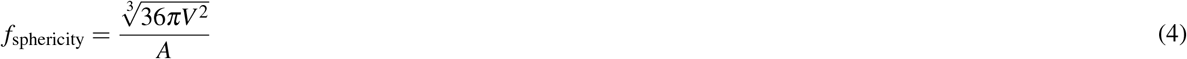

### 4.4 Algorithm Evaluation

For classification of PDAC and non-PDAC cases, we report sensitivity (also known as true-positive rate) and specificity (as known as true-negative rate), defined as:

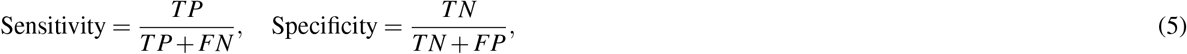

where TP, TN, FP, FN denote the number of true positives, true negatives, false positives, false negatives, respectively. Pie plots of sensitivity and specificity were presented in Figure 4.

We report two metrics, DSC (Dice similarity coefficient) and ASSD (average symmetric surface distance), to measure the segmentation performance. Box plots of these two measures were presented in Figure 5 and Figure 10, respectively. The DSC score is commonly used as an evaluation metric and takes a value of 0 when both masks do not overlap at all and 1 for perfect overlap.

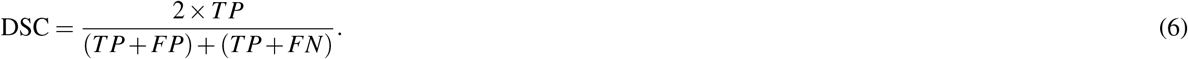

ASSD measures the average distance between the surface of the tumor/organ segmentation result to the nearest boundary voxels of the ground truth in 3D. It has a value in the range of [0, ∞]. They are used to measure the area similarity and the boundary or shape similarity, respectively. A better segmentation algorithm produces a larger value of DSC while a smaller value of ASSD.

**Figure 10.**
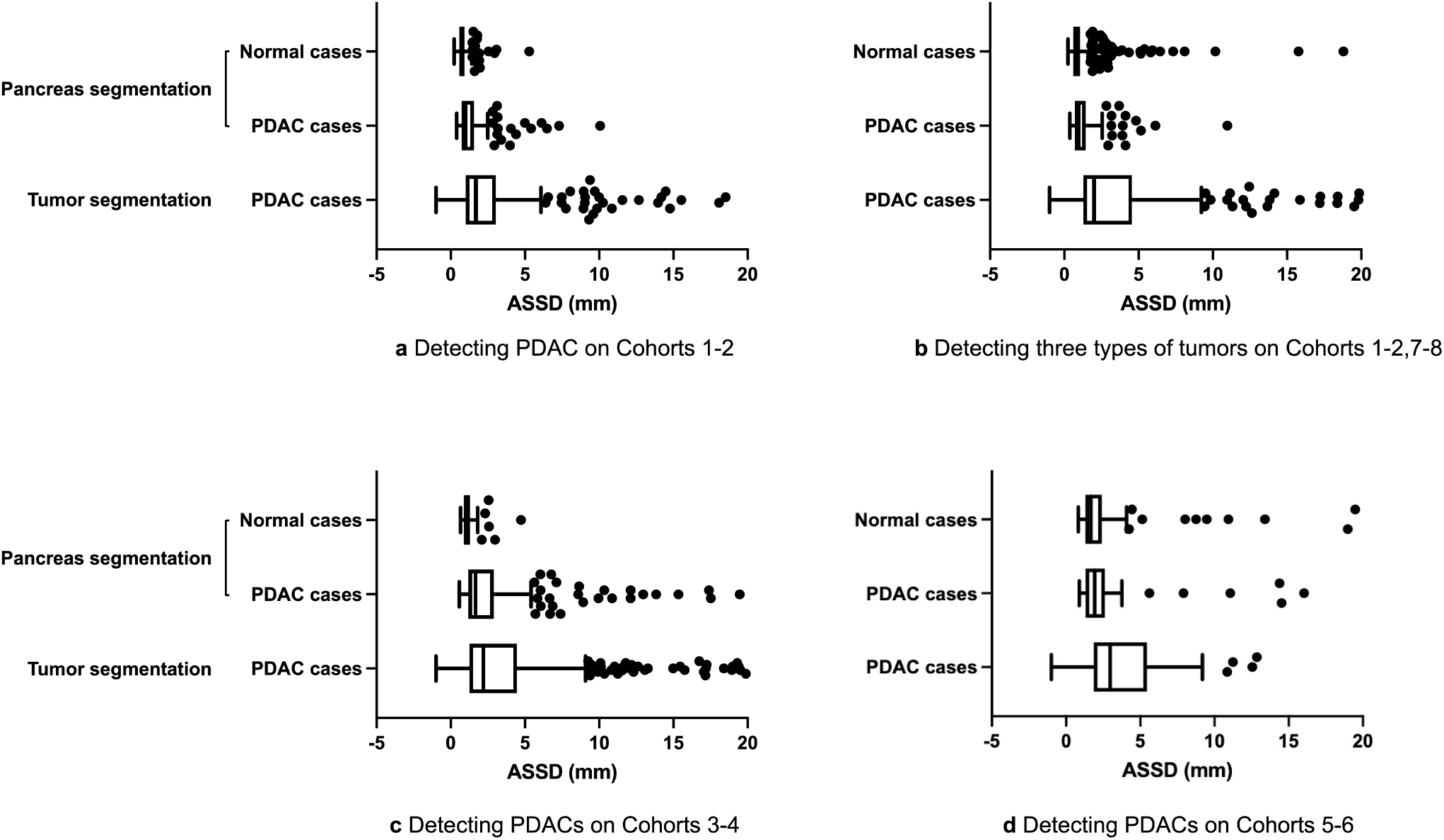
Quantitative performance of pancreas segmentation and tumor localization, evaluated by average symmetric surface distance (ASSD).

## Data Availability

All data produced in the present study are available upon reasonable request to the authors.

## Data availability

Researchers who are interested in our work can request access to the de-identified raw images. Use of data is limited to research purposes and redistribution of data is not allowed. The request should be made to the corresponding authors. Subject to the institutional review boards’ ethical approval, deidentified data have been attached in the supporting file. All experiments and implementation details are described thoroughly in the Methods section so they can be independently replicated with nonproprietary libraries.

## Code availability

The code used to train and evaluate the model performance would be openly available at Github.

## Author contributions

Y.X., Q.Y., L.C., and S.K. contributed equally to the manuscript. A.L.Y., and E.K.F. presented the conception of the work; K.K., R.H.H, B.V., A.L.Y., and E.K.F. obtained funding to support the work and built up the research team for joint development; A.L.Y. and B.V. drafted the manuscript with assistance from Zo.Z, S.P., and R.H.H; Zo.Z, Y.X., Q.Y., F.L., Zh.Z., and Y.L. drafted the supplementary materials. L.C., S.K., B.K., M.K., P.M., and E.K.F. acquired the data; L.C., S.K., D.F.F, S.S, S.J.F, A.B., and E.S.Z. annotated the images for further analysis; A.L.Y., S.P., Zo.Z., Y.L., Y.W., W.S., L.X., C.W., and A.J. designed the study; Y.X., Q.Y., F.L., J.C., Zh.Z., B.L., Y.Z., and S.P. developed the deep networks; S.P. created the new software for visualization. All the authors made substantial contributions to this work and have critically reviewed the manuscript before submission.

## A Background

### Public pancreas CT datasets

There are several publicly available datasets for pancreas detection/segmentation and tumor detection, such as the Medical Segmentation Decathlon (MSD) dataset^6^, the TCIA-PDA dataset^29^, and the National Institutes of Health Pancreas CT (NIH-Pancreas) dataset^30^ The MSD pancreas dataset consists of 420 abdomen CTs of subjects with pancreatic lesions (e.g., intraductal mucinous neoplasms, pancreatic neuroendocrine tumors, or pancreatic ductal adenocarcinoma) from the Memorial Sloan Kettering Cancer Center. The dataset has been split into two groups: a training subset (*n* = 281) and a testing subset (*n* = 139). Only the training subset has voxel-wise pancreas and tumor annotation. All the studies are contrast-enhanced scans acquired in the venous phase. The TCIA-PDA dataset consists of 6 MRIs and 60 CTs of subjects from the National Cancer Institute’s Clinical Proteomic Tumor Analysis Consortium Pancreatic Ductal Adenocarcinoma (CPTAC-PDA) cohort. Age, gender, tumor size, histologic type, and grade are available for all the subjects, but voxel-wise tumor or pancreas annotation is not available. 57 out of 60 CTs are in venous phase. The NIH-Pancreas dataset consists of 82 venous phase CTs performed at the NIH Clinical Center on 80 subjects. All CTs have a morphologically normal pancreas. The dataset provides voxel-wise annotation of pancreas segmentation for all subjects performed by manual slice-by-slice tracings of the pancreas. In addition, numerous abdominal CT datasets are publicly available with manual annotation of organ segmentation including the pancreas, but whether these CTs contain pancreatic tumors is unknown. For example, the Synapse dataset (from the MICCAI Multi-Atlas Labeling Beyond the Cranial Vault challenge)^31^ consists of 30 venous phase CT scans with manual annotation for segmentation of 13 abdominal organs; Abdominal-1K^32^ provides more than 1000 CT scans from 12 medical centers with liver, kidney, pancreas, and spleen annotated; WORD^4^ has 150 CT scans with 16 organs annotated; and most recently, TotalSegmentor^5^ releases 1204 CT scans with 104 anatomical structures annotated. Our curated JHH dataset is unprecedented in scale, consisting of over 2,500 dual-phase contrast-enhanced CT scans with full labels of 20 organs as well as exhaustive labels of cysts, ducts, and tumors in the pancreas.

**Figure 11.**
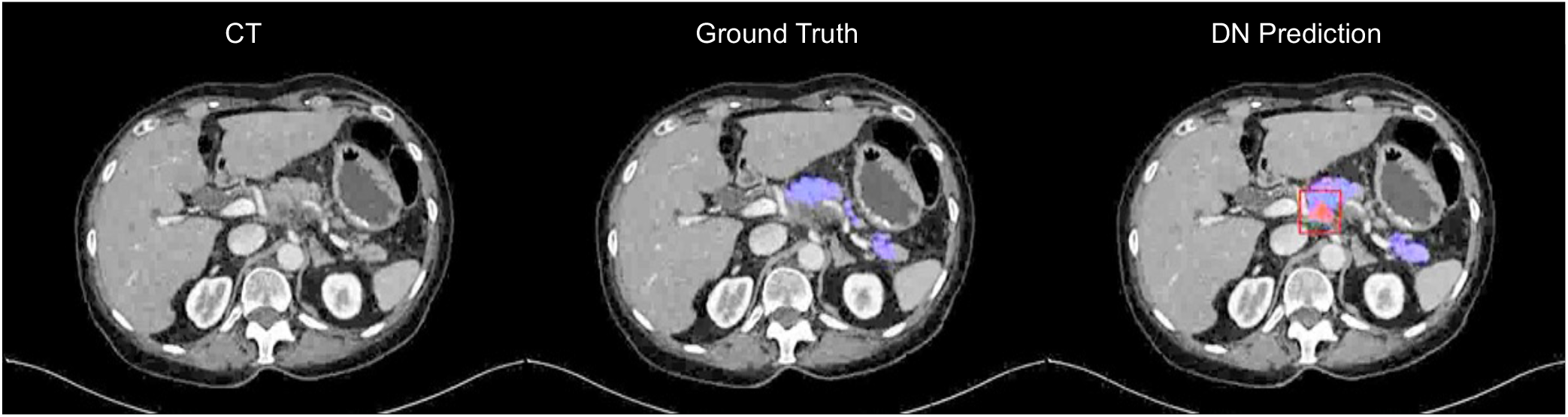
After radiologist re-review, we verified that the prediction framed in red was a true positive of PDAC but it was missed by annotator.

### AI for pancreas and pancreatic tumor detection

With the recent advances of deep learning, automated pancreas segmentation has achieved tremendous improvements^8–10,33–36^, which is an essential prerequisite for pancreatic tumor detection^37–41^. Meanwhile, researchers are pacing towards automated detection of pancreatic adenocarcinoma (PDAC), the most common type of pancreatic tumor (85%)^42^ and with the lowest 5-year survival rate among cancers^43^. Most existing works used venous-phase CT scans for detecting and segmenting pancreatic tumors^7,22^.^21^ developed a hyper-pairing network for PDAC segmentation from multi-phase CT scans to integrate information from both arterial and venous scans.^38^ proposed a framework to improve PDAC segmentation with multi-institutional and multi-phase, partially labeled data. They both used traditional image registration approaches^44,45^ for pre-alignment and then applied a deep network that took the phases as input. Unlike their methods, we particularly investigate how to register multiple phases in feature space with more complex fusion techniques, either in a manually designed or automated way. There are complimentary AI techniques that used texture features (in particular Radiomics features) of the pancreas, and then trained a random forest algorithms classifier algorithm^14,46^. These were able to classify if a pancreas contained a tumor, but were not suitable for localizing the tumor.

**Figure 12.**
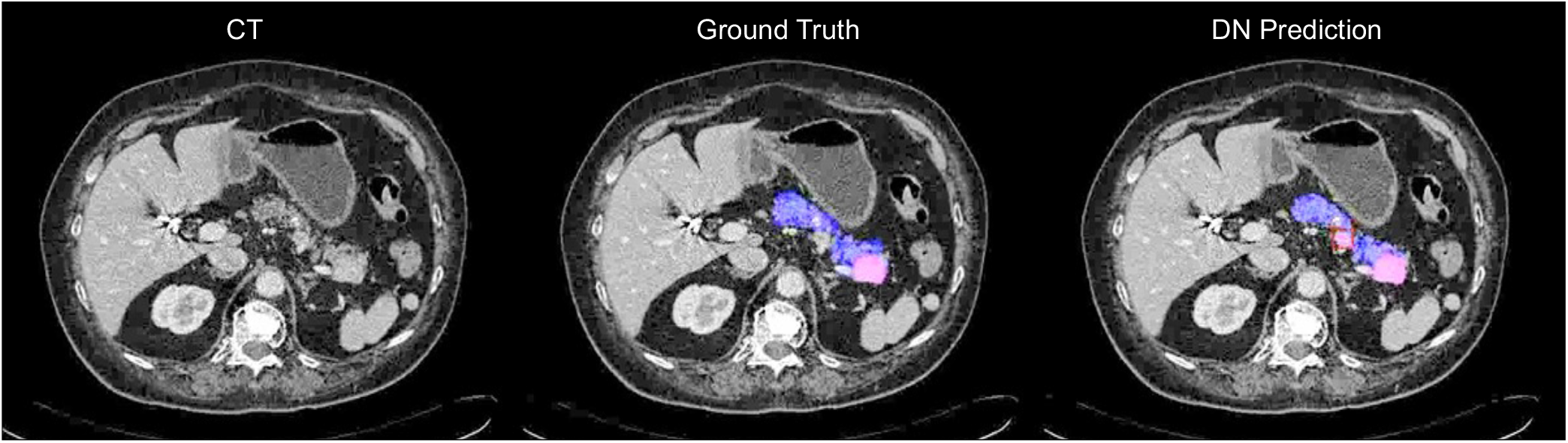
After radiologist re-review, we verified that the prediction framed in red was a true positive of PanNET but it was missed by annotator.

## B Radiologist Re-review

### Overview

After application of the algorithms to the cohorts in this study, radiologists re-reviewed all cases in which there was a discrepancy between the original radiologic annotation of the data and the prediction of the algorithm. In no case was the prediction of the algorithm changed on the basis of this re-review. However, of the 203 cases re-reviewed, the original radiologic annotation was found to be erroneous, and this annotation was accordingly changed in the datasets (Tables 2–3).

**Table 2.** Taxonomy of *false negatives* on the test set of Cohorts 1 and 2 using our dual-phase algorithm.

| Radiologist re-review | PDAC | Cyst | PanNET | Total |
| --- | --- | --- | --- | --- |
| TP (close to GT, AI better than GT) | 4 | 3 | 0 | 7 |
| Exclude (surgery, fluid, not-due-to-stent) | 0 | 1 | 0 | 1 |
| Incorrect annotation (need to be fixed) | 0 | 0 | 1 | 1 |
| Lymph nodes | 0 | 0 | 1 | 1 |
| Classified as duct | 0 | 6 | 0 | 6 |

**Table 3.** Taxonomy of *false positives* on the test set of Cohorts 1 and 2 using our dual-phase algorithm.

| Radiologist re-review | PDAC | Cyst | PanNET | Normal | Total |
| --- | --- | --- | --- | --- | --- |
| TP (correlate to abnormalities) | 15 | 1 | 2 | 0 | 18 |
| TP (close enough, AI predicts better) | 3 | 0 | 1 | 0 | 4 |
| TP (no label, cyst, serous, IPMN) | 10 | 34 | 10 | 3 | 57 |
| Exclude (surgery, fluid, serous, not-due-to-stent) | 3 | 1 | 3 | 0 | 7 |
| Duodenum | 0 | 1 | 0 | 0 | 1 |
| Veins/Vessels/Arteries | 3 | 14 | 4 | 5 | 26 |
| Pancreatic duct | 8 | 8 | 7 | 2 | 25 |
| CBD | 3 | 4 | 4 | 3 | 14 |
| SMV | 0 | 4 | 2 | 1 | 7 |
| Focal fat | 0 | 5 | 2 | 9 | 16 |
| Subtle texture change | 0 | 2 | 5 | 0 | 7 |
| Splenic artery | 0 | 0 | 2 | 3 | 5 |

Radiologist re-review of the false positives and false negatives showed that the false positives and false negatives of the algorithm were almost always understandable. The false positives mainly corresponded to small regions in the scan that an experienced radiologist would consider suspicious and worth inspecting more closely. By contrast, the false negatives were typically lesions that were also hard for experienced radiologists to detect. Radiologist re-review also enabled us to correct for errors in the ground truth which can occur because: (i) there is a small tumor in the scan which was not annotated, (ii) the tumor was annotated but its location was slightly incorrect (considering the difficulty of annotating the tumors the AI results can be more accurate than the ground truth), and (iii) an area was annotated as tumor, but on re-review no lesion was present. We report results both before and after the radiologist re-review. Some of the false negatives occurred when the AI algorithms predicted tumors very close to the annotations and hence direct radiologists to the rough location (and might, considering the difficulty of annotating tumors, be more accurate than the annotations).

### Recognizing a PDAC within the pancreas

FELIX 1.1 has sensitivity and specificity of 93.0% and 99.0%. We were able to localize the PDACs fairly accurately, obtaining DSC scores of 65.3%. After radiologist re-review, sensitivity and specificity improved to 96.6% and 99.0% (Figure 14a). Using the venous phase only, FELIX 1.0 gave a sensitivity of 92.5% and a specificity of 93.0% before radiologist re-review and 92.4% and 93.0% after radiologist re-review. We conclude that the AI algorithms trained and tested on the Hopkins dataset attain high sensitivity and specificity, similar to those of radiologists. The algorithms also accurately localize PDACs enabling radiologists to visually inspect specific locations in the scans.

**Figure 13.**
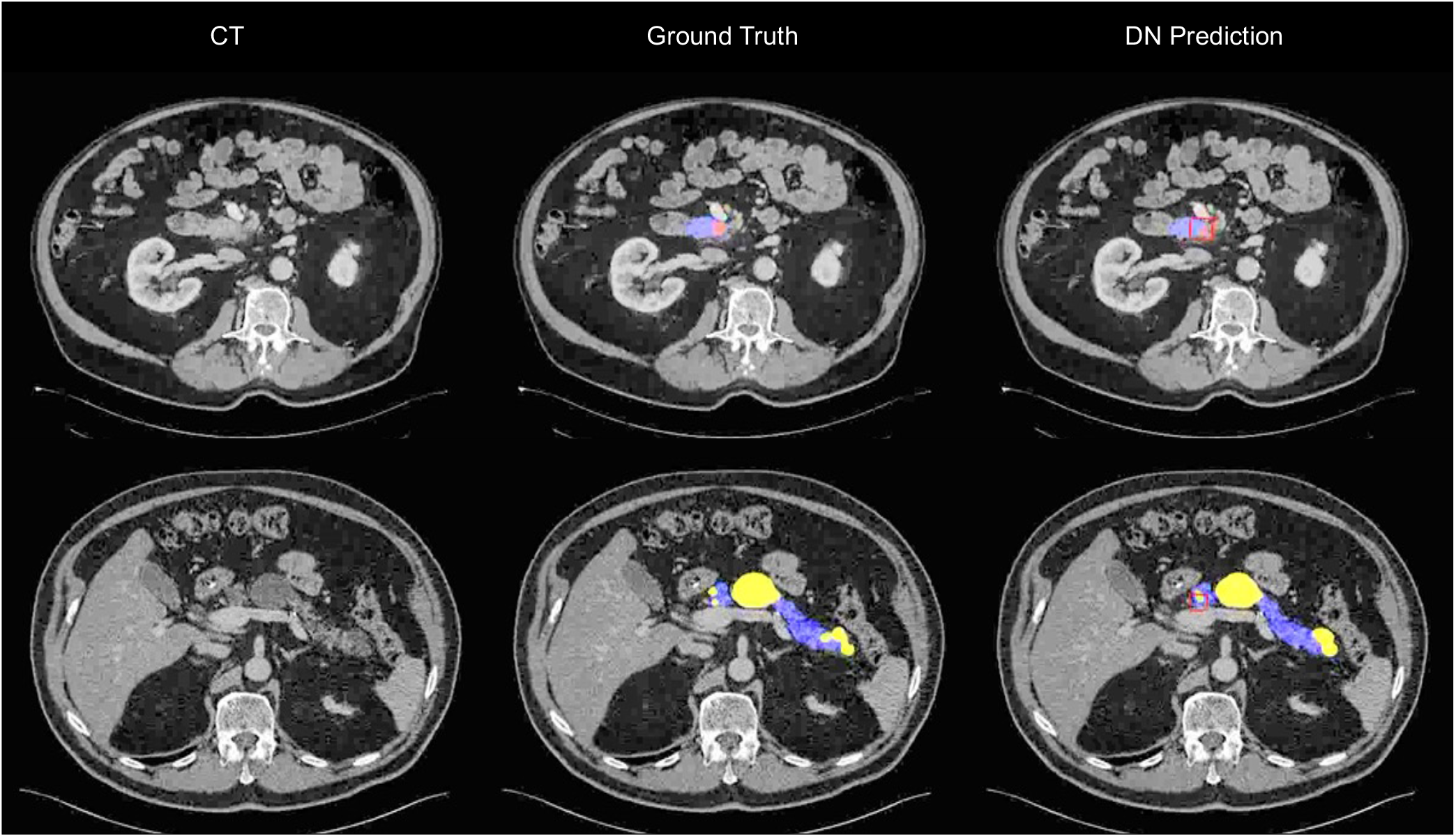
Visualizations of PDAC (top) and Cyst (bottom) false negatives. Our predictions (framed in red boxes) are close enough to the ground truth and therefore could be counted as true positives after radiologist re-review.

**Figure 14.**
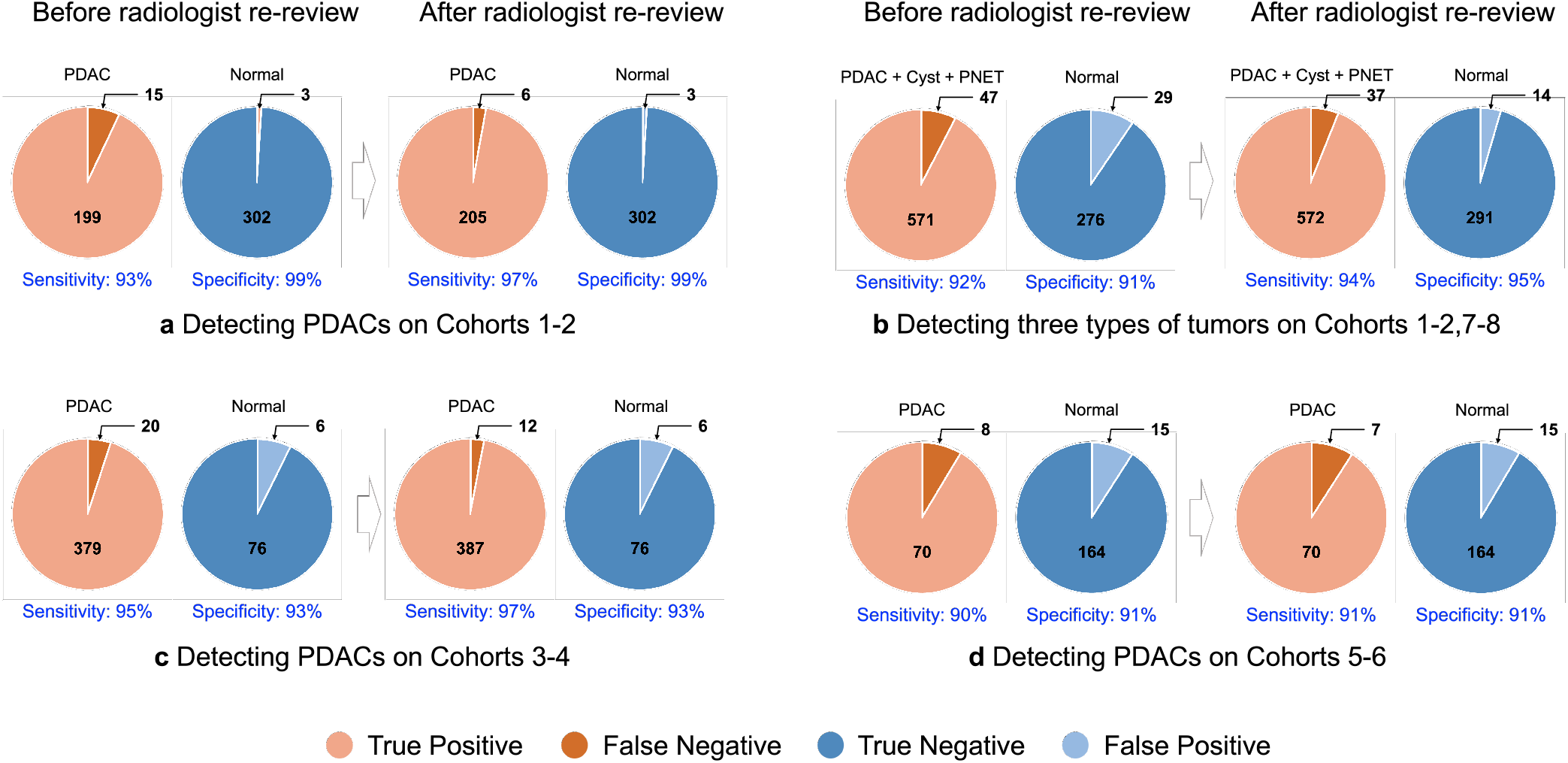
Tumor detection performance before and after radiologist re-review.

### Recognizing other pancreatic tumor types

We trained our AI algorithms to detect all these types of tumors while allowing only a few modifications to our algorithms. The overall performance remained high with sensitivity and specificity of 92.4% and 90.5% before radiologist re-review and 93.9% and 95.4% after radiologist re-review (Figure 14b). They only decrease to sensitivity and specificity of 94.4% and 93.0% before radiologist re-review and 94.8% and 94.3% after radiologist re-review if only the venous phase was used. The segmentation/localization of these tumors remained accurate (DSC scores of 87.0% for the pancreas, 62.42% for PDACs, 62.04% for cyst, and 55.16% for PanNETs). The algorithms were even able to detect some cysts as small as 2mm radius/diameter, which is close to the absolute performance limit of radiologists. Radiologist re-review was particularly useful as the algorithms often detected small cysts that had not been originally annotated by radiologists.

We also studied how performance varied with the size of the tumors. The distributions of sizes of tumors and how size predicted performance are given in Figure 8. We also modified the algorithm slightly as described in §4, using multiscale processing, in order to improve performance on tumors with sizes of less than 2cm diameter. This yielded a sensitivity and specificity of 88.7% and 84.9% before radiologist re-review, and 89.33% and 88.20% after. The DSC score for small tumor segmentation was 52.86%. We conclude that the algorithms could also detect and localize these three types of tumors with very high sensitivity and specificity and performed well even on very small tumors.

### Recognizing PDAC in CT images from other institutions

FELIX 1.2 was trained on Cohort 1 and 2, with modifications described in §4, and tested on Cohort 4. As before, we record a correct detection only if we also correctly localize the PDAC. This produced a sensitivity of 95.0% before radiologist re-review and 97.0% after radiologist re-review (Figure 14c). We achieve DSC scores of 82.8% for the pancreas and 58.4% for PDACs. It was impossible to measure the specificity since all the CTs in Cohort 4 contained PDACs. To do an alternative check of specificity we used Cohort 3 of 82 scans as a surrogate for normal cases. This gave specificity results of 92.7% both before and after radiologist re-review, which is lower than observed with Cohorts 1 and 2 but still acceptable.

Furthermore, we applied FELIX 1.2 to the Heidelberg dataset using the same training as for Cohort 4. This dataset was also annotated with the pancreas and PDACs. This dataset contained new challenges because, for example, the positioning of the patients in some of the scans differed from those in the Hopkins dataset by 30 degrees or more (this is a protocol used at Heidelberg to make it easier to detect tumors). For venous only, we obtained a sensitivity of 91.3% and specificity of 94.8%; for arterial only, we obtained a sensitivity of 95.7% and specificity of 91.4%. For dual-phase, we get 90.9% sensitivity and 91.6% specificity (Figure 14d). We achieved DSC scores of 82.2% for the pancreas and 54.3% for PDAC segmentation. These results were without checking for localization.

