## Supplementary material for "The FELIX Project: Deep Networks To Detect Pancreatic Neoplasms": Dataset information

| Index | Gender (M=0, F=1) | Death (1=Y, 0=N) as of 8/24/2022 | Age at the time of scan | Path Report - Panc | Stent (0=no, 1=yes) | Tumor Size |
|---|---|---|---|---|---|---|
| 1 | 1 | 0 | 51-55 | RENAL DONOR | | |
| 2 | 1 | 0 | 41-45 | RENAL DONOR | | |
| 3 | 1 | 0 | 41-45 | RENAL DONOR | | |
| 4 | 1 | 0 | 56-60 | RENAL DONOR | | |
| 5 | 1 | 0 | 51-55 | RENAL DONOR | | |
| 6 | 0 | 0 | 51-55 | RENAL DONOR | | |
| 7 | 0 | 0 | 51-55 | RENAL DONOR | | |
| 8 | 1 | 0 | 51-55 | RENAL DONOR | | |
| 9 | 0 | 0 | 66-70 | RENAL DONOR | | |
| 10 | 1 | 0 | 51-55 | RENAL DONOR | | |
| 11 | 1 | 0 | 51-55 | RENAL DONOR | | |
| 12 | 1 | 0 | 41-45 | RENAL DONOR | | |
| 13 | 1 | 0 | 36-40 | RENAL DONOR | | |
| 14 | 0 | 0 | 36-40 | RENAL DONOR | | |
| 15 | 1 | 0 | 51-55 | RENAL DONOR | | |
| 16 | 1 | 0 | 51-55 | RENAL DONOR | | |
| 17 | 1 | 0 | 46-50 | RENAL DONOR | | |
| 18 | 0 | 0 | 46-50 | RENAL DONOR | | |
| 19 | 0 | 0 | 21-25 | RENAL DONOR | | |
| 20 | 1 | 0 | 66-70 | RENAL DONOR | | |
| 21 | 1 | 0 | 51-55 | RENAL DONOR | | |
| 22 | 0 | 0 | 51-55 | RENAL DONOR | | |
| 23 | 1 | 0 | 46-50 | RENAL DONOR | | |
| 24 | 1 | 0 | 36-40 | RENAL DONOR | | |
| 25 | 0 | 0 | 46-50 | RENAL DONOR | | |
| 26 | 0 | 0 | 21-25 | RENAL DONOR | | |
| 27 | 0 | 0 | 36-40 | RENAL DONOR | | |
| 28 | 0 | 0 | 51-55 | RENAL DONOR | | |
| 29 | 0 | 0 | 56-60 | RENAL DONOR | | |
| 30 | 0 | 0 | 36-40 | RENAL DONOR | | |
| 31 | 1 | 0 | 61-65 | RENAL DONOR | | |
| 32 | 1 | 0 | 51-55 | RENAL DONOR | | |
| 33 | 0 | 0 | 61-65 | RENAL DONOR | | |
| 34 | 1 | 0 | 46-50 | RENAL DONOR | | |
| 35 | 1 | 0 | 51-55 | RENAL DONOR | | |
| 36 | 1 | 0 | 56-60 | RENAL DONOR | | |
| 37 | 0 | 0 | 36-40 | RENAL DONOR | | |
| 38 | 1 | 0 | 31-35 | RENAL DONOR | | |
| 39 | 0 | 0 | 46-50 | RENAL DONOR | | |
| 40 | 0 | 0 | 36-40 | RENAL DONOR | | |
| 41 | 0 | 0 | 31-35 | RENAL DONOR | | |
| 42 | 1 | 0 | 46-50 | RENAL DONOR | | |
| 43 | 1 | 0 | 41-45 | RENAL DONOR | | |
| 44 | 0 | 0 | 21-25 | RENAL DONOR | | |
| 45 | 0 | 0 | 46-50 | RENAL DONOR | | |
| 46 | 1 | 0 | 46-50 | RENAL DONOR | | |
| 47 | 0 | 0 | 31-35 | RENAL DONOR | | |
| 48 | 1 | 0 | 46-50 | RENAL DONOR | | |
| 49 | 0 | 0 | 26-30 | RENAL DONOR | | |
| 50 | 0 | 0 | 51-55 | RENAL DONOR | | |
| 51 | 1 | 0 | 36-40 | RENAL DONOR | | |
| 52 | 1 | 0 | 41-45 | RENAL DONOR | | |
| 53 | 1 | 0 | 36-40 | RENAL DONOR | | |
| 54 | 0 | 0 | 51-55 | RENAL DONOR | | |
| 55 | 1 | 0 | 41-45 | RENAL DONOR | | |
| 56 | 1 | 0 | 51-55 | RENAL DONOR | | |
| 57 | 1 | 0 | 26-30 | RENAL DONOR | | |
| 58 | 1 | 0 | 56-60 | RENAL DONOR | | |
| 59 | 1 | 0 | 31-35 | RENAL DONOR | | |
| 60 | 1 | 0 | 41-45 | RENAL DONOR | | |
| 61 | 1 | 0 | 61-65 | RENAL DONOR | | |
| 62 | 1 | 0 | 41-45 | RENAL DONOR | | |
| 63 | 0 | 0 | 41-45 | RENAL DONOR | | |
| 64 | 1 | 0 | 51-55 | RENAL DONOR | | |
| 65 | 1 | 0 | 46-50 | RENAL DONOR | | |
| 66 | 1 | 0 | 41-45 | RENAL DONOR | | |
| 67 | 0 | 0 | 36-40 | RENAL DONOR | | |
| 68 | 0 | 0 | 61-65 | RENAL DONOR | | |
| 69 | 1 | 0 | 66-70 | RENAL DONOR | | |
| 70 | 0 | 0 | 56-60 | RENAL DONOR | | |
| 71 | 0 | 0 | 66-70 | RENAL DONOR | | |
| 72 | 1 | 0 | 36-40 | RENAL DONOR | | |
| 73 | 1 | 0 | 56-60 | RENAL DONOR | | |
| 74 | 0 | 0 | 56-60 | RENAL DONOR | | |
| 75 | 1 | 0 | 41-45 | RENAL DONOR | | |
| 76 | 1 | 0 | 66-70 | RENAL DONOR | | |
| 77 | 1 | 0 | 26-30 | RENAL DONOR | | |
| 78 | 1 | 0 | 46-50 | RENAL DONOR | | |
| 79 | 1 | 0 | 36-40 | RENAL DONOR | | |
| 80 | 1 | 0 | 46-50 | RENAL DONOR | | |
| 81 | 1 | 0 | 46-50 | RENAL DONOR | | |
| 82 | 1 | 0 | 31-35 | RENAL DONOR | | |
| 83 | 1 | 0 | 31-35 | RENAL DONOR | | |
| 84 | 0 | 0 | 31-35 | RENAL DONOR | | |
| 85 | 0 | 0 | 41-45 | RENAL DONOR | | |
| 86 | 0 | 0 | 26-30 | RENAL DONOR | | |
| 87 | 1 | 0 | 36-40 | RENAL DONOR | | |
| 88 | 0 | 0 | 41-45 | RENAL DONOR | | |
| 89 | 1 | 0 | 56-60 | RENAL DONOR | | |
| 90 | 0 | 0 | 51-55 | RENAL DONOR | | |
| 91 | 0 | 0 | 36-40 | RENAL DONOR | | |
| 92 | 0 | 0 | 36-40 | RENAL DONOR | | |
| 93 | 1 | 0 | 41-45 | RENAL DONOR | | |
| 94 | 1 | 0 | 36-40 | RENAL DONOR | | |
| 95 | 0 | 0 | 56-60 | RENAL DONOR | | |
| 96 | 1 | 0 | 61-65 | RENAL DONOR | | |

| | | | | |
|---|---|---|---|---|
| 97 | 1 | 0 | 36-40 | RENAL DONOR |
| 98 | 1 | 0 | 46-50 | RENAL DONOR |
| 99 | 0 | 0 | 51-55 | RENAL DONOR |
| 100 | 1 | 0 | 31-35 | RENAL DONOR |
| 101 | 1 | 0 | 51-55 | RENAL DONOR |
| 102 | 0 | 0 | 56-60 | RENAL DONOR |
| 103 | 1 | 0 | 46-50 | RENAL DONOR |
| 104 | 0 | 0 | 31-35 | RENAL DONOR |
| 105 | 0 | 0 | 26-30 | RENAL DONOR |
| 106 | 0 | 0 | 26-30 | RENAL DONOR |
| 107 | 1 | 0 | 46-50 | RENAL DONOR |
| 108 | 0 | 0 | 26-30 | RENAL DONOR |
| 109 | 1 | 0 | 51-55 | RENAL DONOR |
| 110 | 0 | 0 | 41-45 | RENAL DONOR |
| 111 | 1 | 0 | 56-60 | RENAL DONOR |
| 112 | 1 | 0 | 36-40 | RENAL DONOR |
| 113 | 0 | 0 | 51-55 | RENAL DONOR |
| 114 | 0 | 0 | 41-45 | RENAL DONOR |
| 115 | 0 | 0 | 66-70 | RENAL DONOR |
| 116 | 0 | 0 | 61-65 | RENAL DONOR |
| 117 | 1 | 0 | 41-45 | RENAL DONOR |
| 118 | 1 | 0 | 41-45 | RENAL DONOR |
| 119 | 1 | 0 | 21-25 | RENAL DONOR |
| 120 | 1 | 0 | 41-45 | RENAL DONOR |
| 121 | 1 | 0 | 51-55 | RENAL DONOR |
| 122 | 1 | 0 | 61-65 | RENAL DONOR |
| 123 | 0 | 0 | 41-45 | RENAL DONOR |
| 124 | 1 | 0 | 36-40 | RENAL DONOR |
| 125 | 1 | 0 | 41-45 | RENAL DONOR |
| 126 | 1 | 0 | 56-60 | RENAL DONOR |
| 127 | 1 | 0 | 36-40 | RENAL DONOR |
| 128 | 0 | 0 | 36-40 | RENAL DONOR |
| 129 | 0 | 0 | 51-55 | RENAL DONOR |
| 130 | 1 | 0 | 51-55 | RENAL DONOR |
| 131 | 1 | 0 | 61-65 | RENAL DONOR |
| 132 | 1 | 0 | 51-55 | RENAL DONOR |
| 133 | 0 | 0 | 36-40 | RENAL DONOR |
| 134 | 0 | 0 | 16-20 | RENAL DONOR |
| 135 | 1 | 0 | 31-35 | RENAL DONOR |
| 136 | 1 | 0 | 66-70 | RENAL DONOR |
| 137 | 0 | 0 | 41-45 | RENAL DONOR |
| 138 | 1 | 0 | 46-50 | RENAL DONOR |
| 139 | 1 | 0 | 31-35 | RENAL DONOR |
| 140 | 1 | 0 | 51-55 | RENAL DONOR |
| 141 | 0 | 0 | 51-55 | RENAL DONOR |
| 142 | 0 | 0 | 41-45 | RENAL DONOR |
| 143 | 1 | 0 | 61-65 | RENAL DONOR |
| 144 | 1 | 0 | 46-50 | RENAL DONOR |
| 145 | 1 | 0 | 36-40 | RENAL DONOR |
| 146 | 1 | 0 | 46-50 | RENAL DONOR |
| 147 | 1 | 0 | 21-25 | RENAL DONOR |
| 148 | 1 | 0 | 56-60 | |
| 149 | 1 | 0 | 46-50 | RENAL DONOR |
| 150 | 0 | 0 | 56-60 | RENAL DONOR |
| 151 | 1 | 0 | 51-55 | RENAL DONOR |
| 152 | 0 | 0 | 51-55 | RENAL DONOR |
| 153 | 1 | 0 | 26-30 | RENAL DONOR |
| 154 | 1 | 0 | 51-55 | RENAL DONOR |
| 155 | 1 | 0 | 21-25 | RENAL DONOR |
| 156 | 0 | 0 | 26-30 | RENAL DONOR |
| 157 | 1 | 0 | 56-60 | RENAL DONOR |
| 158 | 1 | 0 | 26-30 | RENAL DONOR |
| 159 | 0 | 0 | 36-40 | RENAL DONOR |
| 160 | 0 | 0 | 51-55 | RENAL DONOR |
| 161 | 1 | 0 | 46-50 | RENAL DONOR |
| 162 | 1 | 0 | 41-45 | RENAL DONOR |
| 163 | 0 | 0 | 36-40 | RENAL DONOR |
| 164 | 1 | 0 | 41-45 | RENAL DONOR |
| 165 | 0 | 0 | 41-45 | RENAL DONOR |
| 166 | 0 | 0 | 31-35 | RENAL DONOR |
| 167 | 1 | 0 | 31-35 | RENAL DONOR |
| 168 | 1 | 0 | 46-50 | RENAL DONOR |
| 169 | 0 | 0 | 51-55 | RENAL DONOR |
| 170 | 0 | 0 | 41-45 | RENAL DONOR |
| 171 | 0 | 0 | 16-20 | RENAL DONOR |
| 172 | 0 | 0 | 31-35 | RENAL DONOR |
| 173 | 0 | 0 | 26-30 | RENAL DONOR |
| 174 | 0 | 0 | 51-55 | RENAL DONOR |
| 175 | 1 | 0 | 26-30 | RENAL DONOR |
| 176 | 1 | 0 | 26-30 | RENAL DONOR |
| 177 | 1 | 0 | 26-30 | RENAL DONOR |
| 178 | 0 | 0 | 26-30 | RENAL DONOR |
| 179 | 0 | 0 | 31-35 | RENAL DONOR |
| 180 | 0 | 0 | 31-35 | RENAL DONOR |
| 181 | 1 | 0 | 56-60 | RENAL DONOR |
| 182 | 1 | 0 | 46-50 | RENAL DONOR |
| 183 | 0 | 0 | 36-40 | RENAL DONOR |
| 184 | 1 | 0 | 31-35 | RENAL DONOR |
| 185 | 0 | 0 | 31-35 | RENAL DONOR |
| 186 | 1 | 0 | 41-45 | RENAL DONOR |
| 187 | 1 | 0 | 26-30 | RENAL DONOR |
| 188 | 1 | 0 | 36-40 | RENAL DONOR |
| 189 | 1 | 0 | 51-55 | RENAL DONOR |
| 190 | 1 | 0 | 26-30 | RENAL DONOR |
| 191 | 0 | 0 | 36-40 | RENAL DONOR |
| 192 | 1 | 0 | 56-60 | RENAL DONOR |

| | | | | |
|---|---|---|---|---|
| 193 | 1 | 0 | 56-60 | RENAL DONOR |
| 194 | 1 | 0 | 56-60 | RENAL DONOR |
| 195 | 1 | 0 | 56-60 | RENAL DONOR |
| 196 | 1 | 0 | 51-55 | RENAL DONOR |
| 197 | 1 | 0 | 66-70 | RENAL DONOR |
| 198 | 1 | 0 | 41-45 | RENAL DONOR |
| 199 | 1 | 0 | | RENAL DONOR |
| 200 | 0 | 0 | 16-20 | RENAL DONOR |
| 201 | 0 | 0 | 46-50 | RENAL DONOR |
| 202 | 1 | 0 | 41-45 | RENAL DONOR |
| 203 | 0 | 0 | 61-65 | RENAL DONOR |
| 204 | 0 | 0 | 61-65 | RENAL DONOR |
| 205 | 1 | 0 | 61-65 | RENAL DONOR |
| 206 | 1 | 0 | 61-65 | RENAL DONOR |
| 207 | 1 | 0 | 56-60 | RENAL DONOR |
| 208 | 0 | 0 | 26-30 | RENAL DONOR |
| 209 | 0 | 0 | 66-70 | RENAL DONOR |
| 210 | 0 | 0 | 46-50 | RENAL DONOR |
| 211 | 1 | 0 | 51-55 | RENAL DONOR |
| 212 | 1 | 0 | 51-55 | RENAL DONOR |
| 213 | 1 | 0 | 46-50 | RENAL DONOR |
| 214 | 1 | 0 | 46-50 | RENAL DONOR |
| 215 | 1 | 0 | 41-45 | RENAL DONOR |
| 216 | 1 | 0 | 31-35 | RENAL DONOR |
| 217 | 1 | 0 | 26-30 | RENAL DONOR |
| 218 | 1 | 0 | 36-40 | RENAL DONOR |
| 219 | 0 | 0 | 56-60 | RENAL DONOR |
| 220 | 1 | 0 | 61-65 | RENAL DONOR |
| 221 | 1 | 0 | 66-70 | RENAL DONOR |
| 222 | 1 | 0 | 61-65 | RENAL DONOR |
| 223 | 1 | 0 | 66-70 | RENAL DONOR |
| 224 | 1 | 0 | 51-55 | RENAL DONOR |
| 225 | 1 | 0 | 66-70 | RENAL DONOR |
| 226 | 1 | 0 | 61-65 | RENAL DONOR |
| 227 | 0 | 0 | 66-70 | RENAL DONOR |
| 228 | 1 | 0 | 61-65 | RENAL DONOR |
| 229 | 1 | 0 | 61-65 | RENAL DONOR |
| 230 | 0 | 0 | 56-60 | RENAL DONOR |
| 231 | 1 | 0 | 51-55 | RENAL DONOR |
| 232 | 1 | 0 | 56-60 | RENAL DONOR |
| 233 | 1 | 0 | 51-55 | RENAL DONOR |
| 234 | 1 | 0 | 51-55 | RENAL DONOR |
| 235 | 1 | 0 | 46-50 | RENAL DONOR |
| 236 | 1 | 0 | 41-45 | RENAL DONOR |
| 237 | 0 | 0 | 61-65 | RENAL DONOR |
| 238 | 1 | 0 | 46-50 | RENAL DONOR |
| 239 | 1 | 0 | 56-60 | RENAL DONOR |
| 240 | 1 | 0 | 71-75 | RENAL DONOR |
| 241 | 1 | 0 | 56-60 | RENAL DONOR |
| 242 | 1 | 0 | 66-70 | RENAL DONOR |
| 243 | 0 | 0 | 61-65 | RENAL DONOR |
| 244 | 1 | 0 | 41-45 | RENAL DONOR |
| 245 | 1 | 0 | 41-45 | RENAL DONOR |
| 246 | 1 | 0 | 41-45 | RENAL DONOR |
| 247 | 1 | 0 | 51-55 | RENAL DONOR |
| 248 | 0 | 0 | 51-55 | RENAL DONOR |
| 249 | 0 | 0 | 51-55 | RENAL DONOR |
| 250 | 1 | 0 | 56-60 | RENAL DONOR |
| 251 | 1 | 0 | 51-55 | RENAL DONOR |
| 252 | 1 | 0 | 51-55 | RENAL DONOR |
| 253 | 1 | 0 | 46-50 | RENAL DONOR |
| 254 | 1 | 0 | 46-50 | RENAL DONOR |
| 255 | 0 | 0 | 46-50 | RENAL DONOR |
| 256 | 0 | 0 | 46-50 | RENAL DONOR |
| 257 | 1 | 0 | 46-50 | RENAL DONOR |
| 258 | 1 | 0 | 46-50 | RENAL DONOR |
| 259 | 1 | 0 | 46-50 | RENAL DONOR |
| 260 | 1 | 0 | 51-55 | RENAL DONOR |
| 261 | 1 | 0 | 51-55 | RENAL DONOR |
| 262 | 1 | 0 | 51-55 | RENAL DONOR |
| 263 | 1 | 0 | 46-50 | RENAL DONOR |
| 264 | 0 | 0 | 41-45 | RENAL DONOR |
| 265 | 0 | 0 | 46-50 | RENAL DONOR |
| 266 | 1 | 0 | 46-50 | RENAL DONOR |
| 267 | 1 | 0 | 61-65 | RENAL DONOR |
| 268 | 1 | 0 | 61-65 | RENAL DONOR |
| 269 | 0 | 0 | 61-65 | RENAL DONOR |
| 270 | 0 | 0 | 56-60 | RENAL DONOR |
| 271 | 0 | 0 | 56-60 | RENAL DONOR |
| 272 | 1 | 0 | 46-50 | RENAL DONOR |
| 273 | 1 | 0 | 51-55 | RENAL DONOR |
| 274 | 1 | 0 | 51-55 | RENAL DONOR |
| 275 | 0 | 0 | 46-50 | RENAL DONOR |
| 276 | 1 | 0 | 66-70 | RENAL DONOR |
| 277 | 1 | 0 | 51-55 | RENAL DONOR |
| 278 | 1 | 0 | 56-60 | RENAL DONOR |
| 279 | 1 | 0 | 51-55 | RENAL DONOR |
| 280 | 1 | 0 | 46-50 | RENAL DONOR |
| 281 | 0 | 0 | 46-50 | RENAL DONOR |
| 282 | 0 | 0 | 56-60 | RENAL DONOR |
| 283 | 0 | 0 | 66-70 | RENAL DONOR |
| 284 | 0 | 0 | 61-65 | RENAL DONOR |
| 285 | 1 | 0 | 41-45 | RENAL DONOR |
| 286 | 1 | 0 | 46-50 | RENAL DONOR |
| 287 | 0 | 0 | 46-50 | RENAL DONOR |
| 288 | 1 | 0 | 51-55 | RENAL DONOR |
| 289 | 0 | 0 | 41-45 | RENAL DONOR |

| | | | | | | |
|---|---|---|---|---|---|---|
| 290 | 1 | 0 | 46-50 | RENAL DONOR | | |
| 291 | 1 | 0 | 51-55 | RENAL DONOR | | |
| 292 | 1 | 0 | 66-70 | RENAL DONOR | | |
| 293 | 0 | 0 | 66-70 | RENAL DONOR | | |
| 294 | 0 | 0 | 51-55 | RENAL DONOR | | |
| 295 | 1 | 0 | 41-45 | RENAL DONOR | | |
| 296 | 1 | 0 | 36-40 | RENAL DONOR | | |
| 297 | 1 | 0 | 41-45 | RENAL DONOR | | |
| 298 | 1 | 0 | 46-50 | RENAL DONOR | | |
| 299 | 1 | 0 | 41-45 | RENAL DONOR | | |
| 300 | 1 | 0 | 66-70 | RENAL DONOR | | |
| 301 | 1 | 0 | 41-45 | RENAL DONOR | | |
| 302 | 1 | 0 | 46-50 | RENAL DONOR | | |
| 303 | 1 | 0 | 66-70 | RENAL DONOR | | |
| 304 | 1 | 0 | 46-50 | RENAL DONOR | | |
| 305 | 0 | 0 | 51-55 | RENAL DONOR | | |
| 306 | 1 | 0 | 41-45 | RENAL DONOR | | |
| 307 | 1 | 0 | 46-50 | RENAL DONOR | | |
| 308 | 0 | 0 | 41-45 | RENAL DONOR | | |
| 309 | 0 | 0 | 46-50 | RENAL DONOR | | |
| 310 | 1 | 0 | 36-40 | RENAL DONOR | | |
| 311 | 1 | 0 | 61-65 | RENAL DONOR | | |
| 312 | 0 | 0 | 46-50 | RENAL DONOR | | |
| 313 | 1 | 0 | 51-55 | RENAL DONOR | | |
| 314 | 1 | 0 | 41-45 | RENAL DONOR | | |
| 315 | 1 | 0 | 51-55 | RENAL DONOR | | |
| 316 | 1 | 0 | 61-65 | RENAL DONOR | | |
| 317 | 0 | 0 | 56-60 | RENAL DONOR | | |
| 318 | 0 | 0 | 46-50 | RENAL DONOR | | |
| 319 | 1 | 0 | 26-30 | RENAL DONOR | | |
| 320 | 1 | 0 | 81-85 | PDAC | 0 | 1.1 |
| 321 | 0 | 0 | 51-55 | PDAC | 0 | 3.1 |
| 322 | 1 | 0 | 71-75 | PDAC | 1 | 2.5 |
| 323 | 1 | 1 | 61-65 | PDAC | 0 | 4.6 |
| 324 | 1 | 0 | 76-80 | PDAC | 0 | 1.9 |
| 325 | 1 | 0 | 66-70 | PDAC | 0 | 6.1 |
| 326 | 0 | 0 | 71-75 | PDAC | 0 | 3 |
| 327 | 0 | 1 | 46-50 | PDAC | 1 | 5.3 |
| 328 | 0 | 0 | 66-70 | PDAC | 0 | 4.5 |
| 329 | 1 | 0 | 66-70 | PDAC | 0 | 1.3 |
| 330 | 1 | 0 | 61-65 | PDAC | 0 | 2.1 |
| 331 | 1 | 0 | 66-70 | PDAC | 0 | 3 |
| 332 | 1 | 0 | 56-60 | PDAC | 0 | 2.8 |
| 333 | 1 | 0 | 81-85 | PDAC, IPMN | 0 | 1.8 |
| 334 | 1 | 1 | 76-80 | PDAC | 0 | 2.9 |
| 335 | 0 | 0 | 51-55 | PDAC | 0 | 7.9 |
| 336 | 0 | 0 | 76-80 | PDAC, IPMN | 0 | 4.2 |
| 337 | 0 | 1 | 56-60 | PDAC | 0 | 5.1 |
| 338 | 0 | 1 | 76-80 | PDAC | 0 | 4.7 |
| 339 | 0 | 0 | 76-80 | PDAC, IPMN | 0 | 2.1 |
| 340 | 0 | 1 | 71-75 | PDAC | 0 | 3.9 |
| 341 | 0 | 0 | 56-60 | PDAC | 0 | 1.4 |
| 342 | 1 | 0 | 61-65 | PDAC | 0 | 2 |
| 343 | 1 | 0 | 76-80 | PDAC | 0 | 3.1 |
| 344 | 0 | 0 | 56-60 | PDAC | 0 | 1.8 |
| 345 | 1 | 0 | 36-40 | PDAC | 0 | 5.7 |
| 346 | 1 | 1 | 66-70 | PDAC | 0 | 3.1 |
| 347 | 0 | 0 | 66-70 | PDAC | 0 | 1.6 |
| 348 | 0 | 1 | 86-90 | PDAC | 0 | 4.7 |
| 349 | 1 | 0 | 56-60 | PDAC | 0 | 2.6 |
| 350 | 0 | 0 | 41-45 | PDAC | 0 | 4.3 |
| 351 | 0 | 1 | 66-70 | PDAC | 0 | 9.4 |
| 352 | 1 | 1 | 61-65 | PDAC | 0 | 2.7 |
| 353 | 0 | 1 | 66-70 | PDAC | 0 | 3 |
| 354 | 0 | 1 | 66-70 | PDAC | 0 | 2.9 |
| 355 | 0 | 0 | 76-80 | PDAC | 1 | 2.3 |
| 356 | 0 | 1 | 51-55 | PDAC | 1 | 2 |
| 357 | 0 | 0 | 56-60 | PDAC | 0 | 3.2 |
| 358 | 1 | 1 | 76-80 | PDAC | 0 | 1.6 |
| 359 | 1 | 1 | 56-60 | PDAC | 0 | 2.2 |
| 360 | 0 | 1 | 66-70 | PDAC | 0 | 3.1 |
| 361 | 0 | 1 | 41-45 | PDAC | 0 | 2.8 |
| 362 | 0 | 0 | 66-70 | PDAC | 0 | 2.3 |
| 363 | 1 | 0 | 66-70 | PDAC | 0 | 1.9 |
| 364 | 0 | 1 | 71-75 | PDAC | 0 | 2.2 |
| 365 | 1 | 0 | 66-70 | PDAC | 0 | 4.4 |
| 366 | 0 | 0 | 76-80 | PDAC | 0 | 2.1 |
| 367 | 0 | 1 | 66-70 | PDAC | 0 | 2 |
| 368 | 1 | 0 | 66-70 | PDAC | 0 | 3.3 |
| 369 | 0 | 0 | 66-70 | PDAC | 1 | 3.3 |
| 370 | 0 | 0 | 51-55 | PDAC | 0 | 3.7 |
| 371 | 0 | 0 | 61-65 | PDAC | 1 | 5.7 |
| 372 | 1 | 0 | 76-80 | PDAC | 0 | 3.2 |
| 373 | 1 | 0 | 76-80 | PDAC | 0 | 3 |
| 374 | 1 | 1 | 51-55 | PDAC | 1 | 1.1 |
| 375 | 1 | 0 | 71-75 | PDAC | 0 | 2 |
| 376 | 1 | 1 | 61-65 | PDAC | 0 | N/A |
| 377 | 1 | 0 | 46-50 | PDAC | 0 | 0.3 |
| 378 | 0 | 0 | 46-50 | PDAC | 0 | 3.4 |
| 379 | 1 | 0 | 61-65 | PDAC | 0 | 2 |
| 380 | 0 | 1 | 71-75 | PDAC | 0 | 8.1 |
| 381 | 1 | 1 | 56-60 | PDAC | 1 | 2.6 |
| 382 | 0 | 0 | 76-80 | PDAC | 1 | 4.6 |
| 383 | 0 | 0 | 66-70 | PDAC | 0 | 1 |
| 384 | 0 | 1 | 66-70 | PDAC | 0 | 1 |
| 385 | 1 | 0 | 66-70 | PDAC | 0 | 3.4 |
| 386 | 0 | 0 | 81-85 | PDAC | 0 | 2.2 |

| | | | | | | |
|---|---|---|---|---|---|---|
| 387 | 0 | 0 | 56-60 | PDAC | 0 | 3.7 |
| 388 | 1 | 0 | 61-65 | PDAC | 0 | 2.5 |
| 389 | 1 | 1 | 56-60 | PDAC | 0 | 2.5 |
| 390 | 1 | 0 | 71-75 | PDAC | 0 | 2.5 |
| 391 | 1 | 0 | 51-55 | PDAC | 0 | 1.3 |
| 392 | 0 | 1 | 76-80 | PDAC | 0 | 1.1 |
| 393 | 0 | 0 | 66-70 | PDAC | 0 | 3.6 |
| 394 | 1 | 0 | 86-90 | PDAC | 0 | 1.3 |
| 395 | 1 | 0 | 56-60 | PDAC | 1 | 2.5 |
| 396 | 1 | 1 | 86-90 | PDAC | 1 | 2.5 |
| 397 | 1 | 0 | 66-70 | PDAC | 0 | 5 |
| 398 | 1 | 1 | 71-75 | PDAC | 0 | 3.6 |
| 399 | 1 | 0 | 76-80 | PDAC | 0 | 2 |
| 400 | 1 | 0 | 81-85 | PDAC | 0 | 8.2 |
| 401 | 0 | 0 | 66-70 | PDAC | 0 | 0.7 |
| 402 | 0 | 1 | 66-70 | PDAC | 0 | 5.1 |
| 403 | 1 | 0 | 51-55 | PDAC | 0 | 2 |
| 404 | 1 | 0 | 76-80 | PDAC | 0 | 1.2 |
| 405 | 0 | 0 | 76-80 | PDAC | 0 | 2.5 |
| 406 | 0 | 0 | 71-75 | PDAC | 1 | 3.1 |
| 407 | 0 | 0 | 76-80 | PDAC | 0 | 1.9 |
| 408 | 0 | 0 | 76-80 | PDAC | 0 | 2.6 |
| 409 | 0 | 1 | 61-65 | PDAC | 0 | 2.5 |
| 410 | 1 | 0 | 56-60 | PDAC | 0 | 4.2 |
| 411 | 0 | 0 | 56-60 | PDAC | 0 | 2.7 |
| 412 | 0 | 1 | 56-60 | PDAC | 0 | 5.5 |
| 413 | 0 | 0 | 76-80 | PDAC | 0 | 3.4 |
| 414 | 1 | 1 | 66-70 | PDAC | 0 | 3 |
| 415 | 1 | 0 | 61-65 | PDAC | 1 | 2.5 |
| 416 | 0 | 0 | 81-85 | PDAC | 0 | 4.5 |
| 417 | 1 | 0 | 76-80 | PDAC | 1 | 4.1 |
| 418 | 0 | 1 | 61-65 | PDAC | 0 | 4.9 |
| 419 | 0 | 1 | 66-70 | PDAC | 0 | 1.1 |
| 420 | 1 | 0 | 66-70 | PDAC | 0 | 2.5 |
| 421 | 0 | 0 | 61-65 | PDAC | 0 | 2.5 |
| 422 | 0 | 1 | 41-45 | PDAC | 0 | 3.6 |
| 423 | 1 | 0 | 51-55 | N/A | 0 | 2.9 |
| 424 | 1 | 0 | 51-55 | PanIN | 0 | 3 |
| 425 | 1 | 0 | 71-75 | PDAC | 1 | 4 |
| 426 | 1 | 0 | 61-65 | PDAC | 0 | 4 |
| 427 | 0 | 0 | 61-65 | PDAC | 1 | 4.8 |
| 428 | 0 | 1 | 56-60 | PDAC | 0 | 4.9 |
| 429 | 0 | 1 | 76-80 | PDAC | 0 | 3.2 |
| 430 | 0 | 0 | 51-55 | PDAC | 0 | 5.5 |
| 431 | 0 | 1 | 71-75 | PDAC | 0 | 5 |
| 432 | 1 | 0 | 71-75 | PDAC | 0 | 3.2 |
| 433 | 0 | 1 | 66-70 | PDAC | 0 | 3.3 |
| 434 | 0 | 0 | 56-60 | PDAC | 0 | 2.1 |
| 435 | 0 | 0 | 71-75 | PNET | 0 | 3.3 |
| 436 | 0 | 1 | 76-80 | PDAC | 0 | 3.5 |
| 437 | 1 | 1 | 66-70 | PDAC | 0 | 2.3 |
| 438 | 1 | 0 | 76-80 | PDAC | 1 | 1.7 |
| 439 | 1 | 0 | 56-60 | PNET | 0 | 4.3 |
| 440 | 0 | 0 | 61-65 | PDAC | 0 | 4.5 |
| 441 | 0 | 0 | 51-55 | PDAC | 1 | 3.5 |
| 442 | 0 | 0 | 71-75 | PDAC | 0 | 5 |
| 443 | 1 | 0 | 76-80 | PDAC | 0 | 2 |
| 444 | 0 | 0 | 81-85 | PDAC | 0 | 3.7 |
| 445 | 1 | 1 | 76-80 | PDAC | 0 | 2 |
| 446 | 1 | 1 | 61-65 | PDAC | 1 | 3 |
| 447 | 0 | 1 | 66-70 | PDAC | 0 | 5 |
| 448 | 1 | 1 | 61-65 | PDAC | 0 | 2.5 |
| 449 | 1 | 1 | 71-75 | PDAC | 0 | 3.2 |
| 450 | 1 | 1 | 71-75 | PDAC | 0 | 1.7 |
| 451 | 0 | 1 | 76-80 | PDAC | 1 | 3 |
| 452 | 1 | 0 | 61-65 | PDAC | 0 | 2.7 |
| 453 | 0 | 1 | 61-65 | PDAC | 0 | 6 |
| 454 | 1 | 0 | 76-80 | PDAC | 0 | 2 |
| 455 | 1 | 0 | 66-70 | PDAC | 0 | 2.8 |
| 456 | 0 | 1 | 66-70 | PDAC | 0 | 3.5 |
| 457 | 0 | 0 | 76-80 | PDAC | 0 | 2.5 |
| 458 | 1 | 0 | 61-65 | PDAC | 0 | 3.2 |
| 459 | 0 | 1 | 71-75 | PDAC | 0 | 2.5 |
| 460 | 0 | 1 | 56-60 | PDAC | 0 | 2.8 |
| 461 | 0 | 1 | 51-55 | PDAC | 0 | 2.4 |
| 462 | 0 | 1 | 61-65 | PDAC | 0 | 2.5 |
| 463 | 1 | 1 | 71-75 | PDAC | 0 | 3 |
| 464 | 0 | 0 | 61-65 | PDAC | 0 | 4 |
| 465 | 0 | 1 | 61-65 | PDAC | 0 | 5.8 |
| 466 | 1 | 1 | 71-75 | PDAC | 0 | 1.4 |
| 467 | 0 | 1 | 61-65 | PDAC | 0 | 4 |
| 468 | 0 | 0 | 71-75 | PDAC | 0 | 3.8 |
| 469 | 0 | 0 | 56-60 | PDAC | 0 | 3.6 |
| 470 | 0 | 0 | 56-60 | PDAC | 0 | 2.5 |
| 471 | 1 | 0 | 61-65 | PDAC | 0 | 1.5 |
| 472 | 0 | 1 | 61-65 | PDAC | 0 | 2.5 |
| 473 | 1 | 1 | 76-80 | PDAC | 0 | 2.3 |
| 474 | 1 | 0 | 76-80 | PDAC | 0 | 3 |
| 475 | 1 | 0 | 66-70 | PDAC | 0 | 3.5 |
| 476 | 1 | 1 | 76-80 | PDAC | 0 | 1.5 |
| 477 | 0 | 0 | 71-75 | PDAC | 0 | 2 |
| 478 | 1 | 1 | 51-55 | PDAC | 0 | 2.5 |
| 479 | 0 | 1 | 51-55 | PDAC | 0 | 3.5 |
| 480 | 0 | 1 | 86-90 | PDAC | 0 | 3.8 |
| 481 | 1 | 1 | 56-60 | PDAC | 0 | 2.5 |
| 482 | 1 | 0 | 56-60 | PDAC | 0 | 4.4 |
| 483 | 1 | 0 | 71-75 | PDAC | 0 | 4 |

| | | | | | | |
|---|---|---|---|---|---|---|
| 484 | 0 | 1 | 66-70 | PDAC | 0 | 5.5 |
| 485 | 0 | 0 | 66-70 | PDAC | 0 | 1.8 |
| 486 | 0 | 1 | 31-35 | PDAC | 0 | 8 |
| 487 | 0 | 1 | 61-65 | PDAC | 0 | 1.7 |
| 488 | 0 | 1 | 66-70 | PDAC | 0 | 4 |
| 489 | 1 | 1 | 41-45 | PDAC | 0 | 2.5 |
| 490 | 0 | 1 | 76-80 | PDAC | 0 | 1.5 |
| 491 | 1 | 0 | 66-70 | PDAC | 0 | 4.5 |
| 492 | 1 | 1 | 71-75 | PDAC | 0 | 4.6 |
| 493 | 0 | 1 | 56-60 | PDAC | 0 | 3 |
| 494 | 0 | 1 | 66-70 | PDAC | 0 | 3 |
| 495 | 0 | 0 | 51-55 | PDAC | 0 | 4.6 |
| 496 | 1 | 1 | 46-50 | PDAC | 0 | 3.2 |
| 497 | 0 | 1 | 61-65 | PDAC | 0 | 3.1 |
| 498 | 1 | 1 | 76-80 | PDAC | 0 | 2.6 |
| 499 | 0 | 1 | 76-80 | PDAC | 0 | 10 |
| 500 | 0 | 0 | 51-55 | PDAC | 0 | 3.8 |
| 501 | 0 | 0 | 31-35 | PDAC | 0 | N/A |
| 502 | 0 | 1 | 71-75 | PDAC | 0 | 4.9 |
| 503 | 1 | 1 | 61-65 | PDAC | 0 | 2.3 |
| 504 | 0 | 1 | 71-75 | PDAC | 0 | 4.8 |
| 505 | 0 | 1 | 61-65 | PDAC | 0 | 2.2 |
| 506 | 0 | 0 | 41-45 | PDAC | 0 | 6 |
| 507 | 0 | 1 | 56-60 | PDAC | 0 | 3.2 |
| 508 | 1 | 1 | 56-60 | PDAC | 0 | 3.8 |
| 509 | 1 | 1 | 71-75 | PDAC | 0 | 5.7 |
| 510 | 1 | 0 | 51-55 | PDAC | 0 | 4.7 |
| 511 | 1 | 1 | 66-70 | PDAC | 0 | 4 |
| 512 | 0 | 1 | 61-65 | PDAC | 0 | 2.2 |
| 513 | 1 | 1 | 66-70 | PDAC | 0 | 0.1 |
| 514 | 0 | 1 | 51-55 | PDAC | 0 | 2 |
| 515 | 0 | 1 | 76-80 | PDAC | 0 | 2.6 |
| 516 | 1 | 0 | 61-65 | PDAC | 0 | 4.5 |
| 517 | 1 | 1 | 76-80 | PDAC | 0 | 3.2 |
| 518 | 0 | 1 | 81-85 | PDAC | 0 | 2.1 |
| 519 | 0 | 1 | 61-65 | PDAC | 0 | 0.5 |
| 520 | 1 | 0 | 71-75 | PDAC | 0 | 5.2 |
| 521 | 1 | 0 | 51-55 | PDAC | 0 | 3.3 |
| 522 | 0 | 1 | 81-85 | PDAC | 0 | 7.2 |
| 523 | 0 | 0 | 56-60 | PDAC | 1 | 1.6 |
| 524 | 1 | 1 | 76-80 | PDAC | 0 | 2.5 |
| 525 | 1 | 0 | 61-65 | PDAC | 0 | 3.9 |
| 526 | 1 | 0 | 61-65 | PDAC | 0 | 5.9 |
| 527 | 0 | 1 | 46-50 | PDAC | 0 | 8.5 |
| 528 | 1 | 0 | 61-65 | PDAC | 0 | 1.1 |
| 529 | 0 | 0 | 56-60 | PDAC | 0 | 4.5 |
| 530 | 1 | 1 | 61-65 | PDAC | 0 | 5.4 |
| 531 | 1 | 0 | 61-65 | PDAC | 0 | 0.8 |
| 532 | 0 | 0 | 61-65 | PDAC | 0 | 2.9 |
| 533 | 1 | 0 | 76-80 | IPMN | 0 | 3.5 |
| 534 | 0 | 0 | 36-40 | | 0 | |
| 535 | 1 | 1 | 66-70 | PDAC | 0 | 2.7 |
| 536 | 0 | 1 | 51-55 | PDAC | 1 | 2.5 |
| 537 | 1 | 1 | 56-60 | PDAC | 0 | 1 |
| 538 | 1 | 0 | 66-70 | PDAC | 0 | 5 |
| 539 | 1 | 0 | 61-65 | PDAC | 1 | 0.6 |
| 540 | 0 | 0 | 61-65 | PDAC | 0 | 2.5 |
| 541 | 1 | 1 | 76-80 | PDAC | 1 | 2.5 |
| 542 | 1 | 1 | 56-60 | PDAC | 1 | 0.8 |
| 543 | 0 | 1 | 56-60 | PDAC | 0 | 2 |
| 544 | 1 | 0 | 61-65 | PDAC | 1 | 4.2 |
| 545 | 0 | 1 | 36-40 | PDAC | 0 | 2.8 |
| 546 | 1 | 1 | 56-60 | PDAC | 0 | 3.3 |
| 547 | 0 | 0 | 76-80 | PDAC | 0 | 6.4 |
| 548 | 1 | 1 | 66-70 | PDAC | 0 | 3.5 |
| 549 | 1 | 1 | 66-70 | PDAC | 0 | 3.9 |
| 550 | 0 | 1 | 61-65 | PDAC | 1 | 4.5 |
| 551 | 0 | 1 | 56-60 | PDAC | 1 | 6 |
| 552 | 1 | 1 | 66-70 | PDAC | 1 | 2.2 |
| 553 | 1 | 0 | 81-85 | PDAC | 0 | 2.2 |
| 554 | 1 | 0 | 66-70 | PDAC | 0 | 8.6 |
| 555 | 1 | 1 | 46-50 | PDAC | 0 | 2.4 |
| 556 | 1 | 1 | 56-60 | PDAC | 0 | 8 |
| 557 | 0 | 1 | 56-60 | PDAC | 1 | 3 |
| 558 | 1 | 1 | 66-70 | PDAC | 0 | 1.8 |
| 559 | 0 | 0 | 76-80 | PDAC | 0 | 2.2 |
| 560 | 0 | 1 | 66-70 | PDAC | 0 | 1.8 |
| 561 | 1 | 0 | 56-60 | PDAC | 0 | 2.2 |
| 562 | 0 | 1 | 61-65 | PDAC | 1 | 1.1 |
| 563 | 1 | 0 | 56-60 | PDAC | 0 | 4.2 |
| 564 | 1 | 1 | 81-85 | PDAC | 0 | 2 |
| 565 | 0 | 0 | 76-80 | | 0 | 5.1 |
| 566 | 1 | 0 | 56-60 | Suspicious for PDAC | 0 | 2.1 |
| 567 | 0 | 0 | 56-60 | PDAC | 0 | 0.6 |
| 568 | 0 | 0 | 61-65 | PDAC | 0 | 3.1 |
| 569 | 0 | 1 | 66-70 | PDAC | 0 | 3.1 |
| 570 | 0 | 1 | 71-75 | PDAC | 1 | 0.2 |
| 571 | 0 | 1 | 61-65 | PDAC | 0 | 3.5 |
| 572 | 1 | 0 | 46-50 | PDAC | 0 | 2.5 |
| 573 | 1 | 1 | 71-75 | PDAC | 1 | 3 |
| 574 | 1 | 1 | 66-70 | PDAC | 0 | 1.1 |
| 575 | 0 | 1 | 76-80 | PDAC | 0 | 2.7 |
| 576 | 1 | 0 | 86-90 | PDAC | 0 | 3 |
| 577 | 0 | 1 | 66-70 | PDAC | 0 | 10 |
| 578 | 0 | 0 | 66-70 | PDAC | 0 | 2 |
| 579 | 0 | 0 | 61-65 | PDAC | 0 | 2.2 |
| 580 | 0 | 1 | 66-70 | PDAC | 1 | 3.8 |

| | | | | | | |
|---|---|---|---|---|---|---|
| 581 | 1 | 1 | 71-75 | PDAC | 0 | 3 |
| 582 | 0 | 0 | 66-70 | PDAC | 0 | 6.2 |
| 583 | 1 | 1 | 71-75 | PDAC | 1 | 3 |
| 584 | 1 | 1 | 76-80 | PDAC | 1 | 1 |
| 585 | 1 | 1 | 66-70 | PDAC | 0 | 2.3 |
| 586 | 0 | 1 | 61-65 | PDAC | 1 | 1.5 |
| 587 | 0 | 1 | 71-75 | PDAC | 1 | 4.1 |
| 588 | 0 | 0 | 66-70 | PDAC | 0 | 3.8 |
| 589 | 1 | 1 | 51-55 | PDAC | 1 | 2 |
| 590 | 1 | 1 | 61-65 | PDAC | 0 | 1.4 |
| 591 | 0 | 1 | 81-85 | PDAC | 0 | 4 |
| 592 | 0 | 1 | 61-65 | PDAC | 1 | 3.6 |
| 593 | 0 | 1 | 66-70 | PDAC | 1 | 4 |
| 594 | 0 | 1 | 66-70 | PDAC | 0 | 1.5 |
| 595 | 1 | 0 | 56-60 | PDAC | 1 | 2.6 |
| 596 | 1 | 0 | 51-55 | PDAC | 1 | 4.4 |
| 597 | 1 | 0 | 56-60 | PDAC | 1 | 1.5 |
| 598 | 0 | 0 | 76-80 | PDAC | 0 | 2.5 |
| 599 | 1 | 1 | 41-45 | PDAC | 1 | 0.9 |
| 600 | 1 | 1 | 61-65 | PDAC | 0 | 4.8 |
| 601 | 0 | 1 | 66-70 | PDAC | 1 | 2.5 |
| 602 | 0 | 1 | 46-50 | PDAC | 1 | 2.6 |
| 603 | 1 | 1 | 71-75 | PDAC | 0 | 3 |
| 604 | 0 | 1 | 61-65 | PDAC | 1 | 2.4 |
| 605 | 0 | 1 | 66-70 | PDAC | 1 | 3 |
| 606 | 0 | 1 | 76-80 | PDAC | 0 | 2 |
| 607 | 0 | 1 | 76-80 | PDAC | 0 | 3.6 |
| 608 | 1 | 1 | 66-70 | PDAC | 0 | 1.7 |
| 609 | 1 | 1 | 31-35 | PDAC | 1 | 5 |
| 610 | 1 | 1 | 66-70 | PDAC | 0 | 1 |
| 611 | 1 | 1 | 66-70 | PDAC | 0 | 5.5 |
| 612 | 0 | 0 | 61-65 | PDAC | 0 | 2 |
| 613 | 1 | 1 | 76-80 | PDAC | 0 | 4.1 |
| 614 | 0 | 1 | 71-75 | PDAC | 0 | 2.4 |
| 615 | 1 | 0 | 66-70 | PDAC | 0 | 3 |
| 616 | 0 | 1 | 61-65 | PDAC | 1 | 2.8 |
| 617 | 0 | 1 | 71-75 | PDAC | 0 | 8 |
| 618 | 1 | 0 | 71-75 | PDAC | 0 | 2 |
| 619 | 1 | 0 | 71-75 | PDAC | 0 | 3.6 |
| 620 | 1 | 1 | 61-65 | PDAC | 0 | 2.7 |
| 621 | 0 | 0 | 61-65 | PDAC | 0 | 3.7 |
| 622 | 0 | 1 | 46-50 | PDAC | 1 | 5.8 |
| 623 | 1 | 0 | 76-80 | IPMN | 0 | 1.6 |
| 624 | 1 | 0 | 36-40 | PDAC | 0 | 3.5 |
| 625 | 1 | 1 | 71-75 | PDAC | 0 | 2.8 |
| 626 | 0 | 0 | 61-65 | PDAC | 0 | 4.4 |
| 627 | 0 | 1 | 56-60 | PDAC | 0 | 4.5 |
| 628 | 1 | 1 | 51-55 | PDAC | 1 | 3.9 |
| 629 | 1 | 1 | 71-75 | PDAC | 0 | 2.8 |
| 630 | 1 | 1 | 61-65 | PDAC | 1 | N/A |
| 631 | 0 | 0 | 71-75 | PDAC | 0 | 1.3 |
| 632 | 1 | 1 | 81-85 | PDAC | 1 | 3.7 |
| 633 | 1 | 1 | 71-75 | PDAC | 0 | 4 |
| 634 | 0 | 1 | 66-70 | PDAC | 0 | 4 |
| 635 | 1 | 1 | 61-65 | PDAC | 1 | 4.5 |
| 636 | 0 | 1 | 86-90 | PDAC | 0 | 1.5 |
| 637 | 0 | 0 | 66-70 | PDAC | 0 | 4.8 |
| 638 | 0 | 1 | 81-85 | PDAC | 0 | 2.2 |
| 639 | 1 | 1 | 61-65 | PDAC | 0 | 1.5 |
| 640 | 0 | 0 | 71-75 | PDAC | 0 | 3 |
| 641 | 1 | 0 | 66-70 | PDAC | 1 | 2.5 |
| 642 | 0 | 1 | 56-60 | PDAC | 0 | 2 |
| 643 | 0 | 1 | 56-60 | PDAC | 1 | 2.4 |
| 644 | 0 | 1 | 71-75 | PDAC | 0 | 4 |
| 645 | 1 | 1 | 56-60 | PDAC | 1 | 2.5 |
| 646 | 0 | 1 | 56-60 | PDAC | 1 | 4.5 |
| 647 | 1 | 0 | 56-60 | PDAC | 0 | 1.1 |
| 648 | 1 | 0 | 56-60 | PDAC | 0 | 3.1 |
| 649 | 0 | 1 | 76-80 | PDAC | 0 | 0.8 |
| 650 | 0 | 0 | 71-75 | PDAC | 0 | 1.1 |
| 651 | 0 | 1 | 46-50 | PDAC | 0 | 4 |
| 652 | 0 | 1 | 66-70 | PDAC | 1 | 4.5 |
| 653 | 1 | 1 | 56-60 | PDAC | 0 | 4.3 |
| 654 | 0 | 0 | 66-70 | PDAC | 0 | 4.8 |
| 655 | 0 | 1 | 56-60 | PDAC | 0 | 0.9 |
| 656 | 1 | 1 | 71-75 | PDAC | 1 | 1.5 |
| 657 | 0 | 0 | 71-75 | PDAC | 0 | 2.9 |
| 658 | 1 | 1 | 66-70 | PDAC | 0 | 2.5 |
| 659 | 1 | 0 | 66-70 | PDAC | 0 | 4.1 |
| 660 | 0 | 1 | 76-80 | PDAC | 1 | 2.1 |
| 661 | 0 | 1 | 66-70 | PDAC | 1 | 4 |
| 662 | 0 | 1 | 71-75 | PDAC | 0 | 2 |
| 663 | 0 | 0 | 81-85 | PDAC | 0 | 3.3 |
| 664 | 1 | 0 | 51-55 | PDAC | 0 | 3 |
| 665 | 1 | 0 | 71-75 | PDAC | 0 | 2.5 |
| 666 | 1 | 1 | 61-65 | PDAC | 1 | 3.5 |
| 667 | 1 | 0 | 71-75 | PDAC | 0 | 3.5 |
| 668 | 0 | 1 | 66-70 | PDAC | 0 | 4 |
| 669 | 1 | 0 | 66-70 | PDAC | 0 | 5.5 |
| 670 | 1 | 0 | 66-70 | PDAC | 0 | 2.5 |
| 671 | 1 | 1 | 46-50 | PDAC | 0 | 3.5 |
| 672 | 1 | 0 | 56-60 | PDAC | 0 | 3.1 |
| 673 | 1 | 1 | 76-80 | PDAC | 0 | 1.5 |
| 674 | 0 | 1 | 61-65 | PDAC | 0 | 2 |
| 675 | 0 | 1 | 61-65 | PDAC | 0 | 6.1 |
| 676 | 0 | 1 | 71-75 | PDAC | 1 | 4.8 |
| 677 | 1 | 1 | 81-85 | PDAC | 0 | 1.5 |

| | | | | | | |
|---|---|---|---|---|---|---|
| 678 | 0 | 1 | 66-70 | PDAC | 1 | 3.8 |
| 679 | 1 | 0 | 76-80 | PDAC | 0 | 1.5 |
| 680 | 1 | 0 | 66-70 | PDAC | 0 | 1.9 |
| 681 | 1 | 0 | 71-75 | PDAC | 0 | 2.1 |
| 682 | 1 | 1 | 51-55 | PDAC | 1 | 2.5 |
| 683 | 1 | 0 | 66-70 | PDAC | 0 | 5 |
| 684 | 0 | 0 | 71-75 | PDAC | 0 | 5.6 |
| 685 | 0 | 1 | 61-65 | PDAC | 0 | 3.5 |
| 686 | 1 | 1 | 66-70 | PDAC | 0 | 2.4 |
| 687 | 1 | 0 | 61-65 | PDAC | 0 | 1.6 |
| 688 | 1 | 0 | 66-70 | PDAC | 1 | 3.5 |
| 689 | 0 | 0 | 61-65 | PDAC, IPMN, PNET | 0 | 2.3 (PDAC), 0.5 (PNET) |
| 690 | 1 | 1 | 66-70 | PDAC | 0 | 2.8 |
| 691 | 0 | 1 | 61-65 | PDAC | 0 | 4.4 |
| 692 | 1 | 0 | 71-75 | PDAC | 0 | 3.5 |
| 693 | 1 | 0 | 61-65 | PDAC | 0 | 4.1 |
| 694 | 0 | 0 | 56-60 | PDAC | 0 | 5.6 |
| 695 | 0 | 1 | 66-70 | PDAC | 1 | 2..7 |
| 696 | 1 | 0 | 66-70 | PDAC | 0 | 4 |
| 697 | 1 | 1 | 41-45 | PDAC | 1 | 3.1 |
| 698 | 0 | 0 | 61-65 | PDAC | 1 | 3.1 |
| 699 | 0 | 1 | 66-70 | PDAC | 1 | 5 |
| 700 | 1 | 0 | 61-65 | PDAC | 1 | 3.9 |
| 701 | 0 | 0 | 61-65 | PDAC, PNET | 1 | 2(PDAC), 0.5 (PNET) |
| 702 | 1 | 0 | 66-70 | PDAC | 0 | 2.5 |
| 703 | 1 | 0 | 61-65 | PDAC | 0 | 2.6 |
| 704 | 0 | 0 | 61-65 | PDAC | 0 | 2.6 |
| 705 | 0 | 0 | 71-75 | PDAC | 1 | 4 |
| 706 | 1 | 0 | 36-40 | PDAC | 0 | 4.2 |
| 707 | 0 | 0 | 66-70 | PDAC | 0 | 9 |
| 708 | 0 | 0 | 46-50 | PDAC | 0 | 2 |
| 709 | 0 | 0 | 66-70 | PDAC | 0 | 6 |
| 710 | 1 | 0 | 61-65 | PDAC | 0 | 2.5 |
| 711 | 1 | 1 | 61-65 | PDAC | 0 | 3.5 |
| 712 | 1 | 0 | 71-75 | PDAC | 0 | 3 |
| 713 | 1 | 1 | 56-60 | PDAC | 0 | 5.4 |
| 714 | 0 | 0 | 66-70 | PDAC | 0 | 3.8 |
| 715 | 0 | 1 | 66-70 | PDAC | 0 | 6.5 |
| 716 | 1 | 0 | 56-60 | PDAC | 0 | 2.5 |
| 717 | 0 | 0 | 61-65 | PDAC | 0 | 5.5 |
| 718 | 0 | 0 | 76-80 | PDAC | 0 | 3.3 |
| 719 | 0 | 0 | 26-30 | PDAC | 1 | 3.5 |
| 720 | 0 | 1 | 66-70 | PDAC | 0 | 6 |
| 721 | 0 | 0 | 66-70 | PDAC | 0 | 0.6 |
| 722 | 1 | 0 | 41-45 | PDAC | 0 | 2.5 |
| 723 | 1 | 1 | 61-65 | PDAC | 1 | 2.7 |
| 724 | 1 | 1 | 46-50 | PDAC | 0 | 2.6 |
| 725 | 1 | 0 | 66-70 | PDAC | 0 | 1.5 |
| 726 | 0 | 0 | 36-40 | PDAC | 0 | 5.8 |
| 727 | 0 | 0 | 81-85 | PDAC | 0 | o pancreas but not arising directly from it) |
| 728 | 1 | 1 | 51-55 | PDAC | 1 | 2.5 |
| 729 | 1 | 1 | 56-60 | PDAC | 0 | 3 |
| 730 | 1 | 1 | 51-55 | PDAC | 0 | 1.2 |
| 731 | 0 | 0 | 66-70 | PDAC | 0 | 2.3 |
| 732 | 1 | 1 | 56-60 | PDAC | 1 | 4.8 |
| 733 | 0 | 0 | 46-50 | PDAC | 0 | 1.3 |
| 734 | 1 | 0 | 56-60 | PDAC | 1 | 2.2 |
| 735 | 0 | 1 | 76-80 | PDAC | 1 | 4.5 |
| 736 | 0 | 0 | 76-80 | PDAC | 1 | 2.6 |
| 737 | 0 | 0 | 76-80 | PDAC | 1 | 2.5 |
| 738 | 1 | 0 | 51-55 | PDAC | 1 | 4.4 |
| 739 | 1 | 1 | 81-85 | PDAC | 0 | 3.2 |
| 740 | 1 | 0 | 51-55 | PDAC | 1 | 1.5 |
| 741 | 0 | 0 | 71-75 | PDAC | 0 | 1.9 |
| 742 | 1 | 1 | 86-90 | PDAC | 1 | 2.8 |
| 743 | 1 | 0 | 51-55 | PDAC | 0 | 3.5 |
| 744 | 0 | 0 | 46-50 | PDAC | 1 | 1 |
| 745 | 1 | 0 | 56-60 | PDAC | 1 | 2.5 |
| 746 | 0 | 1 | 76-80 | PDAC | 0 | 6.8 |
| 747 | 0 | 1 | 76-80 | PDAC | 0 | 5.2 |
| 748 | 0 | 0 | 81-85 | PDAC | 0 | 3.2 |
| 749 | 0 | 0 | 56-60 | PDAC | 0 | 4.4 |
| 750 | 0 | 0 | 56-60 | PDAC | 0 | 2.6 |
| 751 | 1 | 0 | 66-70 | PDAC | 0 | 1.6 |
| 752 | 1 | 0 | 66-70 | PDAC | 1 | 2 |
| 753 | 0 | 0 | 71-75 | PDAC | 0 | 3.5 |
| 754 | 0 | 1 | 81-85 | PDAC | 1 | 5 |
| 755 | 0 | 0 | 66-70 | PDAC | 0 | 3.5 |
| 756 | 1 | 0 | 76-80 | PDAC | 0 | 2.3 |
| 757 | 0 | 0 | 71-75 | IPMN, PDAC | 0 | 2.1 |
| 758 | 0 | 1 | 61-65 | PDAC | 0 | 2.2 |
| 759 | 0 | 0 | 66-70 | PDAC | 1 | 2.6 |
| 760 | 0 | 0 | 76-80 | PDAC | 0 | 3.4 |
| 761 | 0 | 1 | 61-65 | PDAC | 1 | 2.6 |
| 762 | 1 | 0 | 56-60 | PDAC | 0 | 4 |
| 763 | 0 | 0 | 71-75 | PDAC | 1 | 5 |
| 764 | 1 | 0 | 86-90 | PDAC | 1 | 4.5 |
| 765 | 1 | 0 | 41-45 | | 0 | 2.4 |
| 766 | 0 | 0 | 71-75 | PDAC | 0 | 3 |
| 767 | 1 | 0 | 71-75 | PDAC | 0 | 3.4 |
| 768 | 0 | 0 | 71-75 | PDAC | 1 | 2.9 |
| 769 | 0 | 0 | 71-75 | PDAC | 1 | 3.7 |
| 770 | 1 | 1 | 56-60 | PDAC | 1 | 2.8 |
| 771 | 1 | 1 | 66-70 | PDAC | 1 | 2.1 |
| 772 | 1 | 1 | 66-70 | PDAC | 1 | 3 |
| 773 | 0 | 1 | 51-55 | PDAC | 0 | 2.3 |
| 774 | 0 | 0 | 66-70 | PDAC | 0 | 2.9 |

| | | | | | | |
|---|---|---|---|---|---|---|
| 775 | 1 | 1 | 71-75 | PDAC | 0 | 6 |
| 776 | 0 | 1 | 76-80 | PDAC | 0 | 3.3 |
| 777 | 0 | 0 | 66-70 | PDAC | 0 | 0.4 |
| 778 | 1 | 0 | 71-75 | PDAC | 0 | 3.1 |
| 779 | 1 | 0 | 71-75 | PDAC | 0 | 4.5 |
| 780 | 1 | 0 | 61-65 | PDAC | 0 | 5.6 |
| 781 | 1 | 0 | 71-75 | PDAC | 0 | 2.1 |
| 782 | 1 | 0 | 66-70 | PDAC | 0 | 3 |
| 783 | 1 | 0 | 61-65 | PDAC | 1 | 5 |
| 784 | 0 | 0 | 61-65 | PDAC | 1 | 3.5 |
| 785 | 0 | 1 | 76-80 | PDAC | 1 | 4.2 |
| 786 | 1 | 0 | 56-60 | PDAC | 0 | 5.1 |
| 787 | 0 | 0 | 81-85 | ed adenoneuroendocrine carcin | 0 | 2.6 |
| 788 | 1 | 1 | 61-65 | PDAC | 0 | 3.3 |
| 789 | 1 | 0 | 51-55 | PDAC | 0 | 2.5 |
| 790 | 0 | 1 | 66-70 | PDAC | 0 | 4.5 |
| 791 | 0 | 1 | 61-65 | PDAC | 0 | 0.3 |
| 792 | 1 | 1 | 61-65 | PDAC | 0 | 3.5 |
| 793 | 1 | 0 | 56-60 | PDAC | 0 | 2 |
| 794 | 0 | 1 | 61-65 | PDAC | 0 | 1 |
| 795 | 1 | 0 | 66-70 | PDAC | 1 | 3 |
| 796 | 0 | 0 | 71-75 | PDAC | 0 | 2.5 |
| 797 | 0 | 0 | 66-70 | PDAC | 0 | 2.3 |
| 798 | 1 | 0 | 71-75 | PDAC | 0 | 1.3 |
| 799 | 0 | 0 | 66-70 | PDAC | 0 | 2.8 |
| 800 | 1 | 0 | 61-65 | PDAC | 0 | 2.8 |
| 801 | 0 | 0 | 51-55 | PDAC | 0 | 1.8 |
| 802 | 0 | 0 | 76-80 | PDAC | 0 | 2.5 |
| 803 | 1 | 0 | 56-60 | PDAC | 1 | 2 |
| 804 | 1 | 1 | 76-80 | PDAC | 0 | 5.1 |
| 805 | 0 | 0 | 81-85 | PDAC | 1 | 2.2 |
| 806 | 0 | 0 | 56-60 | PDAC | 1 | 2.7 |
| 807 | 0 | 1 | 46-50 | PDAC | 0 | 1.2 |
| 808 | 1 | 1 | 71-75 | PDAC | 0 | 4.5 |
| 809 | 0 | 0 | 51-55 | PDAC | 1 | 3.2 |
| 810 | 1 | 1 | 41-45 | PDAC | 0 | 3 |
| 811 | 1 | 0 | 71-75 | PDAC | 1 | 1.5 |
| 812 | 0 | 0 | 71-75 | PDAC | 0 | 2.2 |
| 813 | 0 | 0 | 71-75 | PDAC | 0 | 2.9 |
| 814 | 1 | 0 | 61-65 | PDAC | 0 | 5.5 |
| 815 | 0 | 1 | 46-50 | PDAC | 0 | 2.5 |
| 816 | 1 | 0 | 66-70 | PDAC | 0 | 3 |
| 817 | 0 | 0 | 71-75 | PDAC | 1 | 5 |
| 818 | 1 | 0 | 66-70 | PDAC | 1 | 3 |
| 819 | 1 | 0 | 56-60 | PDAC | 1 | 2.3 |
| 820 | 0 | 0 | 76-80 | PDAC | 0 | 6 |
| 821 | 0 | 0 | 56-60 | PDAC | 1 | 3 |
| 822 | 1 | 0 | 76-80 | PDAC | 0 | 3 |
| 823 | 1 | 0 | 56-60 | PDAC | 0 | 1.2 |
| 824 | 0 | 0 | 46-50 | PDAC | 0 | 3.5 |
| 825 | 0 | 0 | 51-55 | PDAC | 1 | 3.2 |
| 826 | 0 | 1 | 71-75 | PDAC | 0 | 3.5 |
| 827 | 1 | 1 | 56-60 | PDAC | 0 | 2 |
| 828 | 1 | 0 | 66-70 | PDAC | 0 | 7.9 |
| 829 | 0 | 1 | 56-60 | PDAC | 1 | 3.2 |
| 830 | 1 | 1 | 71-75 | PDAC | 0 | 2.8 |
| 831 | 0 | 1 | 41-45 | PDAC | 1 | 2.5 |
| 832 | 0 | 1 | 51-55 | PDAC | 1 | 3 |
| 833 | 0 | 1 | 71-75 | PDAC | 0 | 4.5 |
| 834 | 0 | 0 | 56-60 | PDAC | 1 | 5.5 |
| 835 | 1 | 1 | 56-60 | PDAC | 0 | 3.9 |
| 836 | 0 | 0 | 56-60 | PDAC | 1 | 1.5 |
| 837 | 0 | 1 | 46-50 | PDAC | 1 | 3.5 |
| 838 | 0 | 0 | 56-60 | PDAC | 1 | 4.7 |
| 839 | 1 | 0 | 61-65 | PDAC | 1 | <1mm |
| 840 | 1 | 0 | 51-55 | PDAC | 1 | 2 |
| 841 | 1 | 0 | 81-85 | PDAC | 0 | 3.4 |
| 842 | 1 | 0 | 61-65 | PDAC | 1 | 2 |
| 843 | 1 | 1 | 51-55 | PDAC | 0 | 2.5 |
| 844 | 1 | 0 | 61-65 | | 1 | 3 |
| 845 | 1 | 1 | 56-60 | PDAC | 0 | 5.2 |
| 846 | 0 | 0 | 61-65 | PDAC | 0 | 2.1 |
| 847 | 0 | 1 | 56-60 | PDAC | 0 | 2.7 |
| 848 | 0 | 0 | 51-55 | PDAC | 1 | 3.8 |
| 849 | 0 | 1 | 61-65 | PDAC | 0 | 3 |
| 850 | 1 | 0 | 66-70 | PDAC | 0 | 2 |
| 851 | 1 | 1 | 71-75 | PDAC | 0 | 3 |
| 852 | 1 | 0 | 76-80 | | 0 | 1.7 |
| 853 | 1 | 0 | 56-60 | PDAC | 1 | 4.5 |
| 854 | 1 | 1 | 71-75 | PDAC | 0 | 3.4 |
| 855 | 1 | 1 | 81-85 | PDAC | 0 | 4 |
| 856 | 0 | 0 | 41-45 | PDAC | 1 | 3.2 |
| 857 | 1 | 1 | 66-70 | PDAC | 0 | 3.8 |
| 858 | 0 | 0 | 66-70 | PDAC | 0 | 3.5 |
| 859 | 1 | 0 | 61-65 | PDAC | 0 | 4 |
| 860 | 1 | 0 | 71-75 | PDAC | 0 | 3.6 |
| 861 | 0 | 1 | 66-70 | PDAC | 0 | 4.2 |
| 862 | 1 | 0 | 56-60 | PDAC | 0 | 4 |
| 863 | 1 | 0 | 61-65 | PDAC | 0 | 1.4 |
| 864 | 0 | 0 | 76-80 | PDAC | 1 | 5 |
| 865 | 0 | 0 | 66-70 | PDAC | 1 | 5 |
| 866 | 1 | 1 | 76-80 | PDAC | 0 | 2 |
| 867 | 1 | 1 | 51-55 | PDAC | 1 | 2.7 |
| 868 | 1 | 0 | 71-75 | PDAC | 1 | 2 |
| 869 | 0 | 1 | 56-60 | PDAC | 0 | 4 |
| 870 | 0 | 0 | 76-80 | PDAC | 0 | 5.4 |
| 871 | 1 | 0 | 51-55 | PDAC | 0 | 1.4 |

| | | | | | | |
|---|---|---|---|---|---|---|
| 872 | 1 | 0 | 66-70 | PDAC | 1 | 4.5 |
| 873 | 0 | 0 | 66-70 | PDAC | 0 | Multiple <2cm |
| 874 | 0 | 0 | 66-70 | PDAC | 0 | 3.2 |
| 875 | 0 | 0 | 61-65 | PDAC | 0 | 6 |
| 876 | 0 | 0 | 76-80 | PDAC | 1 | 1.2 |
| 877 | 1 | 0 | 71-75 | PDAC | 1 | 2.8 |
| 878 | 0 | 0 | 51-55 | PDAC | 1 | 3.8 |
| 879 | 0 | 0 | 61-65 | PDAC | 0 | 2.1 |
| 880 | 0 | 0 | 56-60 | PDAC | 1 | 3.4 |
| 881 | 0 | 0 | 51-55 | PDAC | 1 | 7 |
| 882 | 0 | 0 | 76-80 | PDAC | 0 | 2.9 |
| 883 | 0 | 0 | 51-55 | PDAC | 1 | 3 |
| 884 | 0 | 0 | 76-80 | PDAC | 0 | 2.4 |
| 885 | 1 | 0 | 46-50 | PDAC | 1 | 3.5 |
| 886 | 1 | 1 | 76-80 | PDAC | 1 | 5.5 |
| 887 | 0 | 0 | 71-75 | PDAC | 1 | 4.1 |
| 888 | 1 | 1 | 66-70 | PDAC | 1 | 2.2 |
| 889 | 0 | 1 | 61-65 | PDAC | 0 | 4.9 |
| 890 | 0 | 1 | 76-80 | PDAC | 1 | 3 |
| 891 | 0 | 0 | 71-75 | PDAC | 1 | 5.6 |
| 892 | 0 | 0 | 41-45 | PDAC | 0 | 4.7 |
| 893 | 1 | 0 | 61-65 | PDAC | 0 | 3.5 |
| 894 | 0 | 1 | 71-75 | PDAC | 0 | 2.8 |
| 895 | 0 | 0 | 71-75 | PDAC | 1 | 2 |
| 896 | 1 | 1 | 41-45 | PDAC | 1 | 3.5 |
| 897 | 0 | 0 | 61-65 | PDAC | 0 | 3.4 |
| 898 | 1 | 0 | 56-60 | PDAC | 0 | 0.2 |
| 899 | 0 | 1 | 61-65 | PDAC | 1 | 3.5 |
| 900 | 0 | 0 | 71-75 | PDAC | 0 | 3.8 |
| 901 | 1 | 1 | 46-50 | PDAC | 1 | 2.3 |
| 902 | 0 | 0 | 56-60 | PDAC | 0 | 2.3 |
| 903 | 0 | 0 | 66-70 | PDAC | 0 | 4 |
| 904 | 1 | 1 | 51-55 | PDAC | 0 | 5 |
| 905 | 0 | 1 | 71-75 | PDAC | 0 | 1.3 |
| 906 | 0 | 0 | 66-70 | PDAC | 0 | 1.5 |
| 907 | 1 | 0 | 71-75 | PDAC | 0 | 3.5 |
| 908 | 0 | 1 | 66-70 | PDAC | 0 | 3.8 |
| 909 | 1 | 0 | 76-80 | PDAC | (2/24/2014), 1 (5/19/201 | 2.8 |
| 910 | 1 | 1 | 71-75 | PDAC | 0 | 1.7 |
| 911 | 0 | 0 | 56-60 | PDAC | 0 | 1.5 |
| 912 | 0 | 0 | 71-75 | PDAC | 0 | 2.6 |
| 913 | 0 | 0 | 61-65 | PDAC | 1 | 2.3 |
| 914 | 0 | 1 | 71-75 | PDAC | 1 | 1.4 |
| 915 | 0 | 0 | 71-75 | PDAC | 0 | 5.9 |
| 916 | 1 | 0 | 71-75 | PDAC | 0 | 2.1 |
| 917 | 1 | 0 | 76-80 | PDAC | 0 | 2 |
| 918 | 0 | 1 | 61-65 | PDAC | 1 | 1.2 |
| 919 | 0 | 0 | 56-60 | PDAC | 0 | 4 |
| 920 | 1 | 1 | 81-85 | PDAC | 0 | 1.8 |
| 921 | 1 | 0 | 51-55 | PDAC | 0 | 4.5 |
| 922 | 1 | 0 | 61-65 | PDAC | 0 | 3.5 |
| 923 | 0 | 1 | 71-75 | PDAC | 0 | 2.1 |
| 924 | 0 | 0 | 41-45 | PDAC | 0 | 2.5 |
| 925 | 1 | 0 | 66-70 | PDAC | 0 | 2.3 |
| 926 | 1 | 1 | 66-70 | PDAC | 1 | 4.7 |
| 927 | 1 | 0 | 51-55 | PDAC | 0 | 4.4 |
| 928 | 0 | 0 | 71-75 | PDAC | 1 | 3.5 |
| 929 | 0 | 0 | 46-50 | PDAC | 0 | 1.7 |
| 930 | 0 | 0 | 51-55 | PDAC | 1 | 4.8 |
| 931 | 0 | 1 | 66-70 | PDAC | 0 | 2.4 |
| 932 | 0 | 0 | 76-80 | PDAC | 0 | 3.4 |
| 933 | 0 | 1 | 71-75 | PanIN II | 0 | 1.9 |
| 934 | 0 | 0 | 66-70 | PDAC | 0 | 2.5 |
| 935 | 0 | 0 | 76-80 | PDAC | 0 | 6.8 |
| 936 | 1 | 0 | 71-75 | PDAC | 0 | 2 |
| 937 | 0 | 1 | 76-80 | PDAC | 1 | 1.5 |
| 938 | 0 | 0 | 71-75 | PDAC | 0 | 2.4 |
| 939 | 0 | 1 | 66-70 | PDAC | 0 | 4 |
| 940 | 0 | 0 | 86-90 | PDAC | 0 | 7.9 |
| 941 | 0 | 0 | 46-50 | PDAC | 1 | 3.1 |
| 942 | 1 | 1 | 61-65 | PDAC | 0 | 5.1 |
| 943 | 0 | 1 | 36-40 | PDAC | 0 | 2.6 |
| 944 | 0 | 0 | 46-50 | PDAC | 1 | 3.4 |
| 945 | 0 | 0 | 66-70 | PDAC | 1 | 2.9 |
| 946 | 1 | 0 | 81-85 | PDAC | 1 | 2.7 |
| 947 | 1 | 0 | 61-65 | PDAC | 0 | 3.6 |
| 948 | 1 | 1 | 76-80 | PDAC | 0 | 4.6 |
| 949 | 0 | 0 | 61-65 | PDAC | 0 | 7.7 |
| 950 | 1 | 0 | 61-65 | PDAC | 0 | 6.4 |
| 951 | 1 | 0 | 66-70 | PDAC | 0 | 3 |
| 952 | 1 | 0 | 51-55 | PDAC | 0 | 1 |
| 953 | 1 | 1 | 56-60 | PDAC | 0 | 2.9 |
| 954 | 1 | 1 | 61-65 | PDAC | 1 | 2.9 |
| 955 | 0 | 0 | 71-75 | PDAC | 0 | 3.2 |
| 956 | 1 | 0 | 56-60 | PDAC | 1 | 2.5 |
| 957 | 1 | 1 | 81-85 | PDAC | 1 | 2.5 |
| 958 | 1 | 0 | 51-55 | PDAC | 0 | 1.5 |
| 959 | 1 | 0 | 61-65 | PDAC | 1 | 3.5 |
| 960 | 0 | 1 | 61-65 | PDAC | 1 | 4 |
| 961 | 0 | 0 | 76-80 | PDAC | 0 | 2.5 |
| 962 | 1 | 1 | 66-70 | PDAC | 0 | 4.5 |
| 963 | 1 | 1 | 61-65 | PDAC | 0 | 5.2 |
| 964 | 0 | 1 | 61-65 | PDAC | 0 | 2.5 |
| 965 | 0 | 0 | 51-55 | PDAC | 1 | 3 |
| 966 | 1 | 0 | 61-65 | PDAC | 1 | 2.8 |
| 967 | 1 | 1 | 56-60 | PDAC | 0 | 1.9 |
| 968 | 0 | 1 | 66-70 | PDAC | 0 | 3.6 |

| | | | | | | |
|---|---|---|---|---|---|---|
| 969 | 0 | 0 | 76-80 | PDAC | 1 | 2.2 |
| 970 | 1 | 1 | 56-60 | PDAC | 1 | 3 |
| 971 | 0 | 1 | 71-75 | PDAC | 0 | 2.4 |
| 972 | 0 | 0 | 61-65 | PDAC | 1 | 2.5 |
| 973 | 1 | 0 | 81-85 | PDAC | 0 | 5.8 |
| 974 | 1 | 1 | 81-85 | PDAC | 0 | 7.6 |
| 975 | 0 | 1 | 41-45 | PDAC | 1 | 2 |
| 976 | 1 | 0 | 66-70 | PDAC | 0 | 4.9 |
| 977 | 1 | 0 | 56-60 | PDAC | 1 | 1.3 |
| 978 | 1 | 0 | 56-60 | PDAC | 0 | 4.5 |
| 979 | 0 | 0 | 76-80 | PDAC | 0 | 2.4 |
| 980 | 0 | 0 | 71-75 | PDAC | 0 | 6.1 |
| 981 | 1 | 0 | 66-70 | PDAC | 0 | 0.4 |
| 982 | 1 | 0 | 71-75 | PDAC | 0 | 3.2 |
| 983 | 1 | 1 | 51-55 | PDAC | 0 | 2.5 |
| 984 | 0 | 1 | 81-85 | PDAC | 0 | 3.8 |
| 985 | 1 | 0 | 56-60 | PDAC | 1 | 2.8 |
| 986 | 0 | 0 | 71-75 | PDAC | 0 | 2.9 |
| 987 | 1 | 1 | 61-65 | PDAC | 0 | 3.1 |
| 988 | 0 | 1 | 76-80 | PDAC | 0 | 4.5 |
| 989 | 1 | 1 | 71-75 | PDAC | 1 | 3 |
| 990 | 0 | 0 | 66-70 | PDAC | 1 | 2 |
| 991 | 0 | 1 | 66-70 | PDAC | 1 | 2.6 |
| 992 | 1 | 0 | 81-85 | PDAC | 1 | 1.5 |
| 993 | 1 | 1 | 71-75 | PDAC | 0 | 3.4 |
| 994 | 0 | 1 | 81-85 | PDAC | 0 | 3 |
| 995 | 1 | 1 | 56-60 | PDAC | 0 | 2.3 |
| 996 | 0 | 0 | 61-65 | PDAC | 1 | 4 |
| 997 | 1 | 1 | 66-70 | PDAC | 0 | 2.8 |
| 998 | 1 | 0 | 56-60 | PDAC | 0 | 2 |
| 999 | 1 | 0 | 61-65 | PDAC | 0 | 3 |
| 1000 | 0 | 1 | 56-60 | PDAC | 0 | 4 |
| 1001 | 1 | 1 | 56-60 | PDAC | 0 | 3 |
| 1002 | 1 | 0 | 76-80 | PDAC | 1 | 3.9 |
| 1003 | 0 | 1 | 56-60 | PDAC | 1 | 5.5 |
| 1004 | 0 | 1 | 61-65 | PDAC | 0 | 9.8 |
| 1005 | 1 | 0 | 71-75 | PDAC | 1 (7/31/2017) | 2.1 |
| 1006 | 0 | 0 | 56-60 | None | 0 | 3.6 |
| 1007 | 0 | 0 | 66-70 | PDAC | 0 | 2.2 |
| 1008 | 0 | 0 | 61-65 | PDAC | 0 | 1.3 |
| 1009 | 1 | 0 | 61-65 | PDAC | 0 | 3.2 |
| 1010 | 0 | 0 | 56-60 | PDAC | 0 | 5 |
| 1011 | 0 | 0 | 51-55 | PDAC | 0 | 3.7 |
| 1012 | 0 | 0 | 51-55 | PDAC | 0 | 4.7 |
| 1013 | 1 | 0 | 46-50 | PDAC | 1 | 3.1 |
| 1014 | 1 | 1 | 66-70 | PDAC | 0 | 1.5 |
| 1015 | 1 | 1 | 71-75 | PDAC | 0 | 2.5 |
| 1016 | 1 | 0 | 81-85 | PDAC | 0 | 2.5 |
| 1017 | 0 | 0 | 71-75 | PDAC | 0 | 2.5 |
| 1018 | 1 | 0 | 26-30 | RENAL DONOR | | |
| 1019 | 1 | 0 | 51-55 | RENAL DONOR | | |
| 1020 | 1 | 0 | 51-55 | RENAL DONOR | | |
| 1021 | 1 | 0 | 51-55 | RENAL DONOR | | |
| 1022 | 1 | 0 | 46-50 | RENAL DONOR | | |
| 1023 | 1 | 0 | 56-60 | RENAL DONOR | | |
| 1024 | 1 | 0 | 51-55 | RENAL DONOR | | |
| 1025 | 0 | 0 | 51-55 | RENAL DONOR | | |
| 1026 | 1 | 0 | 56-60 | RENAL DONOR | | |
| 1027 | 1 | 0 | 61-65 | RENAL DONOR | | |
| 1028 | 0 | 0 | 61-65 | RENAL DONOR | | |
| 1029 | 1 | 0 | 41-45 | RENAL DONOR | | |
| 1030 | 1 | 0 | 46-50 | RENAL DONOR | | |
| 1031 | 1 | 0 | 46-50 | RENAL DONOR | | |
| 1032 | 1 | 0 | 71-75 | RENAL DONOR | | |
| 1033 | 0 | 0 | 46-50 | RENAL DONOR | | |
| 1034 | 1 | 0 | 31-35 | RENAL DONOR | | |
| 1035 | 1 | 0 | 56-60 | RENAL DONOR | | |
| 1036 | 1 | 0 | 51-55 | RENAL DONOR | | |
| 1037 | 1 | 0 | 46-50 | RENAL DONOR | | |
| 1038 | 1 | 0 | 26-30 | RENAL DONOR | | |
| 1039 | 0 | 0 | 41-45 | RENAL DONOR | | |
| 1040 | 0 | 0 | 41-45 | RENAL DONOR | | |
| 1041 | 0 | 0 | 51-55 | RENAL DONOR | | |
| 1042 | 0 | 0 | 66-70 | RENAL DONOR | | |
| 1043 | 1 | 0 | 61-65 | RENAL DONOR | | |
| 1044 | 1 | 0 | 56-60 | RENAL DONOR | | |
| 1045 | 1 | 0 | 26-30 | RENAL DONOR | | |
| 1046 | 1 | 0 | 46-50 | RENAL DONOR | | |
| 1047 | 1 | 0 | 41-45 | RENAL DONOR | | |
| 1048 | 1 | 0 | 41-45 | RENAL DONOR | | |
| 1049 | 1 | 0 | 46-50 | RENAL DONOR | | |
| 1050 | 1 | 0 | 41-45 | RENAL DONOR | | |
| 1051 | | | | | | |
| 1052 | 1 | 0 | 26-30 | RENAL DONOR | | |
| 1053 | 1 | 0 | 36-40 | RENAL DONOR | | |
| 1054 | 0 | 0 | 21-25 | RENAL DONOR | | |
| 1055 | 1 | 0 | 51-55 | RENAL DONOR | | |
| 1056 | 0 | 0 | 36-40 | RENAL DONOR | | |
| 1057 | 1 | 0 | 41-45 | RENAL DONOR | | |
| 1058 | 0 | 0 | 21-25 | RENAL DONOR | | |
| 1059 | 1 | 0 | 46-50 | RENAL DONOR | | |
| 1060 | 0 | 0 | 46-50 | RENAL DONOR | | |
| 1061 | 1 | 0 | 36-40 | RENAL DONOR | | |
| 1062 | 1 | 0 | 31-35 | RENAL DONOR | | |
| 1063 | 1 | 0 | 31-35 | RENAL DONOR | | |
| 1064 | 1 | 0 | 26-30 | RENAL DONOR | | |
| 1065 | 1 | 0 | 36-40 | RENAL DONOR | | |

| | | | | |
|---|---|---|---|---|
| 1066 | 0 | 0 | 41-45 | RENAL DONOR |
| 1067 | 0 | 0 | 36-40 | RENAL DONOR |
| 1068 | 1 | 0 | 26-30 | RENAL DONOR |
| 1069 | 0 | 0 | 31-35 | RENAL DONOR |
| 1070 | 1 | 0 | 36-40 | RENAL DONOR |
| 1071 | 1 | 0 | 31-35 | RENAL DONOR |
| 1072 | 1 | 0 | 41-45 | RENAL DONOR |
| 1073 | 1 | 0 | 36-40 | RENAL DONOR |
| 1074 | 0 | 0 | 46-50 | RENAL DONOR |
| 1075 | 0 | 0 | 26-30 | RENAL DONOR |
| 1076 | 0 | 0 | 41-45 | RENAL DONOR |
| 1077 | 0 | 0 | 36-40 | RENAL DONOR |
| 1078 | 0 | 0 | 71-75 | RENAL DONOR |
| 1079 | 0 | 0 | 51-55 | RENAL DONOR |
| 1080 | 1 | 0 | 36-40 | RENAL DONOR |
| 1081 | 1 | 0 | 26-30 | RENAL DONOR |
| 1082 | 1 | 0 | 21-25 | RENAL DONOR |
| 1083 | 1 | 0 | 21-25 | RENAL DONOR |
| 1084 | 1 | 0 | 21-25 | RENAL DONOR |
| 1085 | 1 | 0 | 21-25 | RENAL DONOR |
| 1086 | 0 | 0 | 16-20 | RENAL DONOR |
| 1087 | 1 | 0 | 21-25 | RENAL DONOR |
| 1088 | 0 | 0 | 31-35 | RENAL DONOR |
| 1089 | 1 | 0 | 46-50 | RENAL DONOR |
| 1090 | 1 | 0 | 41-45 | RENAL DONOR |
| 1091 | 1 | 0 | 26-30 | RENAL DONOR |
| 1092 | 0 | 0 | 36-40 | RENAL DONOR |
| 1093 | 0 | 0 | 41-45 | RENAL DONOR |
| 1094 | 0 | 0 | 51-55 | RENAL DONOR |
| 1095 | 0 | 0 | 26-30 | RENAL DONOR |
| 1096 | 0 | 0 | 51-55 | RENAL DONOR |
| 1097 | 1 | 0 | 36-40 | RENAL DONOR |
| 1098 | 1 | 0 | 76-80 | RENAL DONOR |
| 1099 | 1 | 0 | 36-40 | RENAL DONOR |
| 1100 | 1 | 0 | 26-30 | RENAL DONOR |
| 1101 | 0 | 0 | 31-35 | RENAL DONOR |
| 1102 | 0 | 0 | 61-65 | RENAL DONOR |
| 1103 | 0 | 0 | 51-55 | RENAL DONOR |
| 1104 | 0 | | 31-35 | RENAL DONOR |
| 1105 | 1 | 0 | 31-35 | RENAL DONOR |
| 1106 | 1 | 0 | 46-50 | RENAL DONOR |
| 1107 | 1 | 0 | 46-50 | RENAL DONOR |
| 1108 | 1 | 0 | 36-40 | RENAL DONOR |
| 1109 | 0 | 0 | 41-45 | RENAL DONOR |
| 1110 | 0 | 0 | 41-45 | RENAL DONOR |
| 1111 | 1 | 0 | 56-60 | RENAL DONOR |
| 1112 | 0 | 0 | 36-40 | RENAL DONOR |
| 1113 | 0 | 0 | 26-30 | RENAL DONOR |
| 1114 | 1 | 0 | 36-40 | RENAL DONOR |
| 1115 | 0 | 0 | 46-50 | RENAL DONOR |
| 1116 | 1 | 0 | 51-55 | RENAL DONOR |
| 1117 | 0 | 0 | 41-45 | RENAL DONOR |
| 1118 | 1 | 0 | 36-40 | RENAL DONOR |
| 1119 | 1 | 0 | 36-40 | RENAL DONOR |
| 1120 | 0 | 0 | 46-50 | RENAL DONOR |
| 1121 | 0 | 0 | 36-40 | RENAL DONOR |
| 1122 | 1 | 0 | 31-35 | RENAL DONOR |
| 1123 | 1 | 0 | 51-55 | RENAL DONOR |
| 1124 | 1 | 0 | 41-45 | RENAL DONOR |
| 1125 | 1 | 0 | 56-60 | RENAL DONOR |
| 1126 | 1 | 0 | 36-40 | RENAL DONOR |
| 1127 | 0 | 0 | 46-50 | RENAL DONOR |
| 1128 | 1 | 0 | 61-65 | RENAL DONOR |
| 1129 | 1 | 0 | 36-40 | RENAL DONOR |
| 1130 | 1 | 0 | 51-55 | RENAL DONOR |
| 1131 | 0 | 0 | 31-35 | RENAL DONOR |
| 1132 | 1 | 0 | 56-60 | RENAL DONOR |
| 1133 | 1 | 0 | 31-35 | RENAL DONOR |
| 1134 | 0 | 0 | 41-45 | RENAL DONOR |
| 1135 | 0 | 0 | 41-45 | RENAL DONOR |
| 1136 | 0 | 0 | 26-30 | RENAL DONOR |
| 1137 | 0 | 0 | 51-55 | RENAL DONOR |
| 1138 | 0 | 0 | 56-60 | RENAL DONOR |
| 1139 | 1 | 0 | 51-55 | RENAL DONOR |
| 1140 | 1 | 0 | 41-45 | RENAL DONOR |
| 1141 | 1 | 0 | 41-45 | RENAL DONOR |
| 1142 | 0 | 0 | 46-50 | RENAL DONOR |
| 1143 | 1 | 0 | 46-50 | RENAL DONOR |
| 1144 | 0 | 0 | 21-25 | RENAL DONOR |
| 1145 | 1 | 0 | 26-30 | RENAL DONOR |
| 1146 | 1 | 0 | 51-55 | RENAL DONOR |
| 1147 | 0 | 0 | 36-40 | RENAL DONOR |
| 1148 | 1 | 0 | 46-50 | RENAL DONOR |
| 1149 | 1 | 0 | 46-50 | RENAL DONOR |
| 1150 | 1 | 0 | 41-45 | RENAL DONOR |
| 1151 | 0 | 0 | 51-55 | RENAL DONOR |
| 1152 | 0 | 0 | 41-45 | RENAL DONOR |
| 1153 | 0 | 0 | 46-50 | RENAL DONOR |
| 1154 | 1 | 0 | 61-65 | RENAL DONOR |
| 1155 | 0 | 0 | 51-55 | RENAL DONOR |
| 1156 | 1 | 0 | 36-40 | RENAL DONOR |
| 1157 | 0 | 0 | 36-40 | RENAL DONOR |
| 1158 | 0 | 0 | 51-55 | RENAL DONOR |
| 1159 | 1 | 0 | 36-40 | RENAL DONOR |
| 1160 | 1 | 0 | 31-35 | RENAL DONOR |

| | | | | |
|---|---|---|---|---|
| 1161 | 1 | 0 | 46-50 | RENAL DONOR |
| 1162 | 1 | 0 | 26-30 | RENAL DONOR |
| 1163 | 0 | 0 | 51-55 | RENAL DONOR |
| 1164 | 0 | 0 | 41-45 | RENAL DONOR |
| 1165 | 0 | 0 | 26-30 | RENAL DONOR |
| 1166 | 0 | 0 | 41-45 | RENAL DONOR |
| 1167 | 1 | 0 | 36-40 | RENAL DONOR |
| 1168 | 0 | 0 | 26-30 | RENAL DONOR |
| 1169 | 0 | 0 | 31-35 | RENAL DONOR |
| 1170 | 1 | 0 | 56-60 | RENAL DONOR |
| 1171 | 1 | 0 | 51-55 | RENAL DONOR |
| 1172 | 1 | 0 | 51-55 | RENAL DONOR |
| 1173 | 1 | 0 | 21-25 | RENAL DONOR |
| 1174 | 1 | 0 | 41-45 | RENAL DONOR |
| 1175 | 1 | 0 | 46-50 | RENAL DONOR |
| 1176 | 0 | 0 | 26-30 | RENAL DONOR |
| 1177 | 1 | 0 | 71-75 | RENAL DONOR |
| 1178 | 0 | 0 | 26-30 | RENAL DONOR |
| 1179 | 0 | 0 | 26-30 | RENAL DONOR |
| 1180 | 0 | 0 | 36-40 | RENAL DONOR |
| 1181 | 0 | 0 | 51-55 | RENAL DONOR |
| 1182 | 1 | 0 | 36-40 | RENAL DONOR |
| 1183 | 1 | 0 | 41-45 | RENAL DONOR |
| 1184 | 1 | 0 | 46-50 | RENAL DONOR |
| 1185 | 1 | 0 | 36-40 | RENAL DONOR |
| 1186 | 1 | 0 | 51-55 | RENAL DONOR |
| 1187 | 0 | 0 | 56-60 | RENAL DONOR |
| 1188 | 1 | 0 | 56-60 | RENAL DONOR |
| 1189 | 1 | 0 | 61-65 | RENAL DONOR |
| 1190 | 0 | 0 | 61-65 | RENAL DONOR |
| 1191 | 1 | 0 | 46-50 | RENAL DONOR |
| 1192 | 1 | 0 | 46-50 | RENAL DONOR |
| 1193 | 1 | 0 | 46-50 | RENAL DONOR |
| 1194 | 1 | 0 | 51-55 | RENAL DONOR |
| 1195 | 1 | 0 | 21-25 | RENAL DONOR |
| 1196 | 1 | 0 | 61-65 | RENAL DONOR |
| 1197 | 0 | 0 | 36-40 | RENAL DONOR |
| 1198 | 1 | 0 | 56-60 | RENAL DONOR |
| 1199 | 0 | 0 | 31-35 | RENAL DONOR |
| 1200 | 1 | 0 | 46-50 | RENAL DONOR |
| 1201 | 1 | 0 | 56-60 | RENAL DONOR |
| 1202 | 0 | 0 | 51-55 | RENAL DONOR |
| 1203 | 1 | 0 | 36-40 | RENAL DONOR |
| 1204 | 1 | 0 | 51-55 | RENAL DONOR |
| 1205 | 1 | 0 | 66-70 | RENAL DONOR |
| 1206 | 1 | 0 | 21-25 | RENAL DONOR |
| 1207 | 1 | 0 | 51-55 | RENAL DONOR |
| 1208 | 1 | 0 | 21-25 | RENAL DONOR |
| 1209 | 1 | 0 | 41-45 | RENAL DONOR |
| 1210 | 0 | 0 | 36-40 | RENAL DONOR |
| 1211 | 1 | 0 | 61-65 | RENAL DONOR |
| 1212 | 1 | 0 | 36-40 | RENAL DONOR |
| 1213 | 0 | 0 | 46-50 | RENAL DONOR |
| 1214 | 0 | 0 | 46-50 | RENAL DONOR |
| 1215 | 0 | 0 | 36-40 | RENAL DONOR |
| 1216 | 1 | 0 | 61-65 | RENAL DONOR |
| 1217 | 1 | 0 | 41-45 | RENAL DONOR |
| 1218 | 0 | 0 | 36-40 | RENAL DONOR |
| 1219 | 1 | 0 | 41-45 | RENAL DONOR |
| 1220 | 1 | 0 | 46-50 | RENAL DONOR |
| 1221 | 0 | 0 | 31-35 | RENAL DONOR |
| 1222 | 1 | 0 | 56-60 | RENAL DONOR |
| 1223 | 1 | 0 | 41-45 | RENAL DONOR |
| 1224 | 0 | 0 | 31-35 | RENAL DONOR |
| 1225 | 1 | 0 | 41-45 | RENAL DONOR |
| 1226 | 0 | 0 | 41-45 | RENAL DONOR |
| 1227 | 0 | 0 | 31-35 | RENAL DONOR |
| 1228 | 0 | 0 | 51-55 | RENAL DONOR |
| 1229 | 1 | 0 | 41-45 | RENAL DONOR |
| 1230 | 0 | 0 | 46-50 | RENAL DONOR |
| 1231 | 1 | 0 | 36-40 | RENAL DONOR |
| 1232 | 1 | 0 | 36-40 | RENAL DONOR |
| 1233 | 1 | 0 | 51-55 | RENAL DONOR |
| 1234 | 1 | 0 | 46-50 | RENAL DONOR |
| 1235 | 0 | 0 | 51-55 | RENAL DONOR |
| 1236 | 0 | 0 | 36-40 | RENAL DONOR |
| 1237 | 1 | 0 | 31-35 | RENAL DONOR |
| 1238 | 1 | 0 | 31-35 | RENAL DONOR |
| 1239 | 1 | 0 | 41-45 | RENAL DONOR |
| 1240 | 1 | 0 | 41-45 | RENAL DONOR |
| 1241 | 1 | 0 | 51-55 | RENAL DONOR |
| 1242 | 0 | 0 | 61-65 | RENAL DONOR |
| 1243 | 0 | 0 | 61-65 | RENAL DONOR |
| 1244 | 0 | 0 | 16-20 | RENAL DONOR |
| 1245 | 0 | 0 | 51-55 | RENAL DONOR |
| 1246 | 1 | 0 | 46-50 | RENAL DONOR |
| 1247 | 0 | 0 | 36-40 | RENAL DONOR |
| 1248 | 1 | 0 | 51-55 | RENAL DONOR |
| 1249 | 1 | 0 | 51-55 | RENAL DONOR |
| 1250 | 0 | 0 | 21-25 | RENAL DONOR |
| 1251 | 0 | 0 | 31-35 | RENAL DONOR |
| 1252 | 0 | 0 | 41-45 | RENAL DONOR |
| 1253 | 0 | 0 | 56-60 | RENAL DONOR |
| 1254 | 1 | 0 | 36-40 | RENAL DONOR |
| 1255 | 0 | 0 | 41-45 | RENAL DONOR |
| 1256 | 0 | 0 | 31-35 | RENAL DONOR |
| 1257 | 0 | 0 | 51-55 | RENAL DONOR |

| | | | | |
|---|---|---|---|---|
| 1258 | 1 | 0 | 56-60 | RENAL DONOR |
| 1259 | 0 | 0 | 21-25 | RENAL DONOR |
| 1260 | 1 | 0 | 56-60 | RENAL DONOR |
| 1261 | 0 | 0 | 41-45 | RENAL DONOR |
| 1262 | 1 | 0 | 41-45 | RENAL DONOR |
| 1263 | 1 | 0 | 56-60 | RENAL DONOR |
| 1264 | 0 | 0 | 51-55 | RENAL DONOR |
| 1265 | 0 | 0 | 41-45 | RENAL DONOR |
| 1266 | 1 | 0 | 36-40 | RENAL DONOR |
| 1267 | | | | RENAL DONOR |
| 1268 | 0 | 0 | 66-70 | RENAL DONOR |
| 1269 | 0 | 0 | 56-60 | RENAL DONOR |
| 1270 | 0 | 0 | 51-55 | RENAL DONOR |
| 1271 | 1 | 0 | 46-50 | RENAL DONOR |
| 1272 | 1 | 0 | 46-50 | RENAL DONOR |
| 1273 | 1 | 0 | 46-50 | RENAL DONOR |
| 1274 | | | | RENAL DONOR |
| 1275 | 0 | 0 | 26-30 | RENAL DONOR |
| 1276 | 1 | 0 | 56-60 | RENAL DONOR |
| 1277 | 1 | 0 | 56-60 | RENAL DONOR |
| 1278 | 1 | 0 | 26-30 | RENAL DONOR |
| 1279 | 1 | 0 | 56-60 | RENAL DONOR |
| 1280 | 1 | 0 | 41-45 | RENAL DONOR |
| 1281 | 0 | 0 | 61-65 | RENAL DONOR |
| 1282 | 0 | 0 | 31-35 | RENAL DONOR |
| 1283 | 1 | 0 | 56-60 | RENAL DONOR |
| 1284 | 0 | 0 | 56-60 | RENAL DONOR |
| 1285 | 1 | 0 | 46-50 | RENAL DONOR |
| 1286 | 1 | 0 | 26-30 | RENAL DONOR |
| 1287 | 1 | 0 | 36-40 | RENAL DONOR |
| 1288 | 0 | 0 | 31-35 | RENAL DONOR |
| 1289 | 0 | 0 | 31-35 | RENAL DONOR |
| 1290 | 0 | 0 | 51-55 | RENAL DONOR |
| 1291 | 1 | 0 | 56-60 | RENAL DONOR |
| 1292 | 0 | 0 | 46-50 | RENAL DONOR |
| 1293 | 1 | 0 | 56-60 | RENAL DONOR |
| 1294 | 0 | 0 | 61-65 | RENAL DONOR |
| 1295 | 1 | 0 | 51-55 | RENAL DONOR |
| 1296 | 1 | 0 | 66-70 | RENAL DONOR |
| 1297 | 0 | 0 | 41-45 | RENAL DONOR |
| 1298 | 1 | 0 | 51-55 | RENAL DONOR |
| 1299 | 0 | 0 | 51-55 | RENAL DONOR |
| 1300 | | | | RENAL DONOR |
| 1301 | 1 | 0 | 66-70 | RENAL DONOR |
| 1302 | 1 | 0 | 61-65 | RENAL DONOR |
| 1303 | 1 | 0 | 46-50 | RENAL DONOR |
| 1304 | 1 | 0 | 31-35 | RENAL DONOR |
| 1305 | 1 | 0 | 56-60 | RENAL DONOR |
| 1306 | 1 | 0 | 36-40 | RENAL DONOR |
| 1307 | 1 | 0 | 31-35 | RENAL DONOR |
| 1308 | 1 | 0 | 51-55 | RENAL DONOR |
| 1309 | 1 | 0 | 46-50 | RENAL DONOR |
| 1310 | 1 | 0 | 46-50 | RENAL DONOR |
| 1311 | 1 | 0 | 61-65 | RENAL DONOR |
| 1312 | 0 | 0 | 51-55 | RENAL DONOR |
| 1313 | 1 | 0 | 36-40 | RENAL DONOR |
| 1314 | 0 | 0 | 36-40 | RENAL DONOR |
| 1315 | 1 | 0 | 16-20 | RENAL DONOR |
| 1316 | 1 | 0 | 56-60 | RENAL DONOR |
| 1317 | 1 | 0 | 36-40 | RENAL DONOR |
| 1318 | 0 | 0 | 56-60 | RENAL DONOR |
| 1319 | 1 | 0 | 36-40 | RENAL DONOR |
| 1320 | | | | RENAL DONOR |
| 1321 | 1 | 0 | 46-50 | RENAL DONOR |
| 1322 | 1 | 0 | 31-35 | RENAL DONOR |
| 1323 | 0 | 0 | 21-25 | RENAL DONOR |
| 1324 | 0 | 0 | 56-60 | RENAL DONOR |
| 1325 | 1 | 0 | 51-55 | RENAL DONOR |
| 1326 | 1 | 0 | 51-55 | RENAL DONOR |
| 1327 | 0 | 0 | 46-50 | RENAL DONOR |
| 1328 | 1 | 0 | 66-70 | RENAL DONOR |
| 1329 | 0 | 0 | 31-35 | RENAL DONOR |
| 1330 | | | | RENAL DONOR |
| 1331 | 1 | | >90 | RENAL DONOR |
| 1332 | 1 | 0 | 41-45 | RENAL DONOR |
| 1333 | 1 | 0 | 21-25 | RENAL DONOR |
| 1334 | 0 | 0 | 46-50 | RENAL DONOR |
| 1335 | 0 | 0 | 36-40 | RENAL DONOR |
| 1336 | 1 | 0 | 41-45 | RENAL DONOR |
| 1337 | 1 | 0 | 46-50 | RENAL DONOR |
| 1338 | 0 | 0 | 51-55 | RENAL DONOR |
| 1339 | 0 | 0 | 41-45 | RENAL DONOR |
| 1340 | 0 | 0 | 36-40 | RENAL DONOR |
| 1341 | 1 | 0 | 21-25 | RENAL DONOR |
| 1342 | 1 | 0 | 56-60 | RENAL DONOR |
| 1343 | 0 | 0 | 51-55 | RENAL DONOR |
| 1344 | 0 | 0 | 26-30 | RENAL DONOR |
| 1345 | 0 | 0 | 21-25 | RENAL DONOR |
| 1346 | 1 | 0 | 46-50 | RENAL DONOR |
| 1347 | 1 | 0 | 66-70 | RENAL DONOR |
| 1348 | 1 | 0 | 36-40 | RENAL DONOR |
| 1349 | 1 | 0 | 36-40 | RENAL DONOR |
| 1350 | 0 | 0 | 66-70 | RENAL DONOR |
| 1351 | 0 | 0 | 41-45 | RENAL DONOR |
| 1352 | 1 | 0 | 51-55 | RENAL DONOR |
| 1353 | 0 | 0 | 51-55 | RENAL DONOR |
| 1354 | 1 | 0 | 46-50 | RENAL DONOR |

| | | | | |
|---|---|---|---|---|
| 1355 | 1 | 0 | 51-55 | RENAL DONOR |
| 1356 | 1 | 0 | 31-35 | RENAL DONOR |
| 1357 | 0 | 0 | 46-50 | RENAL DONOR |
| 1358 | 0 | 0 | 41-45 | RENAL DONOR |
| 1359 | 0 | 0 | 36-40 | RENAL DONOR |
| 1360 | | | | RENAL DONOR |
| 1361 | 0 | 0 | 41-45 | RENAL DONOR |
| 1362 | 1 | 0 | 41-45 | RENAL DONOR |
| 1363 | 1 | 0 | 56-60 | RENAL DONOR |
| 1364 | 1 | 0 | 41-45 | RENAL DONOR |
| 1365 | 1 | 0 | 46-50 | RENAL DONOR |
| 1366 | 1 | 0 | 31-35 | RENAL DONOR |
| 1367 | 1 | 0 | 61-65 | RENAL DONOR |
| 1368 | 1 | 0 | 36-40 | RENAL DONOR |
| 1369 | 1 | 0 | 56-60 | RENAL DONOR |
| 1370 | 0 | 0 | 46-50 | RENAL DONOR |
| 1371 | 1 | 0 | 41-45 | RENAL DONOR |
| 1372 | 1 | 0 | 61-65 | RENAL DONOR |
| 1373 | 1 | 0 | 31-35 | RENAL DONOR |
| 1374 | 0 | 0 | 31-35 | RENAL DONOR |
| 1375 | 0 | 0 | 66-70 | RENAL DONOR |
| 1376 | 0 | 0 | 51-55 | RENAL DONOR |
| 1377 | 1 | 0 | 56-60 | RENAL DONOR |
| 1378 | 1 | 0 | 46-50 | RENAL DONOR |
| 1379 | 1 | 0 | 36-40 | RENAL DONOR |
| 1380 | 0 | 0 | 56-60 | RENAL DONOR |
| 1381 | 0 | 0 | 46-50 | RENAL DONOR |
| 1382 | 0 | 0 | 56-60 | RENAL DONOR |
| 1383 | 1 | 0 | 61-65 | RENAL DONOR |
| 1384 | 1 | 0 | 56-60 | RENAL DONOR |
| 1385 | 0 | 1 | 41-45 | RENAL DONOR |
| 1386 | 0 | 0 | 56-60 | RENAL DONOR |
| 1387 | 0 | 0 | 36-40 | RENAL DONOR |
| 1388 | 1 | 0 | 41-45 | RENAL DONOR |
| 1389 | 1 | 0 | 56-60 | RENAL DONOR |
| 1390 | 0 | 0 | 56-60 | RENAL DONOR |
| 1391 | 0 | 0 | 41-45 | RENAL DONOR |
| 1392 | 1 | 0 | 41-45 | RENAL DONOR |
| 1393 | 1 | 0 | 46-50 | RENAL DONOR |
| 1394 | 0 | 0 | 46-50 | RENAL DONOR |
| 1395 | 1 | 0 | 61-65 | RENAL DONOR |
| 1396 | 1 | 0 | 21-25 | RENAL DONOR |
| 1397 | 0 | 0 | 51-55 | RENAL DONOR |
| 1398 | 0 | 0 | 21-25 | RENAL DONOR |
| 1399 | 1 | 0 | 56-60 | RENAL DONOR |
| 1400 | 1 | 0 | 46-50 | RENAL DONOR |
| 1401 | 0 | 0 | 51-55 | RENAL DONOR |
| 1402 | 1 | 0 | 46-50 | RENAL DONOR |
| 1403 | 1 | 0 | 51-55 | RENAL DONOR |
| 1404 | 0 | 0 | 46-50 | RENAL DONOR |
| 1405 | 0 | 0 | 21-25 | RENAL DONOR |
| 1406 | 1 | 0 | 66-70 | RENAL DONOR |
| 1407 | 1 | 0 | 41-45 | RENAL DONOR |
| 1408 | 0 | 0 | 31-35 | RENAL DONOR |
| 1409 | 0 | 0 | 36-40 | RENAL DONOR |
| 1410 | 1 | 0 | 61-65 | RENAL DONOR |
| 1411 | 0 | 0 | 51-55 | RENAL DONOR |
| 1412 | 1 | 0 | 41-45 | RENAL DONOR |
| 1413 | 1 | 0 | 36-40 | RENAL DONOR |
| 1414 | 0 | 0 | 31-35 | RENAL DONOR |
| 1415 | 1 | 0 | 41-45 | RENAL DONOR |
| 1416 | 0 | 0 | 66-70 | RENAL DONOR |
| 1417 | 1 | 0 | 56-60 | RENAL DONOR |
| 1418 | 0 | 0 | 66-70 | RENAL DONOR |
| 1419 | 1 | 0 | 46-50 | RENAL DONOR |
| 1420 | 1 | 0 | 26-30 | RENAL DONOR |
| 1421 | 1 | 0 | 56-60 | RENAL DONOR |
| 1422 | 1 | 0 | 61-65 | RENAL DONOR |
| 1423 | 1 | 0 | 51-55 | RENAL DONOR |
| 1424 | 0 | 0 | 51-55 | RENAL DONOR |
| 1425 | 1 | 0 | 51-55 | RENAL DONOR |
| 1426 | 1 | 0 | 51-55 | RENAL DONOR |
| 1427 | 1 | 0 | 56-60 | RENAL DONOR |
| 1428 | 1 | 0 | 46-50 | RENAL DONOR |
| 1429 | 0 | 0 | 21-25 | RENAL DONOR |
| 1430 | 1 | 0 | 61-65 | RENAL DONOR |
| 1431 | 1 | 0 | 61-65 | RENAL DONOR |
| 1432 | 0 | 0 | 61-65 | RENAL DONOR |
| 1433 | 1 | 0 | 61-65 | RENAL DONOR |
| 1434 | 0 | 0 | 66-70 | RENAL DONOR |
| 1435 | 1 | 0 | 56-60 | RENAL DONOR |
| 1436 | 1 | 0 | 56-60 | RENAL DONOR |
| 1437 | 1 | 0 | 51-55 | RENAL DONOR |
| 1438 | 1 | 0 | 51-55 | RENAL DONOR |
| 1439 | 1 | 0 | 51-55 | RENAL DONOR |
| 1440 | 1 | 0 | 66-70 | RENAL DONOR |
| 1441 | 0 | 0 | 51-55 | RENAL DONOR |
| 1442 | 1 | 0 | 46-50 | RENAL DONOR |
| 1443 | 0 | 0 | 51-55 | RENAL DONOR |
| 1444 | 0 | 0 | 46-50 | RENAL DONOR |
| 1445 | 1 | 0 | 31-35 | RENAL DONOR |
| 1446 | 0 | 0 | 51-55 | RENAL DONOR |
| 1447 | 1 | 0 | 56-60 | RENAL DONOR |
| 1448 | 0 | 0 | 51-55 | RENAL DONOR |
| 1449 | 0 | 0 | 71-75 | RENAL DONOR |
| 1450 | 1 | 0 | 71-75 | RENAL DONOR |
| 1451 | 0 | 0 | 51-55 | RENAL DONOR |

| | | | | | | |
|---|---|---|---|---|---|---|
| 1452 | 1 | 0 | 51-55 | RENAL DONOR | | |
| 1453 | 1 | 0 | 56-60 | RENAL DONOR | | |
| 1454 | 0 | 0 | 51-55 | RENAL DONOR | | |
| 1455 | 0 | 0 | 51-55 | RENAL DONOR | | |
| 1456 | 1 | 0 | 51-55 | RENAL DONOR | | |
| 1457 | 1 | 0 | 46-50 | RENAL DONOR | | |
| 1458 | 1 | 0 | 46-50 | RENAL DONOR | | |
| 1459 | 1 | 0 | 46-50 | RENAL DONOR | | |
| 1460 | 1 | 0 | 46-50 | RENAL DONOR | | |
| 1461 | 0 | 0 | 61-65 | RENAL DONOR | | |
| 1462 | 1 | 0 | 56-60 | RENAL DONOR | | |
| 1463 | 1 | 0 | 56-60 | RENAL DONOR | | |
| 1464 | 1 | 0 | 56-60 | RENAL DONOR | | |
| 1465 | 1 | 0 | 56-60 | RENAL DONOR | | |
| 1466 | 0 | 0 | 56-60 | RENAL DONOR | | |
| 1467 | 0 | 0 | 56-60 | RENAL DONOR | | |
| 1468 | 0 | 0 | 51-55 | RENAL DONOR | | |
| 1469 | 1 | 0 | 56-60 | RENAL DONOR | | |
| 1470 | 1 | 0 | 56-60 | RENAL DONOR | | |
| 1471 | 0 | 0 | 46-50 | RENAL DONOR | | |
| 1472 | 0 | 0 | 46-50 | RENAL DONOR | | |
| 1473 | 1 | 0 | 56-60 | RENAL DONOR | | |
| 1474 | 1 | 0 | 61-65 | RENAL DONOR | | |
| 1475 | 1 | 0 | 56-60 | RENAL DONOR | | |
| 1476 | 1 | 0 | 56-60 | RENAL DONOR | | |
| 1477 | 0 | 0 | 41-45 | RENAL DONOR | | |
| 1478 | 1 | 0 | 41-45 | RENAL DONOR | | |
| 1479 | 1 | 0 | 46-50 | RENAL DONOR | | |
| 1480 | 0 | 0 | 51-55 | RENAL DONOR | | |
| 1481 | 1 | 0 | 51-55 | RENAL DONOR | | |
| 1482 | 0 | 0 | 41-45 | RENAL DONOR | | |
| 1483 | 0 | 0 | 36-40 | RENAL DONOR | | |
| 1484 | 1 | 0 | 51-55 | RENAL DONOR | | |
| 1485 | 0 | 0 | 56-60 | RENAL DONOR | | |
| 1486 | 1 | 0 | 51-55 | RENAL DONOR | | |
| 1487 | 0 | 0 | 56-60 | RENAL DONOR | | |
| 1488 | 1 | 0 | 41-45 | RENAL DONOR | | |
| 1489 | 1 | 0 | 41-45 | RENAL DONOR | | |
| 1490 | 0 | 0 | 41-45 | RENAL DONOR | | |
| 1491 | 1 | 0 | 36-40 | RENAL DONOR | | |
| 1492 | 0 | 0 | 41-45 | RENAL DONOR | | |
| 1493 | 1 | 0 | 36-40 | RENAL DONOR | | |
| 1494 | 0 | 0 | 56-60 | RENAL DONOR | | |
| 1495 | 1 | 0 | 66-70 | RENAL DONOR | | |
| 1496 | 1 | 0 | 46-50 | RENAL DONOR | | |
| 1497 | 0 | 0 | 51-55 | RENAL DONOR | | |
| 1498 | 1 | 0 | 56-60 | RENAL DONOR | | |
| 1499 | 0 | 0 | 31-35 | RENAL DONOR | | |
| 1500 | 0 | 0 | 66-70 | PDAC | 0 | 4.8 |
| 1501 | 1 | 0 | 66-70 | PNET | 0 | 3.2 |
| 1502 | 0 | 0 | 71-75 | PNET | 0 | 2.2 |
| 1503 | 1 | 0 | 66-70 | PNET | 0 | 10.5 |
| 1504 | 1 | 0 | 66-70 | PNET | 0 | 6 |
| 1505 | 1 | 0 | 56-60 | PNET | 0 | 3.2 |
| 1506 | 0 | 0 | 56-60 | PNET | 0 | 1.8 |
| 1507 | 0 | 0 | 61-65 | PNET | 0 | 3.5 |
| 1508 | 1 | 1 | 76-80 | PDAC | 0 | 3 |
| 1509 | 0 | 1 | 41-45 | PNET | 0 | 6 |
| 1510 | 0 | 0 | 56-60 | PNET | 0 | 3 |
| 1511 | 1 | 0 | 56-60 | PNET | 0 | 4.5 |
| 1512 | 0 | 0 | 66-70 | PNET | 1 | 2.2 |
| 1513 | 0 | 0 | 71-75 | PNET | 0 | 7 |
| 1514 | 1 | 0 | 31-35 | PNET | 0 | 1.5 |
| 1515 | 0 | 0 | 41-45 | PNET | 0 | 2.4 |
| 1516 | 1 | 0 | 61-65 | PNET | 0 | 0.8 |
| 1517 | 1 | 0 | 41-45 | PNET | 0 | 1.8 |
| 1518 | 0 | 0 | 61-65 | PNET | 0 | 7 |
| 1519 | 0 | 0 | 56-60 | PNET | 0 | 6 |
| 1520 | 0 | 0 | 66-70 | PNET | 0 | 2 |
| 1521 | 0 | 0 | 61-65 | PNET | 0 | 2 |
| 1522 | 0 | 0 | 61-65 | PNET | 0 | 3.5 |
| 1523 | 1 | 0 | 61-65 | PNET | 0 | 1.4 |
| 1524 | 0 | 0 | 66-70 | PNET | 0 | 7.3 |
| 1525 | 1 | 0 | 66-70 | PNET | 0 | 10 |
| 1526 | 0 | 1 | 81-85 | PNET | 0 | 4 |
| 1527 | 0 | 0 | 31-35 | PNET | 0 | 1.3 |
| 1528 | 1 | 0 | 56-60 | PNET | 0 | 1 |
| 1529 | 0 | 0 | 56-60 | PNET | 0 | 0.9 |
| 1530 | 1 | 0 | 51-55 | PNET | 0 | 1.1 |
| 1531 | 1 | 0 | 26-30 | PNET | 0 | 1 |
| 1532 | 0 | 0 | 66-70 | PNET | 0 | 6 |
| 1533 | 0 | 0 | 41-45 | PNET | 0 | 2.7 |
| 1534 | 0 | 0 | 46-50 | PNET | 0 | 0.8 |
| 1535 | 0 | 0 | 56-60 | PNET | 0 | 0.9 |
| 1536 | 1 | 0 | 31-35 | PNET | 0 | 1.2 |
| 1537 | 1 | 0 | 46-50 | PNET | 0 | 15.3 |
| 1538 | 1 | 1 | 51-55 | PNET | 0 | 2.5 |
| 1539 | 1 | 0 | 46-50 | PNET | 0 | 1.5 |
| 1540 | 0 | 0 | 61-65 | PNET | 0 | 2.4 |
| 1541 | 0 | 0 | 56-60 | PNET | 0 | 1.3 |
| 1542 | 1 | 0 | 36-40 | PNET | 0 | 11.3 |
| 1543 | 1 | 0 | 36-40 | PNET | 0 | 1.1 |
| 1544 | 0 | 0 | 21-25 | PNET | 0 | 4.5 |
| 1545 | 1 | 0 | 31-35 | PNET | 0 | 3 |
| 1546 | 0 | 0 | 56-60 | PNET | 0 | 1.6, 1.5 |
| 1547 | 1 | 0 | 46-50 | PNET | 0 | 1.5 |
| 1548 | 1 | 0 | 71-75 | PNET | 0 | 2.5 |

| | | | | | | |
|---|---|---|---|---|---|---|
| 1549 | 0 | 0 | 71-75 | PNET | 0 | 1.9 |
| 1550 | 0 | 0 | 61-65 | PNET | 0 | 2.5 |
| 1551 | 1 | 1 | 66-70 | PNET | 0 | 1.5 |
| 1552 | 1 | 0 | 41-45 | PNET | 0 | 0.8 |
| 1553 | 1 | 0 | 56-60 | PNET | 1 | 7.5 |
| 1554 | 0 | 0 | 71-75 | PNET | 0 | 1.2 and 0.4 |
| 1555 | 1 | 0 | 51-55 | PNET | 0 | 1.4 |
| 1556 | 1 | 0 | 51-55 | PNET | 0 | 0.7 |
| 1557 | 0 | 0 | 66-70 | PNET | 0 | 5 |
| 1558 | 0 | 0 | 71-75 | PNET | 0 | 1.2 |
| 1559 | 0 | 0 | 46-50 | PNET | 0 | 6.2 |
| 1560 | 0 | 0 | 61-65 | PNET | 0 | 1.7 |
| 1561 | 0 | 0 | 51-55 | PNET | 0 | 1.7 |
| 1562 | 0 | 0 | 66-70 | PNET | 0 | 0.9 |
| 1563 | 0 | 0 | 66-70 | PNET | 0 | 1.8 |
| 1564 | 1 | 0 | 56-60 | PNET | 0 | 1.9 |
| 1565 | 1 | 0 | 71-75 | PNET | 0 | 5.2 |
| 1566 | 1 | 0 | 56-60 | PNET | 0 | 2.8 |
| 1567 | 1 | 0 | 66-70 | PNET | 0 | 0.8 |
| 1568 | 0 | 0 | 51-55 | PNET | 0 | 2 |
| 1569 | 1 | 0 | 31-35 | PNET | 0 | 1 |
| 1570 | 0 | 0 | 26-30 | PNET | 0 | 2.3 |
| 1571 | 1 | 0 | 51-55 | PNET | 0 | 5.5 |
| 1572 | 0 | 0 | 46-50 | PNET | 0 | 9.5 |
| 1573 | 1 | 0 | 61-65 | PNET | 0 | 2.5 |
| 1574 | 1 | 0 | 66-70 | PNET | 0 | 3.3 |
| 1575 | 1 | 0 | 36-40 | PNET | 0 | 3.5 |
| 1576 | 0 | 0 | 46-50 | PNET | 0 | 5 |
| 1577 | 0 | 0 | 46-50 | PNET | 0 | 1 |
| 1578 | 1 | 0 | 61-65 | PNET | 0 | 4 |
| 1579 | 0 | 0 | 51-55 | PNET | 0 | 10.5 |
| 1580 | 0 | 0 | 76-80 | PNET | 0 | 1.2 |
| 1581 | 0 | 0 | 76-80 | PNET | 0 | 2.1 |
| 1582 | 1 | 0 | 61-65 | PNET | 0 | 1.5 |
| 1583 | 0 | 1 | 61-65 | PNET | 0 | 2.2 |
| 1584 | 0 | 0 | 46-50 | PNET | 0 | 2.4 |
| 1585 | 1 | 0 | 66-70 | PNET | 0 | 1.2 |
| 1586 | 0 | 0 | 71-75 | PNET | 0 | 4.8 |
| 1587 | 0 | 0 | 61-65 | PNET | 0 | 2.5 |
| 1588 | 0 | 0 | 51-55 | PNET | 0 | 7 |
| 1589 | 0 | 1 | 81-85 | PNET | 0 | 4.3 |
| 1590 | 1 | 0 | 51-55 | PNET | 0 | 8.7 |
| 1591 | 1 | 0 | 51-55 | PNET | 0 | 0.9 |
| 1592 | 1 | 0 | 66-70 | PNET | 0 | 5 |
| 1593 | 1 | 0 | 56-60 | PNET | 0 | 9 |
| 1594 | 0 | 0 | 61-65 | PNET | 0 | 8 |
| 1595 | 1 | 0 | 56-60 | PNET | 0 | 8.4 |
| 1596 | 1 | 0 | 31-35 | PNET | 0 | 3 |
| 1597 | 1 | 0 | 76-80 | IPMN | 0 | 0.2 |
| 1598 | 1 | 0 | 46-50 | PNET | 1 | 1.4 |
| 1599 | 1 | 0 | 66-70 | PNET | 0 | 1.5 |
| 1600 | 0 | 0 | 51-55 | PNET | 0 | 3.5 |
| 1601 | 1 | 0 | 51-55 | PNET | 0 | 7 |
| 1602 | 0 | 0 | 51-55 | PNET | 0 | 9.2 |
| 1603 | 1 | 0 | 56-60 | PNET | 0 | 1.5 |
| 1604 | 0 | 0 | 66-70 | PNET | 0 | 1.8 |
| 1605 | 0 | 0 | 56-60 | PNET | 0 | 1.5 |
| 1606 | 0 | 1 | 66-70 | PNET | 0 | 2.5 |
| 1607 | 0 | 0 | 46-50 | PNET | 0 | 2.2 |
| 1608 | 1 | 0 | 56-60 | PNET | 1 | 3 |
| 1609 | 1 | 0 | 61-65 | PNET | 0 | 2.2 |
| 1610 | 1 | 0 | 76-80 | PNET | 0 | 5 |
| 1611 | 1 | 0 | 61-65 | PNET | 0 | 2 |
| 1612 | 0 | 0 | 56-60 | PNET | 0 | 2.1 |
| 1613 | 0 | 0 | 61-65 | PNET | 0 | 1.5 |
| 1614 | 0 | 0 | 66-70 | PNET | 0 | 1.1 |
| 1615 | 0 | 0 | 61-65 | PNET | 0 | 0.5 |
| 1616 | 0 | 0 | 46-50 | PNET | 0 | 2.2 |
| 1617 | 1 | 0 | 61-65 | PNET | 0 | 3.5 |
| 1618 | 0 | 0 | 36-40 | PNET | 0 | 0.8 |
| 1619 | 1 | 0 | 76-80 | PNET | 0 | 7.6 |
| 1620 | 0 | 0 | 16-20 | PNET | 1 | 2.5 |
| 1621 | 1 | 0 | 51-55 | PNET | 0 | 4.5 |
| 1622 | 0 | 0 | 41-45 | PNET | 1 | 3.5 |
| 1623 | 1 | 0 | 66-70 | PNET | 0 | 1.1 |
| 1624 | 0 | 1 | 76-80 | PNET | 0 | 5.5 |
| 1625 | 0 | 0 | 61-65 | PNET | 0 | 1.1 |
| 1626 | 0 | 0 | 66-70 | PNET | 0 | 3 |
| 1627 | 0 | 0 | 36-40 | PNET | 1 | 3.3 |
| 1628 | 0 | 0 | 56-60 | PNET | 0 | 5 |
| 1629 | 0 | 1 | 56-60 | PNET | 0 | 6 |
| 1630 | 1 | 0 | 46-50 | PNET | 0 | 4.6 |
| 1631 | 1 | 1 | 81-85 | PNET | 0 | 6.5 |
| 1632 | 0 | 0 | 71-75 | PNET | 1 | 4 |
| 1633 | 1 | 0 | 61-65 | PNET | 1 | 4.8 |
| 1634 | 0 | 0 | 61-65 | PNET | 0 | 3 |
| 1635 | 0 | 1 | 66-70 | PNET | 0 | 4 |
| 1636 | 0 | 0 | 61-65 | PNET | 0 | 1.1 |
| 1637 | 1 | 0 | 56-60 | PNET | 0 | 5 |
| 1638 | 0 | 0 | 61-65 | PNET | 0 | 0.9 |
| 1639 | 1 | 0 | 56-60 | PNET | 0 | 1 |
| 1640 | 0 | 0 | 36-40 | PNET | 0 | 8 |
| 1641 | 0 | 0 | 36-40 | PNET | 0 | 3 |
| 1642 | 1 | 0 | 41-45 | PNET | 0 | 4.2 |
| 1643 | 0 | 0 | 61-65 | PNET | 0 | 12.5 |
| 1644 | 0 | 0 | 71-75 | PNET | 0 | 2.1 |
| 1645 | 1 | 0 | 26-30 | PNET | 0 | 1.3 |

| | | | | | | |
|---|---|---|---|---|---|---|
| 1646 | 0 | 0 | 66-70 | PNET | 0 | 2.4 |
| 1647 | 1 | 0 | 61-65 | PNET | 0 | 1.6 |
| 1648 | 0 | 0 | 41-45 | PNET | 0 | 2 |
| 1649 | 1 | 1 | 81-85 | PNET | 0 | 1.7 |
| 1650 | 1 | 0 | 46-50 | PNET | 1 | 1.7 |
| 1651 | 0 | 0 | 46-50 | PNET | 0 | 3.5 |
| 1652 | 1 | 1 | 46-50 | PNET | 0 | 3.8 |
| 1653 | 1 | 0 | 61-65 | PNET | 0 | 3.2 |
| 1654 | 0 | 0 | 56-60 | PNET | 0 | 3.6 |
| 1655 | 0 | 0 | 51-55 | PNET | 0 | 1.1 |
| 1656 | 0 | 0 | 66-70 | PNET | 0 | 3.7 |
| 1657 | 0 | 0 | 36-40 | PNET | 0 | 3.1, 2.8 |
| 1658 | 1 | 0 | 66-70 | PNET | 0 | 1.5 |
| 1659 | 1 | 0 | 31-35 | PNET | 0 | 1 |
| 1660 | 1 | 0 | 51-55 | PNET | 0 | 0.9 |
| 1661 | 0 | 0 | 36-40 | PNET | 0 | 2.1 |
| 1662 | 1 | 1 | 66-70 | PNET | 0 | 2.8 |
| 1663 | 0 | 0 | 21-25 | PNET | 0 | 2.8 |
| 1664 | 0 | 0 | 46-50 | PNET | 0 | 2.5 |
| 1665 | 0 | 0 | 61-65 | PNET | 0 | 0.7 |
| 1666 | 0 | 0 | 71-75 | PNET | 0 | 2.5 |
| 1667 | 0 | 1 | 61-65 | PNET | 0 | 4.5 |
| 1668 | 0 | 0 | 56-60 | PNET | 0 | 1 |
| 1669 | 0 | 0 | 41-45 | PNET | 0 | 1.8 |
| 1670 | 1 | 0 | 76-80 | PNET | 1 | 1.8 |
| 1671 | 0 | 0 | 56-60 | PNET | 0 | 1.2 |
| 1672 | 0 | 0 | 46-50 | PNET | 0 | 1.6 |
| 1673 | 1 | 0 | 61-65 | PNET | 0 | 1.9 |
| 1674 | 0 | 0 | 41-45 | PNET | 0 | 4 |
| 1675 | 1 | 1 | 61-65 | PNET | 0 | 3.5, 2 |
| 1676 | 0 | 0 | 61-65 | PNET | 0 | 4 |
| 1677 | 1 | 0 | 51-55 | PNET | 0 | 6 |
| 1678 | 0 | 0 | 31-35 | PNET | 0 | 3.8 |
| 1679 | 1 | 0 | 61-65 | PNET | 1 | 6 |
| 1680 | 0 | 0 | 46-50 | PNET | 0 | 1 |
| 1681 | 1 | 0 | 36-40 | PNET | 1 | 1.1 |
| 1682 | 0 | 0 | 66-70 | PNET | 0 | 1 |
| 1683 | 1 | 0 | 56-60 | PNET | 0 | 1.2 |
| 1684 | 1 | 0 | 41-45 | PNET | 0 | 1.5 |
| 1685 | 1 | 0 | 46-50 | PNET | 0 | 1 |
| 1686 | 1 | 1 | 56-60 | PNET | 0 | 1 |
| 1687 | 0 | 0 | 66-70 | PNET | 1 | 2.5 |
| 1688 | 1 | 0 | 46-50 | PNET | 0 | 2.5 |
| 1689 | 0 | 0 | 61-65 | PNET | 0 | 2.5 |
| 1690 | 1 | 0 | 76-80 | PNET | 0 | 4 |
| 1691 | 0 | 0 | 71-75 | PNET | 0 | 2.6 |
| 1692 | 0 | 0 | 61-65 | PNET | 0 | 2 |
| 1693 | 0 | 0 | 46-50 | PNET | 0 | 0.2 |
| 1694 | 1 | 0 | 36-40 | PNET | 0 | 0.6 |
| 1695 | 0 | 0 | 66-70 | PNET | 0 | 1.1 |
| 1696 | 1 | 0 | 41-45 | PNET | 0 | 3 |
| 1697 | 0 | 0 | 56-60 | PNET | 0 | 2.6 |
| 1698 | 1 | 0 | 51-55 | PNET | 0 | 1.5 |
| 1699 | 1 | 0 | 76-80 | PNET | 0 | 2.6 |
| 1700 | 0 | 0 | 71-75 | PNET | 0 | 0.9 |
| 1701 | 0 | 0 | 61-65 | PNET | 0 | 9 |
| 1702 | 1 | 0 | 56-60 | PNET | 0 | 1.1 |
| 1703 | 0 | 0 | 66-70 | PNET | 0 | 1.3 |
| 1704 | 1 | 0 | 46-50 | PNET | 0 | 2.5 |
| 1705 | 1 | 0 | 56-60 | PNET | 0 | 2.3 |
| 1706 | 1 | 0 | 36-40 | PNET | 0 | 3.7 |
| 1707 | 0 | 0 | 71-75 | PNET | 0 | 1.5 |
| 1708 | 1 | 0 | 26-30 | PNET | 0 | 2 |
| 1709 | 0 | 0 | 46-50 | PNET | 0 | 4 |
| 1710 | 0 | 0 | 56-60 | PNET | 0 | 2.7 |
| 1711 | 1 | 0 | 31-35 | PNET | 0 | 5.5 |
| 1712 | 0 | 0 | 51-55 | PNET | 0 | 5.2 |
| 1713 | 0 | 0 | 61-65 | PNET | 0 | 5.5 |
| 1714 | 0 | 0 | 56-60 | PNET | 0 | 4 |
| 1715 | 0 | 0 | 46-50 | PNET | 0 | 1.8 |
| 1716 | 1 | 0 | 56-60 | PNET | 0 | 5.5 |
| 1717 | 0 | 0 | 71-75 | PNET | 0 | 1.8 |
| 1718 | 0 | 0 | 76-80 | PNET | 1 | 1.8 |
| 1719 | 1 | 0 | 46-50 | PNET | 0 | 1.5 |
| 1720 | 0 | 0 | 61-65 | PNET | 0 | 2.5 |
| 1721 | 0 | 0 | 56-60 | PNET | 0 | 2.4 |
| 1722 | 0 | 0 | 41-45 | PNET | 0 | 1.5 |
| 1723 | 1 | 0 | 46-50 | PNET | 0 | 0.7 |
| 1724 | 0 | 0 | 66-70 | PNET | 1 | 2 |
| 1725 | 1 | 0 | 61-65 | PNET | 0 | 2.5 |
| 1726 | 0 | 0 | 41-45 | PNET | 0 | 2.5 |
| 1727 | 1 | 0 | 66-70 | PNET | 0 | 6 |
| 1728 | 0 | 1 | 61-65 | PNET | 0 | 4.5 |
| 1729 | 1 | 0 | 56-60 | PNET | 0 | 4 |
| 1730 | 1 | 0 | 61-65 | PNET | 0 | 1.2 |
| 1731 | 1 | 0 | 66-70 | PNET | 0 | 3 |
| 1732 | 1 | 0 | 76-80 | PNET | 1 | 3.5 |
| 1733 | 1 | 0 | 46-50 | PNET | 0 | 2.5 |
| 1734 | 1 | 0 | 41-45 | PNET | 0 | 2.6 |
| 1735 | 0 | 0 | 61-65 | PNET | 0 | 1.3 |
| 1736 | 0 | 0 | 66-70 | PNET | 0 | 1.5 |
| 1737 | 1 | 0 | 51-55 | PNET | 0 | 4.5 |
| 1738 | 1 | 0 | 56-60 | PNET | 0 | 1.4 |
| 1739 | 0 | 0 | 71-75 | PNET | 0 | 1.2 |
| 1740 | 0 | 0 | 41-45 | PNET | 0 | 1.4 |
| 1741 | 1 | 0 | 31-35 | PNET | 0 | 1.7 |
| 1742 | 0 | 0 | 56-60 | PNET | 0 | 2 |

| | | | | | | |
|---|---|---|---|---|---|---|
| 1743 | 1 | 0 | 46-50 | PNET | 0 | 1.5 |
| 1744 | 1 | 1 | 76-80 | PNET | 0 | 3.5 |
| 1745 | 1 | 0 | 51-55 | PNET | 0 | 2.1 |
| 1746 | 0 | 0 | 56-60 | PNET | 0 | 4.5 |
| 1747 | 1 | 0 | 31-35 | PNET | 1 | 3 |
| 1748 | 0 | 1 | 46-50 | PNET | 0 | 2.5 |
| 1749 | 0 | 0 | 41-45 | PNET | 1 | 3.5 |
| 1750 | 0 | 0 | 71-75 | PNET | 0 | 3 |
| 1751 | 0 | 0 | 21-25 | PNET | 0 | 1.6 |
| 1752 | 1 | 0 | 21-25 | PNET | 0 | 0.9 |
| 1753 | 0 | 0 | 46-50 | PNET | 0 | 3 |
| 1754 | 1 | 1 | 66-70 | PNET | 0 | 2.8 |
| 1755 | 1 | 0 | 56-60 | PNET | 0 | 2.6 |
| 1756 | 1 | 0 | 61-65 | PNET | 0 | 7 |
| 1757 | 0 | 0 | 46-50 | PNET | 0 | 0.5 |
| 1758 | 1 | 0 | 56-60 | PNET | 0 | 2.7 |
| 1759 | 1 | 0 | 26-30 | PNET | 0 | 0.5 |
| 1760 | 1 | 0 | 46-50 | PNET | 0 | 2 |
| 1761 | 0 | 0 | 46-50 | PNET | 0 | 5 |
| 1762 | 1 | 0 | 46-50 | PNET | 0 | 3 |
| 1763 | 1 | 0 | 71-75 | PNET | 0 | 2.2 |
| 1764 | 0 | 0 | 46-50 | PNET | 0 | 2.1 |
| 1765 | 1 | 0 | 46-50 | PNET | 0 | 1 |
| 1766 | 1 | 0 | 51-55 | PNET | 1 | 4.5 |
| 1767 | 1 | 1 | 31-35 | PANIN | 1 | 0.3 |
| 1768 | 0 | 0 | 31-35 | PNET | 0 | 5.1 |
| 1769 | 1 | 0 | 46-50 | PNET | 0 | 2.5 |
| 1770 | 0 | 0 | 46-50 | PNET | 0 | 2.2 |
| 1771 | 0 | 0 | 66-70 | PNET | 0 | 3 |
| 1772 | 1 | 0 | 61-65 | PNET | 0 | 1.2 |
| 1773 | 0 | 0 | 46-50 | PNET | 0 | 5 |
| 1774 | 1 | 0 | 56-60 | PNET | 0 | 0.6 |
| 1775 | 1 | 0 | 66-70 | PNET | 0 | 7.7 |
| 1776 | 0 | 0 | 56-60 | PNET | 1 | 5.8 |
| 1777 | 1 | 0 | 76-80 | PNET | 0 | 1.3 |
| 1778 | 1 | 0 | 61-65 | PNET | 0 | 3.6 |
| 1779 | 1 | 0 | 51-55 | PNET | 0 | 2.5 |
| 1780 | 0 | 0 | 56-60 | PNET | 0 | 12.4 |
| 1781 | 0 | 0 | 51-55 | PNET | 0 | 1.2 |
| 1782 | 1 | 0 | 61-65 | PNET | 1 | 1.9 |
| 1783 | 0 | 0 | 51-55 | PNET | 0 | 8.1 |
| 1784 | 0 | 0 | 71-75 | Cyst | 0 | 2.3 |
| 1785 | 1 | 0 | 41-45 | Cyst | 0 | 0.8 |
| 1786 | 0 | 0 | 66-70 | Cyst | 0 | "tiny" |
| 1787 | 0 | 0 | 61-65 | PNET | 0 | 0.8 |
| 1788 | 1 | 0 | 41-45 | Cyst | 0 | 3.1 |
| 1789 | 0 | 0 | 66-70 | Cyst | 0 | 5 |
| 1790 | 1 | 0 | 81-85 | IMPN | 0 | 4.2 |
| 1791 | 1 | 0 | 66-70 | Serous cystadenoma | 0 | 3.4 |
| 1792 | 1 | 0 | 76-80 | IMPN | 0 | 2 |
| 1793 | 1 | 0 | 36-40 | IMPN | 0 | 6.2 |
| 1794 | 1 | 0 | 56-60 | Mucinous cystic neoplasm | 0 | 1.6 |
| 1795 | 0 | 0 | 81-85 | Cyst | 0 | 6.3 |
| 1796 | 0 | 0 | 81-85 | Cyst | 0 | 4.3 |
| 1797 | 1 | 0 | 36-40 | Cyst | 0 | 4.8 |
| 1798 | 1 | 0 | 81-85 | Cyst | 0 | 8.6 |
| 1799 | 1 | 0 | 71-75 | Cyst | 0 | 8.2 |
| 1800 | 0 | 0 | 71-75 | Cyst | 0 | 4.2 |
| 1801 | 1 | 0 | 56-60 | Cyst | 0 | 5.5 |
| 1802 | 1 | 0 | 41-45 | Cyst | 0 | 4.8 |
| 1803 | 1 | 0 | 41-45 | Cyst | 0 | 3.2 |
| 1804 | 1 | 0 | 71-75 | Cyst | 0 | 3.3 |
| 1805 | 1 | 0 | 26-30 | Cyst | 0 | 3 |
| 1806 | 1 | 0 | 66-70 | serous adenoma | 0 | 3 |
| 1807 | 1 | 0 | 66-70 | IMPN | 1 | 6.5 |
| 1808 | 1 | 0 | 46-50 | Cyst | 0 | 3.7 |
| 1809 | 1 | 0 | 56-60 | Cyst | 0 | 8 |
| 1810 | 1 | 0 | 51-55 | serous cystadenoma | 1 | 11.9 |
| 1811 | 1 | 0 | 26-30 | Cyst | 0 | 4.3 |
| 1812 | 1 | 0 | 41-45 | Cyst | 1 | 3.7 |
| 1813 | 1 | 0 | 61-65 | IPMN | 0 | 7.3 |
| 1814 | 1 | 0 | 81-85 | Cyst | 0 | 5.3 |
| 1815 | 1 | 0 | 36-40 | Mucinous cystic neoplasn | 0 | 1.3 |
| 1816 | 1 | 0 | 41-45 | Cyst | 0 | 4.4 |
| 1817 | 1 | 0 | 66-70 | IPMN | 0 | 4.5 |
| 1818 | 1 | 0 | 71-75 | IPMN | 0 | 4.5 |
| 1819 | 0 | 0 | 61-65 | Cyst | 0 | 10.8 |
| 1820 | 0 | 0 | 66-70 | Cyst | 0 | 2.9 |
| 1821 | 0 | 0 | 66-70 | PNET | 0 | 0.9 |
| 1822 | 1 | 0 | 71-75 | Cyst | 0 | 2.4 |
| 1823 | 0 | 0 | 46-50 | Cyst | 0 | 3.3 |
| 1824 | 1 | 0 | 61-65 | serous cystadenoma | 0 | 4.9 |
| 1825 | 0 | 0 | 86-90 | Cyst | 0 | 5.4 |
| 1826 | 1 | 0 | 56-60 | Cyst | 0 | 3.9 |
| 1827 | 1 | 0 | 76-80 | Cyst | 0 | 6.9 |
| 1828 | 1 | 0 | 66-70 | Cyst | 0 | 4.1 |
| 1829 | 0 | 0 | 26-30 | Cyst | 0 | 7.7 |
| 1830 | 1 | 0 | 36-40 | serous cystadenoma | 0 | 3.2 |
| 1831 | 0 | 1 | 76-80 | IPMN | 0 | 2.5 |
| 1832 | 0 | 0 | 61-65 | Cyst | 0 | 4 |
| 1833 | 0 | 0 | 71-75 | Cyst | 0 | 5.2 |
| 1834 | 1 | 0 | 56-60 | Cyst | 0 | 3.2 |
| 1835 | 1 | 0 | 56-60 | Cyst | 0 | 3.5 |
| 1836 | 1 | 0 | 46-50 | Cyst | 0 | 3.7 |
| 1837 | 0 | 1 | 51-55 | Cyst | 0 | 4.6 |
| 1838 | 1 | 0 | 36-40 | Cyst | 0 | 4.4 |
| 1839 | 0 | 0 | 61-65 | Cyst | 0 | 1.1 |

| | | | | | | |
|---|---|---|---|---|---|---|
| 1840 | 1 | 0 | 76-80 | Cyst | 0 | 4.2 |
| 1841 | 1 | 0 | 71-75 | Cyst | 0 | 1.4 |
| 1842 | 1 | 0 | 46-50 | Cyst | 0 | 1.9 |
| 1843 | 1 | 0 | 71-75 | Cyst | 0 | 4 |
| 1844 | 1 | 0 | 61-65 | Cyst | 0 | 3.6 |
| 1845 | 0 | 0 | 61-65 | Cyst | 0 | 1.8 |
| 1846 | 0 | 0 | 46-50 | Cyst | 0 | 4.3 |
| 1847 | 1 | 0 | 51-55 | Cyst | 1 | 3.3 |
| 1848 | 0 | 0 | 66-70 | Cyst | 0 | 2 |
| 1849 | 1 | 0 | 61-65 | Cyst | 0 | 11.1 |
| 1850 | 1 | 0 | 66-70 | Cyst | 0 | 3 |
| 1851 | 1 | 0 | 51-55 | Cyst | 0 | 5 |
| 1852 | 1 | 0 | 86-90 | Cyst | 0 | multiple |
| 1853 | 1 | 0 | 46-50 | Cyst | 0 | 3.4 |
| 1854 | 0 | 0 | 61-65 | Cyst | 0 | 2.2 |
| 1855 | 1 | 0 | 81-85 | Cyst | 0 | 4.1 |
| 1856 | 1 | 0 | 66-70 | Cyst | 0 | 0.8 |
| 1857 | 1 | 0 | 56-60 | Cyst | 0 | 1 |
| 1858 | 0 | 0 | 51-55 | Cyst | 0 | 1.5 |
| 1859 | 0 | 1 | >90 | Cyst | 0 | 3.2 |
| 1860 | 0 | 1 | 81-85 | Cyst | 0 | 3.6 |
| 1861 | 1 | 0 | 31-35 | Cyst | 0 | 3.5 |
| 1862 | 1 | 0 | 36-40 | Cyst | 0 | 10 |
| 1863 | 1 | 0 | 66-70 | Cyst | 0 | 12.4 |
| 1864 | 1 | 0 | 61-65 | Cyst | 0 | 5.2 |
| 1865 | 1 | 0 | 46-50 | Cyst | 0 | 6.2 |
| 1866 | 1 | 0 | 61-65 | Cyst | 0 | 1.5 |
| 1867 | 1 | 0 | 51-55 | Cyst | 0 | 2 |
| 1868 | 1 | 0 | 61-65 | Cyst | 0 | 5 |
| 1869 | 1 | 0 | 61-65 | Cyst | 0 | 1.6 |
| 1870 | 0 | 0 | 41-45 | Cyst | 0 | 2.6 |
| 1871 | 1 | 0 | 66-70 | Cyst | 0 | 2.4 |
| 1872 | 0 | 0 | 61-65 | Cyst | 0 | 2.2 |
| 1873 | 1 | 0 | 56-60 | Cyst | 0 | 2.3 |
| 1874 | 1 | 0 | 46-50 | Cyst | 0 | 2.4 |
| 1875 | 1 | 0 | 31-35 | Cyst | 0 | 3 |
| 1876 | 1 | 0 | 66-70 | Cyst | 0 | 1.2 |
| 1877 | 1 | 0 | 66-70 | Cyst | 0 | 4 |
| 1878 | 0 | 0 | 56-60 | Cyst | 0 | 1 |
| 1879 | 1 | 0 | 46-50 | Cyst | 0 | 1.2 |
| 1880 | 0 | 0 | 61-65 | Cyst | 0 | 2.4 |
| 1881 | 0 | 0 | 51-55 | Cyst | 0 | 2.9 |
| 1882 | 1 | 0 | 61-65 | Cyst | 0 | 1.2 |
| 1883 | 1 | 0 | 66-70 | Cyst | 0 | 1.4 |
| 1884 | 0 | 0 | 86-90 | Cyst | 0 | 1.7 |
| 1885 | 1 | 0 | 61-65 | Cyst | 0 | 2.5 |
| 1886 | 1 | 0 | 56-60 | Cyst | 0 | 1.4 |
| 1887 | 1 | 0 | 81-85 | Cyst | 0 | 1.8 |
| 1888 | 1 | 0 | 76-80 | Cyst | 0 | 2.4 |
| 1889 | 0 | 0 | 66-70 | Cyst | 0 | 1.5 |
| 1890 | 1 | 0 | 51-55 | Cyst | 0 | 0.9 |
| 1891 | 0 | 0 | 61-65 | Cyst | 0 | 2.2 |
| 1892 | 1 | 0 | 66-70 | Cyst | 0 | 3.4 |
| 1893 | 1 | 0 | 61-65 | Cyst | 0 | 0.5 |
| 1894 | 0 | 0 | 66-70 | Cyst | 1 | 1.2 |
| 1895 | 0 | 0 | 76-80 | Cyst | 0 | multiple small |
| 1896 | 1 | 0 | 56-60 | Cyst | 0 | 1 |
| 1897 | 0 | 0 | 66-70 | Cyst | 0 | 3 |
| 1898 | 1 | 0 | 46-50 | Cyst | 0 | 0.7 |
| 1899 | 1 | 0 | 51-55 | Cyst | 0 | 1.2 |
| 1900 | 1 | 0 | 71-75 | Cyst | 1 | 2.1 |
| 1901 | 1 | 1 | 61-65 | Cyst | 0 | 1.8 |
| 1902 | 1 | 0 | 71-75 | Cyst | 0 | 1.3 |
| 1903 | 1 | 0 | 61-65 | Cyst | 0 | 2.6 |
| 1904 | 1 | 0 | 61-65 | Cyst | 0 | 0.6 |
| 1905 | 0 | 0 | 61-65 | Cyst | 0 | 2.5 |
| 1906 | 1 | 0 | 66-70 | Cyst | 1 | 2.4 |
| 1907 | 1 | 0 | 61-65 | Cyst | 0 | 2.5 |
| 1908 | 1 | 0 | 61-65 | Cyst | 0 | 0.9 |
| 1909 | 1 | 0 | 71-75 | Cyst | 1 | 1.5 |
| 1910 | 0 | 0 | 36-40 | Cyst | 0 | 3 |
| 1911 | 0 | 0 | 71-75 | Cyst | 0 | 2.2 |
| 1912 | 1 | 0 | 76-80 | Cyst | 0 | 1.5 |
| 1913 | 0 | 0 | 76-80 | Cyst | 0 | 1.6 |
| 1914 | 1 | 0 | 71-75 | Cyst | 0 | 1.1 |
| 1915 | 1 | 0 | 76-80 | Cyst | 0 | 1.4 |
| 1916 | 0 | 0 | 76-80 | Cyst | 0 | 1.9 |
| 1917 | 0 | 0 | 66-70 | Cyst | 0 | 1.5 |
| 1918 | 0 | 0 | 56-60 | Cyst | 0 | 1.7 |
| 1919 | 0 | 0 | 86-90 | Cyst | 0 | 1.4 |
| 1920 | 1 | 0 | 66-70 | Cyst | 0 | 0.8 |
| 1921 | 1 | 0 | 51-55 | Cyst | 0 | 1 |
| 1922 | 1 | 0 | 56-60 | Cyst | 0 | 0.7 |
| 1923 | 1 | 0 | 41-45 | Cyst | 0 | 2 |
| 1924 | 1 | 0 | 51-55 | Cyst | 0 | 2 |
| 1925 | 0 | 0 | 71-75 | Cyst | 0 | 1.4 |
| 1926 | 0 | 1 | 76-80 | Cyst | 0 | 0.9 |
| 1927 | 1 | 0 | 26-30 | Cyst | 1 | 4.4 |
| 1928 | 1 | 0 | 56-60 | Cyst | 0 | 0.5 |
| 1929 | 1 | 0 | 41-45 | Cyst | 0 | 1 |
| 1930 | 1 | 0 | 41-45 | Cyst | 0 | 1 |
| 1931 | 1 | 0 | 51-55 | Cyst | 0 | 1.8 |
| 1932 | 1 | 0 | 41-45 | Cyst | 0 | 1.6 |
| 1933 | 0 | 0 | 61-65 | Cyst | 0 | 1.5 |
| 1934 | 1 | 0 | 81-85 | Cyst | 0 | 1.7 |
| 1935 | 0 | 0 | 76-80 | Cyst | 0 | 1.8 |
| 1936 | 1 | 0 | 36-40 | Cyst | 0 | 1.6 |

| | | | | | | |
|---|---|---|---|---|---|---|
| 1937 | 0 | 0 | 66-70 | Cyst | 0 | 3.8 |
| 1938 | 1 | 0 | 51-55 | Cyst | 0 | 0.6 |
| 1939 | 1 | 0 | 56-60 | Cyst | 0 | 1.4 |
| 1940 | 1 | 0 | 51-55 | Cyst | 0 | 1.8 |
| 1941 | 1 | 0 | 21-25 | Cyst | 1 | 2.4 |
| 1942 | 1 | 0 | 71-75 | Cyst | 0 | 1.3 |
| 1943 | 1 | 0 | 46-50 | Cyst | 0 | 1.7 |
| 1944 | 0 | 0 | 66-70 | Cyst | 0 | 2.5 |
| 1945 | 1 | 0 | 66-70 | Cyst | 0 | 1.2 |
| 1946 | 1 | 0 | 66-70 | Cyst | 0 | 1.7 |
| 1947 | 0 | 0 | 56-60 | Cyst | 0 | 1.6 |
| 1948 | 1 | 0 | 66-70 | Cyst | 0 | 1.3 |
| 1949 | 1 | 0 | 56-60 | Cyst | 0 | 3.8 |
| 1950 | 1 | 0 | 66-70 | Cyst | 0 | 2.8 |
| 1951 | 1 | 0 | 71-75 | Cyst | 0 | 2.7 |
| 1952 | 1 | 0 | 66-70 | Cyst | 0 | 2.6 |
| 1953 | 1 | 0 | 56-60 | Cyst | 0 | 0.4 |
| 1954 | 0 | 0 | 66-70 | Cyst | 0 | 0.5 |
| 1955 | 1 | 0 | 56-60 | Cyst | 1 | 5.1 |
| 1956 | 1 | 0 | 71-75 | Cyst | 0 | 2.7 |
| 1957 | 0 | 0 | 71-75 | Cyst | 0 | 2.2 |
| 1958 | 1 | 0 | 51-55 | Cyst | 0 | 1.6 |
| 1959 | 0 | 0 | 76-80 | Cyst | 0 | 1.7 |
| 1960 | 1 | 0 | 66-70 | Cyst | 0 | 0.7 |
| 1961 | 1 | 0 | 56-60 | Cyst | 0 | 1.7 |
| 1962 | 0 | 0 | 81-85 | Cyst | 0 | 4 |
| 1963 | 1 | 0 | 81-85 | Cyst | 0 | 4.5 |
| 1964 | 1 | 0 | 76-80 | Cyst | 0 | 1.7 |
| 1965 | 1 | 0 | 51-55 | Cyst | 0 | 2.3 |
| 1966 | 0 | 0 | 61-65 | Cyst | 0 | 2.2 |
| 1967 | 1 | 0 | 71-75 | Cyst | 0 | 2.7 |
| 1968 | 1 | 0 | 66-70 | Cyst | 0 | 1.4 |
| 1969 | 0 | 0 | 56-60 | Cyst | 0 | 2.5 |
| 1970 | 0 | 0 | 76-80 | Cyst | 0 | 1.4 |
| 1971 | 1 | 0 | 51-55 | Cyst | 0 | 1.6 |
| 1972 | 1 | 0 | 66-70 | Cyst | 0 | 1.8 |
| 1973 | 0 | 0 | 81-85 | Cyst | 0 | 1.4 |
| 1974 | 1 | 0 | 61-65 | Cyst | 0 | 2.7 |
| 1975 | 1 | 0 | 56-60 | Cyst | 0 | 1.5 |
| 1976 | 1 | 0 | 51-55 | Cyst | 0 | 3.1 |
| 1977 | 1 | 0 | 56-60 | Cyst | 1 | 2.5 |
| 1978 | 0 | 0 | 76-80 | Cyst | 0 | 7.7 |
| 1979 | 0 | 0 | 66-70 | Cyst | 0 | 1.6 |
| 1980 | 1 | 0 | 51-55 | Cyst | 0 | 3 |
| 1981 | 1 | 0 | 66-70 | Cyst | 1 | 0.8 |
| 1982 | 0 | 0 | 71-75 | Cyst | 0 | 4.5 |
| 1983 | 1 | 0 | 46-50 | Cyst | 0 | 0.6 |
| 1984 | 1 | 0 | 46-50 | Cyst | 0 | 1.5 |
